# A human microbiome-derived therapeutic for ulcerative colitis promotes mucosal healing and immune homeostasis

**DOI:** 10.1101/2025.04.18.25325957

**Authors:** Páraic Ó Cuív, Johanna K. Ljungberg, Joyce Zhou, Michael Nissen, Annika Krueger, Joel Boyd, Mareike Bongers, Jimenez Loayza Jeimy, Charlotte Vivian, Ella Reich, Andrea Rabellino, Rhys Newell, Liang Fang, Samantha MacDonald, Alena Pribyl, Shandelle Caban, Huw McCarthy, Joanne Soh, Luke Reid, Blake Wills, Duncan McHale, David L.A. Wood, Ian H. Frazer, Nicola Angel, Hiram Chipperfield, Jakob Begun, Simon Keely, Gene W. Tyson, Philip Hugenholtz, Trent Munro, Lutz Krause

**Author notes:** Corresponding authors: Lutz Krause Páraic Ó Cuív.

## Abstract

The gut microbiome is central to the pathogenesis of ulcerative colitis (UC), and microbiome-derived therapeutics are a promising new treatment option. Using a metagenome-guided, large cohort-based approach, we identified *Hominenteromicrobium mulieris* as prevalent in healthy individuals but depleted in UC. Here, we present a Live Biotherapeutic Product (LBP) candidate, MAP 315, derived from a newly isolated strain of this species (MH27-2). In mouse colitis models, MH27-2 improved disease pathology, and accelerated gut healing, marked by epithelial restitution and reduced immune cell infiltration. *In vitro* studies showed that MH27-2 promotes mucosal healing through accelerated epithelial cell migration and proliferation, accelerated wound closure, and improved gut barrier integrity. Importantly, MH27-2 supports immune homeostasis through the promotion of regulatory T-cells, suppression of TL1A signaling, and induction of anti-inflammatory IL-10. Manufacturing processes were developed, and MH27-2 drug product (MAP 315) demonstrated to be safe and well-tolerated in a first-in-human Phase 1 clinical trial.

## Introduction

Inflammatory bowel disease (IBD), including the major subtypes ulcerative colitis (UC) and Crohn’s disease (CD), is a chronic, relapsing inflammatory disorder that significantly contributes to global disability and mortality^1–3^. During flare-ups, patients experience a variety of symptoms including gastrointestinal pain, diarrhea, bloody stools, and weight loss, with severe episodes often requiring hospitalisation^4^. Although the pathogenesis of IBD is poorly understood, it is believed to be driven by complex interactions between environmental, microbial, and immune-mediated factors in genetically susceptible individuals^2,5^.

There is currently no cure for IBD and current treatment goals aim to decrease the frequency and severity of inflammatory episodes to prevent progression of bowel damage and avoid surgery^6,7^. IBD is characterized by the loss of gut barrier function, and disrupted immune homeostasis, leading to excessive immune responses and chronic inflammation^2^. Consistent with this, there is an emerging emphasis on developing therapies that promote mucosal healing and intestinal homeostasis to achieve long-term remission^8–10^. Mucosal healing – the resolution of inflammation and restoration of gut barrier function – is associated with improved clinical outcomes^11–13^ and is therefore a critical therapeutic goal. Re-establishment of immune homeostasis is also an important treatment objective, as it is essential for maintaining tolerance to commensal microbes, food antigens, and self-antigens, through mechanisms such as differentiation of regulatory T cells, or secretion of anti-inflammatory cytokines^14–16^. While current therapies are effective in suppressing acute inflammatory responses, they have significant disadvantages, including patient immunosuppression and, in the case of biologicals, loss of efficacy through immunogenicity^17^. Therefore, there is an unmet clinical need for treatments that can restore gut barrier function and immune homeostasis, ultimately halting disease progression and facilitating long-term remission^10,18^.

In healthy individuals, gut bacteria are known to modulate specific biological functions critical for maintaining host health, including regulation of immune homeostasis and gut barrier integrity^19,20^. Structure-function alterations to the gut microbiome are a key feature of IBD, and restoration of these functions through the use of Live Biotherapeutic Products (LBPs) has emerged as a promising treatment strategy^5,21–25^. The efficacy of these cell-based therapies is often due to a combination of bioactive compounds and cellular interactions, which work together to modulate the host’s immune response, metabolism or microbiome composition. Their potential to replicate and engraft within the host allows for a sustained therapeutic effect that could be particularly advantageous in chronic conditions. Their multifaceted therapeutic capacity may further allow LBPs to address complex diseases in ways that conventional therapeutics, which typically act through a single mechanism, cannot.

Microbiome-derived therapeutics have shown promise in IBD, particularly for UC^26,27^. However, the existing limitations in microbial lead discovery, such as cultivation bias and low resolution sequencing-based approaches^28^, have constrained efforts to develop more effective products. In UC, the average age of diagnosis is 30 years with over 80% of patients having mild-to-moderate disease^29,30^. 5-aminosalicylates (5-ASAs) are the first line agent of choice for mild-to-moderate UC and they are prescribed to 90% of patients shortly after diagnosis^31^. However, 30-60% of UC patients fail to respond to the standard first line treatment and 5-10% of patients experience adverse events or drug intolerance^32–35^. For these patients, escalation to corticosteroid therapy or advanced small molecules and biologics is typically initiated. While corticosteroids and advanced therapies effectively target symptom suppression, they are associated with poor compliance, undesirable side-effects and low rates of mucosal healing. Indeed, prior steroid use in patients with acute severe UC is associated with a 75% increase in the likelihood of requiring colectomy within a year ^36^. There is a significant demand among UC patients and clinicians for new oral treatments that are safe and well tolerated, provide rapid symptomatic relief and sustainable remission with minimal loss of response.

Using a large human cohort and metagenome-guided approach, we previously proposed *Candidatus* Intestinicoccus colisanans (previously termed s *UBA1417 sp003531055*)^37^ as a target species for the development of a novel LBP to treat IBD^38^. However, a representative of the same species was recently isolated and validly named *Hominenteromicrobium mulieris*^39^, giving it nomenclatural priority. Although commonly occurring within the healthy human gut^37^, *H. mulieris* is significantly depleted in individuals with UC^38^, suggesting that restitution of this bacterial population in patients may alleviate disease. Here, we characterize MH27-2, a previously unreported strain of *H. mulieris* with potent therapeutic effects and an excellent safety profile. MH27-2 has been manufactured into the drug product MAP 315, which shows similar beneficial effects to MH27-2 *in vivo*. MAP 315 was evaluated in human adults in a randomized, double-blind, placebo-controlled Phase 1 clinical trial, revealing a strong safety and tolerability profile. Altogether, these data highlight the potential of MAP 315 as a safe and efficacious future therapy for UC.

## Results

### Isolation of *H. mulieris* strain MH27-2

*H. mulieris* is a strictly anaerobic bacterial species that is both abundant and prevalent in healthy individuals, but significantly depleted in patients with both major IBD subtypes, UC and CD (**Table S1**)^38^. Other related species do not significantly correlate with IBD (**Figure 1A**), highlighting the need for at least species-level resolution to select the most promising therapeutic candidates. *H. mulieris* belongs to the recently described family Acutalibacteraceae^40^ of the phylum Bacillota (previously Firmicutes), and is located in a cluster of genera mostly represented by as-yet-uncultivated bacteria (**Figure 1A**). The closest cultivated neighboring species are *Acutalibacter muris* and *Acutalibacter timonensis* that were isolated from mouse and human feces, respectively^40,41^ (**Figure 1B**). A novel axenic strain of this species, MH27-2 (**Figure S1A**), was obtained from the feces of a healthy human volunteer. This was achieved using genome-directed isolation, which leverages genomic data to design custom growth media, thereby supporting selective growth of target organisms^38,42^ (**Supplementary Information**).

**Figure 1.**
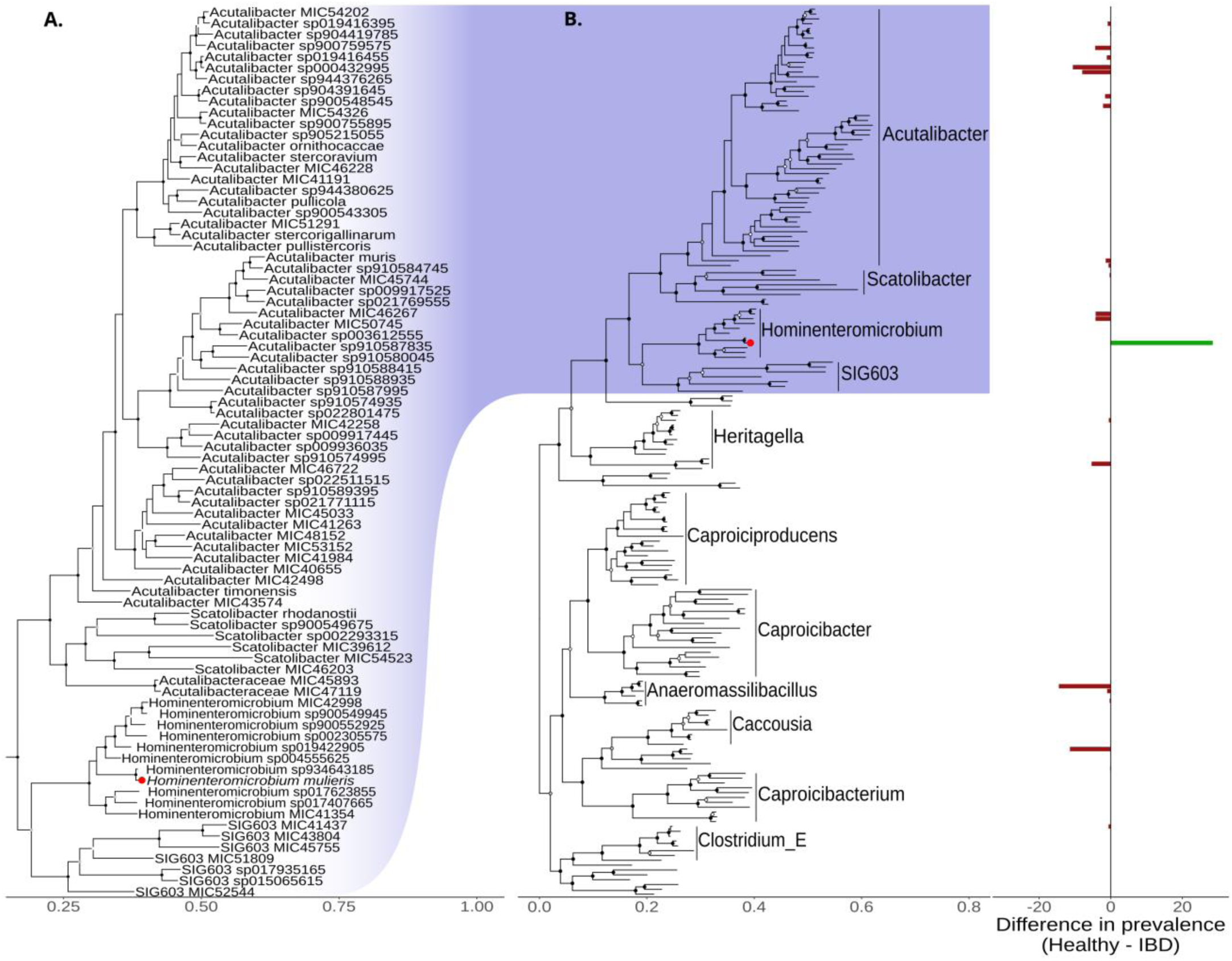
Phylogenetic trees of *Hominenteromicrobium mulieris* (s *UBA1417 sp003531055*) and surrounding species with their prevalence in IBD patients and healthy subjects. **A**. Genome tree constructed using publicly available genomes from NCBI using GTDBtk. Each leaf is a single representative of a species within the bacterial family *Acutalibacteraceae*. Colored nodes represent the bootstrap support values (white ≥50%, grey ≥75%, black 100%). **B.** Phylogenetic tree showing the broader phylogenetic context of *H. mulieris*. Genera with ≥5 member species are labelled, with *H. mulieris* indicated by a red node. Each leaf of the tree connects to a bar in the adjacent bar plot, which represents the difference in prevalence of that species in Healthy (n) vs IBD (n), where green bars represent positive associations (higher in healthy), and red bars show negative associations (higher in IBD).

### Genomic and initial phenotypic characterization of MH27-2

The MH27-2 genome comprises a circular chromosome (2,979,490 bp, 52.6% GC) encoding an estimated 2,796 proteins, and a small plasmid (5,962 bp, 50.1% GC) encoding eight proteins. Pathways were identified for the biosynthesis of all amino acids except tryptophan. Two biosynthetic gene clusters were found, predicted to encode a ranthipeptide and a quorum sensing cyclic-lactone-autoinducer, respectively. MH27-2 was predicted and experimentally shown to use a wide range of carbon and nitrogen sources, with the major end products of glucose fermentation being acetate and succinate (**Supplementary Information, Table S2**). Consistent with a lack of genes for oxygen tolerance, the strain is strictly anaerobic, and is able to grow in the presence of bile, albeit with an extended lag phase and reduced growth rate and yield (**Figure S1B**).

### MH27-2 reduces disease severity in acute murine colitis

The therapeutic potential of MH27-2 was first evaluated in a blinded, dextran sulphate sodium (DSS) induced model of acute murine colitis, a well-documented model of UC flares that resembles the pathology of gut epithelial injury and inflammation^43^. Live cultures of MH27-2 or vehicle controls were delivered by oral gavage once daily for seven days, commencing on the fifth day of DSS administration (**Figure 2A**). The glucocorticoid prednisone was used as a positive control, as it is a potent anti-inflammatory agent and mainstay of IBD treatment during flares^44^, with proven efficacy in the selected model^38,45,46^. DSS administration caused significant colitis that was ameliorated by treatment with MH27-2 or prednisone, as evidenced by histological scoring of epithelial damage and inflammation (**Figure 2B-E)**. Treatment with MH27-2 did not affect body weight or endoscopic and biochemical measures of inflammation (**Figure S2A-C**), and only marginally recovered DSS-induced reduction in fecal microbial diversity (**Figure S3**).

**Figure 2.**
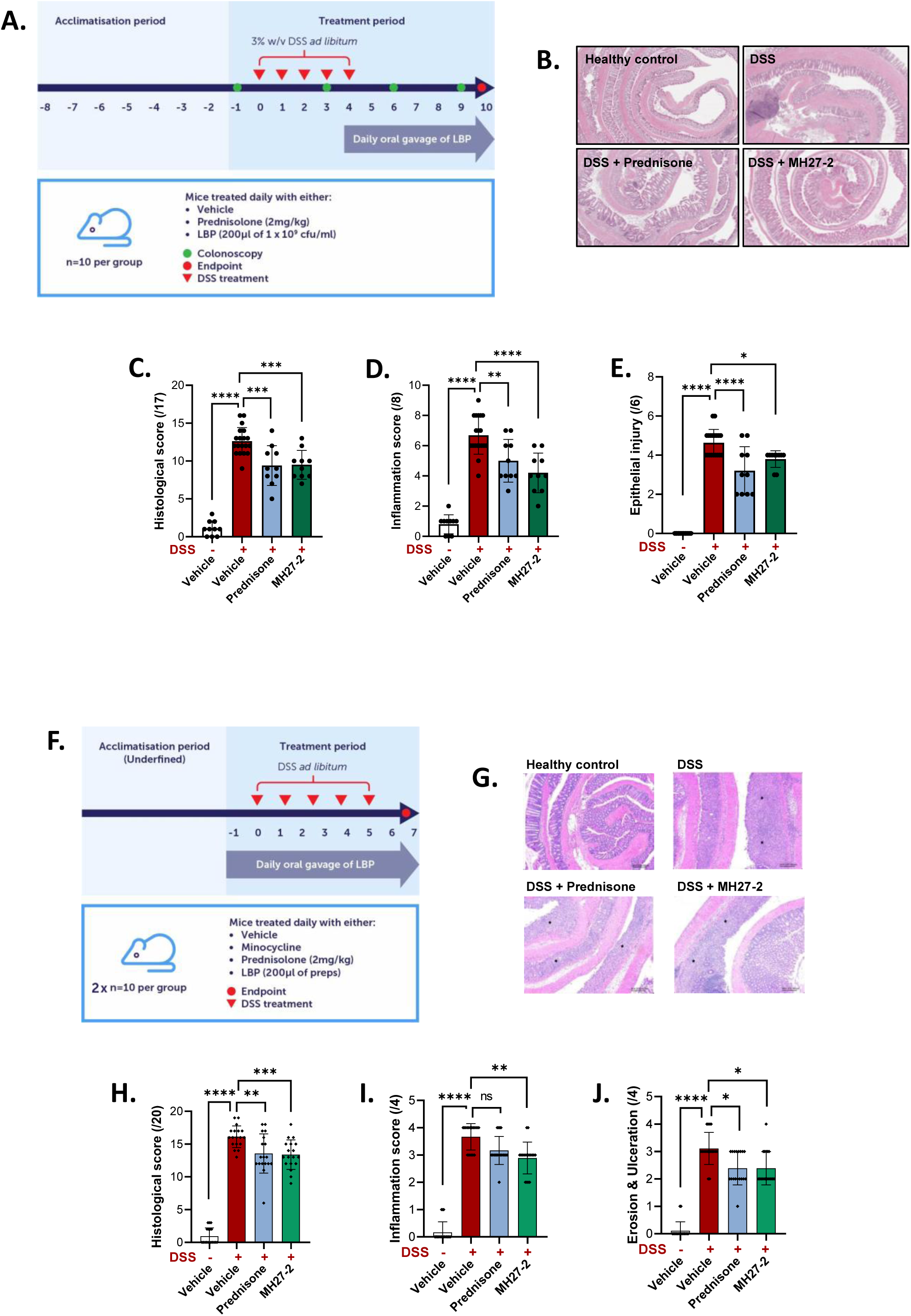
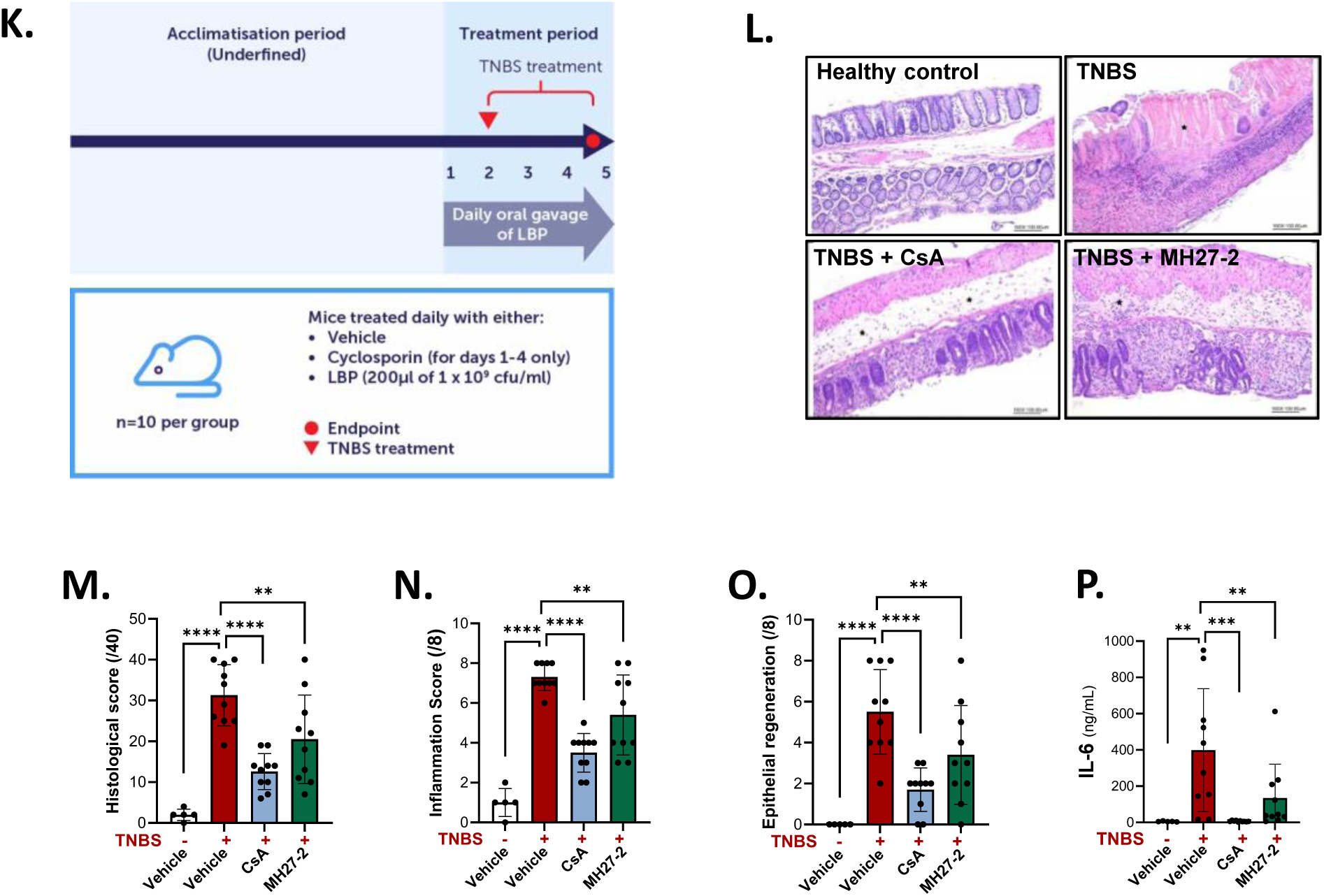
**A**. Live cultures of MH27-2 were assessed in a therapeutic dextran sulphate sodium (DSS) mouse model of colitis (n=10 per experimental group, except DSS control group n=20). Treatment with MH27-2 ameliorated DSS induced colitis as illustrated by representative gut histology images (**B.**) and as evidenced by a reduction of the histopathological score (**C.^$^**), including the sub-score measures of inflammation (**D.^$^**) and epithelial injury (**E.^#^**), compared to the DSS vehicle control. **F.** Live cultures of MH27-2 were assessed in a prophylactic DSS mouse model (n=20 per group). Treatment with MH27-2 ameliorated DSS induced colitis as illustrated by representative gut histology images (**G.**) and as evidenced by a reduction of the histopathological score (**H.^$^**), including the sub-score measures of inflammation (**I.^#^**) as well as erosion and ulceration (**J.^#^**), compared to the DSS vehicle control. **K.** Prophylactic effects of live MH27-2 were assessed in a mouse model of acute murine colitis that uses the haptenizing agent 2,4,6-Trinitrobenzene sulfonic acid (TNBS) to cause predominately Th1-driven pro-inflammatory responses in the murine colon characteristic of CD in humans. MH27-2 ameliorated TNBS induced colitis as illustrated by representative gut histology images (**L.**) and as evidenced by a reduction of the histopathological score (**M.^$^**), including the sub-score measures of inflammation (**N.^$^**) as well as epithelial regeneration (**O.^#^**), and reduced levels of pro-inflammatory cytokine interleukin-6 (IL-6) in colonic tissue (**P.^#^**) compared to the DSS vehicle control. For all data, ns: not significant; *, p < 0.05; **, p < 0.01; ***, p < 0.001; ****, p < 0.0001; presented is the mean and standard deviation. For statistical testing: **^$^**One-way ANOVA with uncorrected Fisher’s LSD test for multiple comparison. **^#^**Kruskal-Wallis test with uncorrected Dunn’s multiple comparisons.

The prophylactic efficacy of MH27-2 was next evaluated by commencing treatment one day prior to DSS administration (**Figure 2F**). Live cultures of MH27-2 were delivered daily by oral gavage for nine days. As animal housing can impact disease pathogenesis^47^, these experiments were conducted in an independent animal house at Eurofins (Taiwan). Prednisone was less efficacious in the prophylactic DSS model (**Figures 2H-J**), where innate immune responses play a prominent role, and the model had a shorter timeframe. This outcome was expected as corticosteroids principally work by inhibiting the activation and function of T cells and are therefore more effective in the therapeutic DSS model where adaptive immunity plays the more prominent role. Based on the histological assessment, and consistent with the therapeutic DSS experiments, MH27-2 treatment showed comparable beneficial effects to the prednisone control (**Figure 2G-J, Figure S4**).

Prophylactic effects of MH27-2 were also assessed in a second model of acute murine colitis using the haptenizing agent 2,4,6-Trinitrobenzene sulfonic acid (TNBS) (**Figure 2K, Figure S5**). TNBS causes a strong immune cell-mediated response in the colon largely driven by Th1 inflammation that culminates in deep tissue inflammation affecting all layers of the gut wall (transmural) characteristic of CD in humans^48^. Live MH27-2 was delivered daily by oral gavage for five days commencing one day prior to TNBS administration (**Figure 2K**). Cyclosporine A, a potent immunosuppressive peptide commonly utilized as a rescue treatment for IBD^49,50^ and with proven efficacy in the TNBS murine model^51^, was administered as a positive control. MH27-2 significantly ameliorated TNBS-induced histological damage in the colon by increasing epithelial regeneration, and reducing inflammation, erosion, ulceration, and abnormalities in mucosal architecture (**Figure 2L-O**, **Figure S5**). MH27-2 also significantly reduced protein levels of the IBD-associated pro-inflammatory cytokine interleukin-6 (IL-6) (**Figure 2P**), but not IL-12, IL-17, TNF or the neutrophil inflammatory marker myeloperoxidase, in colonic tissue (**Figure S5**).

In summary, MH27-2 effectively promotes murine wound healing and gut barrier integrity; and suppresses disease-relevant inflammatory responses such as IL-6 expression. Protective effects of MH27-2 were both therapeutic and prophylactic, demonstrated independently in two mouse models and in different animal houses.

### MH27-2 promotes mucosal healing

Mucosal healing, the repair of the intestinal lining with visible resolution of inflammation, has three distinct phases – *restitution*, characterized by migration of epithelial cells into the damaged mucosa, *proliferation* of epithelial cells to repair wounded areas, and *restoration* of gut barrier integrity^52^. The effects of MH27-2 on all three phases were investigated using human cell-based assays and mouse intestinal organoids. We used MH27-2 culture supernatant concentrated for non-polar metabolites (metabolite extract), as they are an important source of bioactives^53,54^. This MH27-2 metabolite extract significantly promoted epithelial cell migration (**Figure 3A-B**) and proliferation (**Figure S6A-C**), and accelerated wound repair in a scratch assay using IFNγ-stimulated gut epithelial cells to model an inflamed gut environment (**Figure 3C-D**).

**Figure 3.**
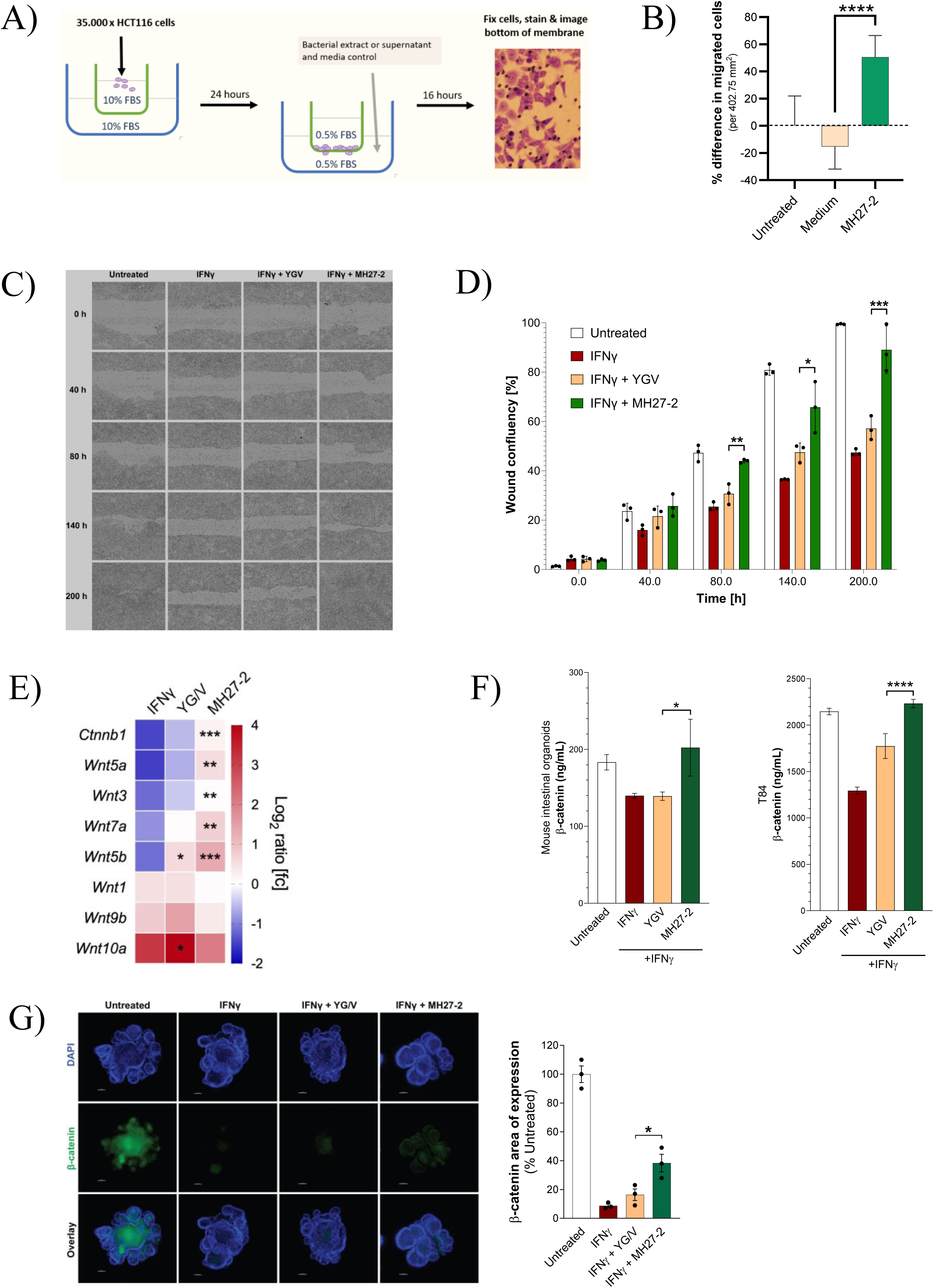
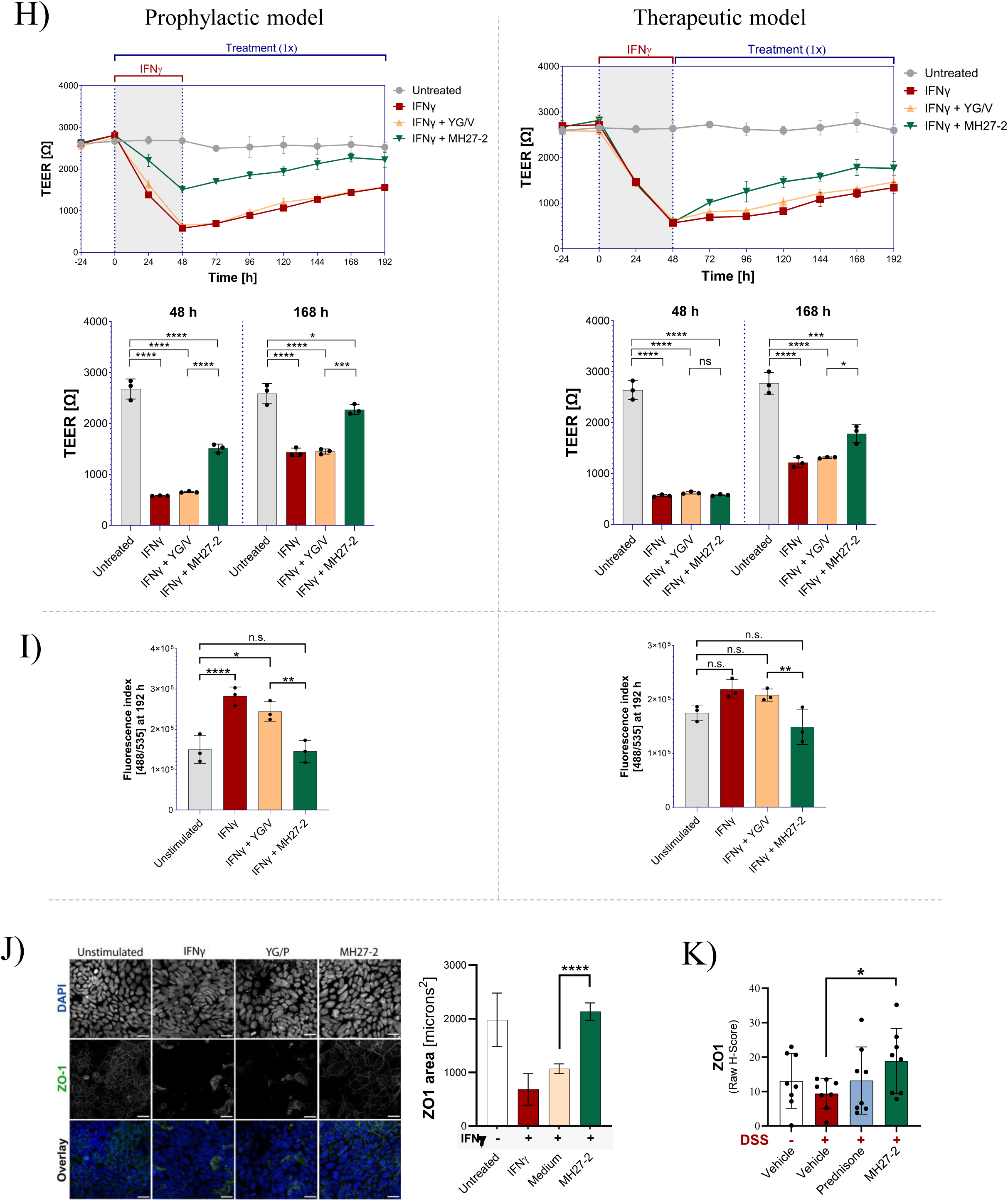
**A**. Schematic representation of Transwell cell migration. **B.** Compared to the medium control, MH27-2 extract induced faster migration of HCT116 gut epithelial cells. Unpaired t-test; biological duplicates, four technical replicates each. **C.** The impact of MH27-2 on intestinal epithelial cell migration was also assessed using an *in vitro* scratch wound assay. The rate of wound closure of T84 gut epithelial cells challenged with IFNγ was monitored for up to 200 hours post-scratch. MH27-2 extract (1X) promoted wound closure as evidenced by representative images of biological triplicates. **D.** Quantitation of wound confluency of the experiment presented in Figure 3C. T84 cells showed a significantly accelerated rate of wound closure in the presence of MH27-2 secretome extract 80, 140 and 200 hours after wounding, compared to the control cells treated with bacterial medium extract control. **E.** In mouse intestinal organoids, MH27-2 metabolite extract (1X) significantly mitigates IFNγ (100 ng/mL) mediated transcriptional changes of β-catenin (*Ctnnb1*) and multiple WNT ligands after 24 hours. Experiment performed in biological triplicates, two technical replicates each; paired t-test comparing IFNγ + YGV medium control to IFNγ + MH27-2. **F.** At the protein level, as assessed by ELISA, suppression of β-catenin by IFNγ is ameliorated by treatment with MH27-2 extract (1X) after 24 hours in mouse intestinal organoids and T84 gut epithelial cells. Biological triplicates, technical duplicates; unpaired t-test comparing IFNγ + YGV medium control to IFNγ + MH27-2. **G**. β-catenin protein expression in mouse intestinal organoids as assessed by immunofluorescence straining and confocal microscopy. Representative images of three independent biological replicates. Image quantitation is presented to the right (the average ratio of β-catenin area of expression relative to the area of expression of DAPI (µm^2^) and normalized to the untreated organoids). **H.** The effect of MH27-2 on barrier function of T84 gut epithelial cells was assessed in two trans-epithelial electrical resistance (TEER) models. In both models, IFNγ was used as a barrier disruptor which, after 48 hours, led to an approximately 80% reduction in TEER, indicative of an increased barrier permeability, which naturally and gradually recovered after removing of the stimulus. The prophylactic model is presented to the left, where T84 cells were treated with MH27-2 extract (1X) for one hour prior to challenge with IFNγ, and MH27-2 was replenished every 24 hours throughout the duration of the experiment. In the prophylactic model, to the right, T84 cells received MH27-2 treatment only after barrier disruption occurred in response to IFNγ. In both models, MH27-2 promoted recovery compared to the medium control extract, and prophylactic treatment was more effective than therapeutic administration. One-way ANOVA at 48 and 168 hours. **I.** At experimental endpoint of the TEER assays presented in Figure 3H, cell permeability was assessed by paracellular translocation of FITC-labelled dextran. Compared to the medium control, MH27-2 extract significantly reduced the flux of FITC-dextran across the T84 cell monolayer. One-way ANOVA. For all bar graphs presented in Figure 3, data is presented as the mean and SD, and ns: not significant; *, p < 0.05; **, p < 0.01; ***, p < 0.001; ****, p < 0.0001.

At a molecular level, the WNT/β-catenin signaling pathway plays an important role in cell proliferation and barrier restitution, and dysregulation of WNT ligands and associated genes has been observed in IBD^55,56^. In murine intestinal organoids, MH27-2 metabolite extract reversed IFNγ-induced transcriptional changes in expression of select WNT ligands and the WNT transcriptional regulator β-catenin (**Figure 3E**). The metabolite extract also mitigated IFNγ-mediated loss of β-catenin at a protein level in both intestinal organoids and human gut epithelial cells (**Figure 3F-G**). This is important, as β-catenin is a dual function protein that promotes the proliferation and differentiation of epithelial cells through β-catenin dependent WNT signaling, and also links epithelial cells together to form an impermeable barrier.

We next investigated the effect of MH27-2 on restoration of gut barrier integrity by challenging epithelial cells with IFNγ or IL-6, both well-characterized barrier disruptors that are highly upregulated in IBD and known drivers of the disease^57,58^. Prophylactic, and to a lesser degree therapeutic, administration of MH27-2 metabolite extract ameliorated IFNγ-induced loss of barrier integrity in a T84 cell monolayer as evidenced by trans-epithelial electrical resistance (TEER) (**Figure 3H**) and translocation of fluorescently labelled dextran across the epithelial layer (**Figure 3I**). Similar observations were made when using IL-6 as a barrier disruptor (**Figure S7A-B**). These observed protective effects on gut barrier integrity can, in part, be explained by transcriptional and translational changes in the tight junction protein ZO-1, which plays a key role in barrier function by connecting epithelial cells to form a continuous barrier between the lumen and underlying tissues of the gut^59,60,61^. MH27-2 metabolite extract restored ZO-1 protein expression in T84 cells (**Figure 3J**) and ameliorated IFNγ-induced transcriptional changes in *ZO1* in murine intestinal organoids (**Figure S7C**). Consistent with these *in vitro* results, administration of live MH27-2 cells enhanced colonic ZO-1 protein expression in mice with DSS-induced colitis (**Figure 3K**). MH27-2 may also indirectly protect gut barrier integrity by suppressing immune cell infiltration, which can compromise gut barrier function through the production of pro-inflammatory cytokines that downregulate tight junction protein expression, induce epithelial cell death and reduce mucus production^62^. MH27-2 metabolite extract ameliorated IFNγ-induced transcription of *IL6* in murine intestinal organoids (**Figure S7C**), and reduced IFNγ-induced transcription of *CCL5* and *CXCL10* in T84 human epithelial cells (**Figure S7C**). This is consistent with reduced immune cell infiltration observed *in vivo* (**Figures 2D, I, N**).

The direct effects of IFNγ and IL-6 (classical and trans) signaling on gut epithelial cells are mediated through JAK-STAT signaling, a pathway central to epithelial integrity and mucosal immunity^63^. We therefore evaluated the ability of MH27-2 to affect these pathways using reporter cell assays, revealing suppression of both STAT1 and STAT3 signaling activation (**Figure S7D-F, Supplementary Information**). Consistent with these findings, MH27-2 metabolite extract also ameliorated IFNγ-mediated induction of STAT1 regulated genes in T84 cells and intestinal organoids (**Figure S7C**). Together, these results suggest that MH27-2 mitigates inflammation-associated effects on barrier integrity and promotes mucosal healing.

### MH27-2 produces aromatic lactic acids that support barrier integrity

To identify bioactives secreted by MH27-2 that support gut barrier integrity, cell-free supernatant was fractionated, and the effect of individual fractions on ZO-1 expression examined in T84 gut epithelial cells. Of 36 fractions assessed, 11 ameliorated IFNγ-induced reduction of ZO-1 protein expression (**Figure S8A**), and five of these were enriched for aromatic lactic acids, including indole-3-lactic acid (ILA; >200,000-fold increase) and phenyllactic acid (PLA; >1,200-fold increase). Aromatic lactic acids are important extrinsic microbial regulators of mucosal immunity and gut barrier integrity and have previously been recognized to hold therapeutic potential in IBD^64,65^.

Genome-based metabolic reconstruction and *in vitro* testing confirmed that MH27-2 produces high quantities of ILA, HPLA (4-hydroxyphenyllactic acid) and the L-isomer of PLA (L-PLA) when supplemented with the respective aromatic pyruvic acids (**Table S3, Figure S8B, Supplementary Information**). A small amount of the D-isomer of PLA (D-PLA) was also produced constitutively (**Figure S8B**). Heterologous gene expression revealed that aromatic lactic acid production by MH27-2 was driven by a specific D-lactate dehydrogenase (**Table S4, Supplementary Information**). All four aromatic lactic acids ameliorated IFNγ-induced reduction of ZO-1 protein expression in T84 cells (**Figure S8C-D**), but only HPLA enhanced *ZO-1* expression at a transcriptional level (**Figure S8E**). Further, treatment of T84 cells with ILA, D-PLA, and L-PLA suppressed IFNγ-induced expression of one or multiple of the pro-inflammatory cytokine genes *CCL3*, *IL6*, or *ISG15* (**Figure S8E)**, while HPLA did not affect expression of these genes. ILA was further examined in a TEER assay, demonstrating its ability to mitigate IFNγ-induced loss of gut barrier integrity in a dose dependent manner (**Figure S8F)**.

Interestingly, multiple fractions that lacked detectable aromatic lactic acids also induced ZO-1 expression (**Figure S8A**), supporting the conclusion that MH27-2 produces multiple classes of molecularly distinct bioactives that collectively modulate gut barrier integrity. Together, these results suggest that the aromatic lactic acids and other unidentified bioactives secreted by MH27-2 contribute to its ability to restore gut barrier integrity and promote immune homeostasis. The potential of MH27-2 to be metabolically active *in situ* means that these functionalities could be exerted directly at the site of injury in the gut.

### MH27-2 promotes immune homeostasis

We next used human cell-based assays to explore the potential of MH27-2 to re-establish immune homeostasis – the delicate balance between effective immune response to pathogens while maintaining tolerance to commensal bacteria, food, and self-antigens. In IBD, differentiation of naïve T-cells is critical to the observed immune imbalance^1,66^. This process is regulated in part by dendritic cells that can either promote tolerance or drive inflammation, by inducing regulatory T-cells (Tregs), or helper T-cells (e.g. Th1, Th17, Th2), respectively. Immature monocyte-derived dendritic cells were incubated with MH27-2, then co-cultured with naïve CD4^+^ T cells, leading to a significant increase in Treg differentiation (**Figure 4A**). Exposure to MH27-2 did not increase secretion of the anti-inflammatory cytokine IL-10 in dendritic cells, indicating that Treg differentiation was stimulated in an IL-10-independent manner (**Figure 4B**). Our data suggests that MH27-2’s ability to induce Tregs is driven directly by bacterial cells and/or cell components and is independent of aromatic lactic acid production.

**Figure 4.**
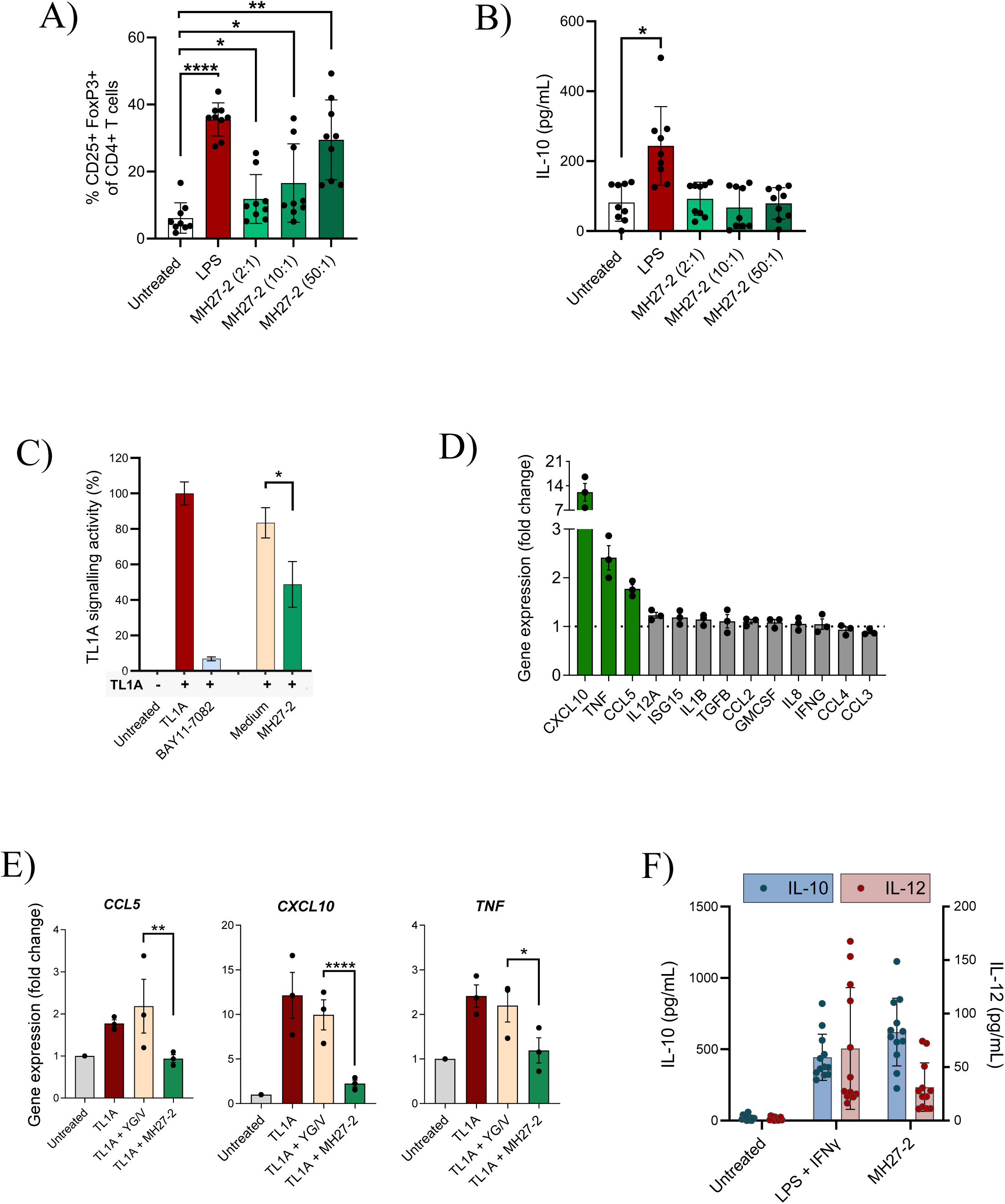
**A**. Immature monocyte-derived dendritic cells were exposed to MH27-2 bacterial cells, followed by co-culture with naïve CD4+ T cells, resulting in differentiation to Foxp3^+^ regulatory T cells. **B.** MH27-2 did not induce secretion of the anti-inflammatory cytokine IL-10 by dendritic cells, indicating that Treg differentiation was probably IL-10-independent. **C.** In TL1A reporter cells challenged with recombinant human TL1A (100 ng/mL), MH27-2 15% culture supernatant significantly mitigates TL1A signaling activation, compared to the bacterial medium control; Data presents mean ± SD from 3 biological replicates, 3 technical replicates each; unpaired t-test *p* = 0.025. **D.** Relative gene expression of CCL5, CXCL10, and TNF in DR3-expressing TF-1 cells challenged with TL1A is significantly reduced in cells treated with MH27-2 1x extract, compared to the medium extract. 3 biological replicates, 2 technical replicates; mean ± SD, unpaired t-test. **E.** Co-incubating MH27-2 bacterial cells with human PBMCs, resulted in PBMCs secreting higher levels of anti-inflammatory IL-10 than pro-inflammatory IL-12.

In IBD, dendritic cells upregulate TNF-like cytokine 1A (TL1A), thereby disrupting Treg polarization and favoring the differentiation of Th1 and Th17 cells^67^. Consequently, blocking TL1A signaling is a promising new therapeutic strategy to restore immune homeostasis^68^. MH27-2 culture supernatant significantly inhibited TL1A signaling in a reporter cell line (**Figure 4C**). We next examined the ability of MH27-2 culture supernatant to modulate TL1A regulated gene expression in the TF-1 erythroblast cell line as it is known to express the TLA1 receptor, DR3^69^. Analysis of 13 pro-inflammatory genes revealed that TL1A treatment strongly induced three genes (*CCL5*, *CXCL10*, and *TNF*), encoding disease relevant cytokines and chemokines (**Figure 4D**). These TL1A-induced transcriptional changes were significantly mitigated by MH27-2 metabolite extract (**Figure 4E**).

Unlike with dendritic cells, MH27-2 strongly induced the anti-inflammatory IL-10 in PBMCs relative to low levels of the pro-inflammatory IL-12 (**Figure 4F**), resulting in an IL-10/IL-12 ratio of 20. A high IL-10/IL-12 ratio has previously been shown to correlate with anti-inflammatory effects *in vivo* and the promotion of tolerogenic immune responses^70,71^.

Collectively, our results demonstrate the potential of MH27-2 to promote immune homeostasis, through induction of Tregs and IL-10, and suppression of TL1A signaling in disease-relevant immune cells.

### Safety assessment and drug product manufacture

To support development of MH27-2 as an LBP, its safety was assessed using a combination of *in silico* and laboratory-based methods. Analysis of the closed MH27-2 genome did not reveal the presence of any known bile salt hydrolases, or ability to produce biogenic amines (**Table S5**). Although genes associated with resistance against tetracyclines (*tet(W)*), aminoglycosides (*ant(6)*), and macrolides (ABC-F type ribosomal protection protein) were present, *in vitro* testing demonstrated that MH27-2 was sensitive to clinically relevant antibiotics including amoxicillin, clindamycin, vancomycin and metronidazole (**Table S5**). Genome screening did not identify any relevant virulence factors, with the only hits being commonly found in gut bacteria, including proteins that may carry out functions relevant to colonization and persistence (**Table S5, Supplementary Information**). No intact prophages were detected *in silico* and no virulent or inducible phages were detected *in vitro* (**Figure S9A**). Consistent with preferred manufacturing practices, MH27-2 could be grown with animal component-free media, with a growth rate and yield suitable for manufacturing (**Figure S9B**).

With no immediate barriers to drug substance production identified, a current Good Manufacturing Practice (cGMP) production process for MH27-2 was developed alongside Bacthera A/S (Denmark). A characterized master cell bank was established as the starting material for fermentation, and animal component-free media and production conditions were optimized to support clinical scale 100 L production under anaerobic conditions. The production process consisted of fermentation, harvest of bulk bacteria and concentration by cross flow filtration, mixing with proprietary cryopreservative, and lyophilization (freeze drying). For clinical administration, the resulting bulk drug substance (**Figure S10A**), named MAP 315, was further formulated with proprietary excipients and encapsulated using enteric capsules at Bacthera AG (Switzerland) to produce the MAP 315 drug product (**Figure S10B**). Characterization and release testing of the cGMP material was conducted in accordance with regulatory requirements^72^ and supported the viability of the material for clinical evaluation.

The therapeutic efficacy of the MAP 315 drug substance was assessed in a prophylactic model of DSS-induced murine colitis (**Figure S11A**). Prednisone treatment resulted in a small reduction in histopathological score (**Figure S11B**) consistent with its modest efficacy in the prophylactic DSS model. In contrast, both the MAP 315 drug substance and MH27-2 prepared using our standard protocol (MH27-2 (live)) significantly ameliorated disease activity and improved the histopathological score (**Figure S11B**), including reduced inflammation (**Figure S11C**), reduced erosion and ulceration (**Figure S11D**), and lower percentage of the colonic epithelium affected (**Figure S11E**). Importantly, there was no significant difference in the efficacy of the MAP 315 drug substance and MH27-2 (live), indicating that the drug substance manufacturing process did not impact therapeutic efficacy (**Figures S11B-G**, *p* >0.05 for all measures except epithelial regeneration where MAP 315 (5E4) is significantly different to MH27-2 (live) (**Figure S11B**, *p* <0.05).

### Tolerability in mice and Phase 1 clinical study in healthy volunteers

Tolerability of the MAP 315 drug substance was evaluated in healthy adult C57BL/6 mice. For this, MAP 315 was administered orally for 14 days, at three doses of up to 1×10^8^ live cells/mouse/day. No clinically relevant effects on clinical signs, hematology, blood chemistry, or histopathology of the gastrointestinal tract, spleen, and mesenteric lymph nodes, in response to MAP 315 were noted (**Figure S12**). Further, no significant translocation to the serum, spleen, thymus, mesenteric lymph nodes, liver, kidneys, heart, lungs, brain, ovaries, or testis, was observed using strain-specific qPCR. Overall, the study demonstrated that the drug substance is well tolerated in healthy mice.

Next, a Phase 1 clinical study was conducted to evaluate the safety and tolerability of MAP 315 drug product in humans (**Figure S13**). The study was a single-center, randomized, double-blind, placebo-controlled, multiple dose study in 32 healthy adults. Participants were randomized 3:1 to receive MAP 315 or a matching placebo for 14 consecutive days, and divided into low-dose (one placebo or MAP 315 (daily dose of 4 x 10^7^ MAP 315 colony forming units (CFU)) capsule daily), and high-dose (eight placebo or MAP 315 (daily dose of 3.2 x 10^8^ MAP 315 CFU) or placebo capsules daily) cohorts. All participants completed the study as scheduled, and most received all doses of their assigned study treatment (MAP 315 98.7% mean compliance; placebo 100% mean compliance). No serious adverse events occurred, and any reported adverse events were mild, with none-ongoing at the end of the study (**Table 1)**. There were no clinically significant changes in vital signs, electrocardiograms, physical examinations, or safety laboratory findings. No bacterial translocation of MAP 315 to plasma was detected, and there were no clinically significant changes in inflammatory markers (i.e., plasma C-reactive protein, or fecal calprotectin).

**Table 1:** Incidence of Treatment-Emergent Adverse Events (TEAEs) in Phase 1 Study of MAP 315.

| System Organ Class (SOC)<br>Preferred Term (PT) | Low dose MAP 315<br>1 capsule per day<br>(N=12)<br>n (%) E | High dose MAP 315<br>8 capsules per day<br>(N=12)<br>n (%) E | Pooled<br>Placebo<br>(N=8)<br>n (%) E |
| --- | --- | --- | --- |
| At Least One Possibly<br>Related TEAE | 2 (16.7) 3 | 3 (25.0) 3 | 2 (25.0) 2 |
| Gastrointestinal disorders | 1 (8.3) 1 | 2 (16.7) 2 | 1 (12.5) 1 |
| Abdominal discomfort | 0 | 1 (8.3) 1 | 0 |
| Dry mouth | 0 | 1 (8.3) 1 | 0 |
| Nausea | 1 (8.3) 1 | 0 | 0 |
| Vomiting | 0 | 0 | 1 (12.5) 1 |
| Nervous system disorders | 2 (16.7) 2 | 0 | 1 (12.5) 1 |
| Headache | 2 (16.7) 2 | 0 | 1 (12.5) 1 |
| Respiratory, thoracic and<br>mediastinal disorders | 0 | 1 (8.3) 1 | 0 |
| Rhinorrhoea | 0 | 1 (8.3) 1 | 0 |

All adverse events were mild. Participants who experienced multiple events within a category are counted only once in the specific category (n), however each instance of the event is counted (E). Percentage (%) of participants (n) are calculated based on the number of participants in the relevant analysis set (N).

## Discussion

In this study, we present the isolation and characterization of *H. mulieris* MH27-2 as a novel LBP drug candidate for ulcerative colitis, the subsequent development of cGMP manufactured MAP 315 drug product, and its evaluation in a Phase 1 clinical trial. MAP 315 was discovered using a metagenome-guided, large cohort-based approach^38^, and developed from a novel strain of *H. mulieris* isolated from a healthy human donor. Existing IBD drugs primarily function by directly targeting effector immune cells or their effector cytokines thereby suppressing the host inflammatory response. However, these drugs suffer from toxicity, poor patient compliance, and there are currently no approved IBD drugs that directly target mucosal healing and immune homeostasis. In preclinical models, MH27-2 promotes mucosal healing, improves gut barrier integrity, promotes anti-inflammatory effects, and induces immune homeostasis (**Figure 5**). It has excellent safety characteristics and was well tolerated in a Phase 1 human clinical trial. Based on its efficacy and safety profile, we therefore propose that MAP 315 could benefit patients with mild-to-moderate UC, including patients who are not adequately responding to 5-ASAs or are on other therapeutics where mucosal healing has not been achieved. The multi-modal mechanism of action of MAP 315 may overcome the limitations of many current therapies that have single immune targets, thereby improving initial response rates, prevent secondary loss of response, and increase length of remission.

**Figure 5.**
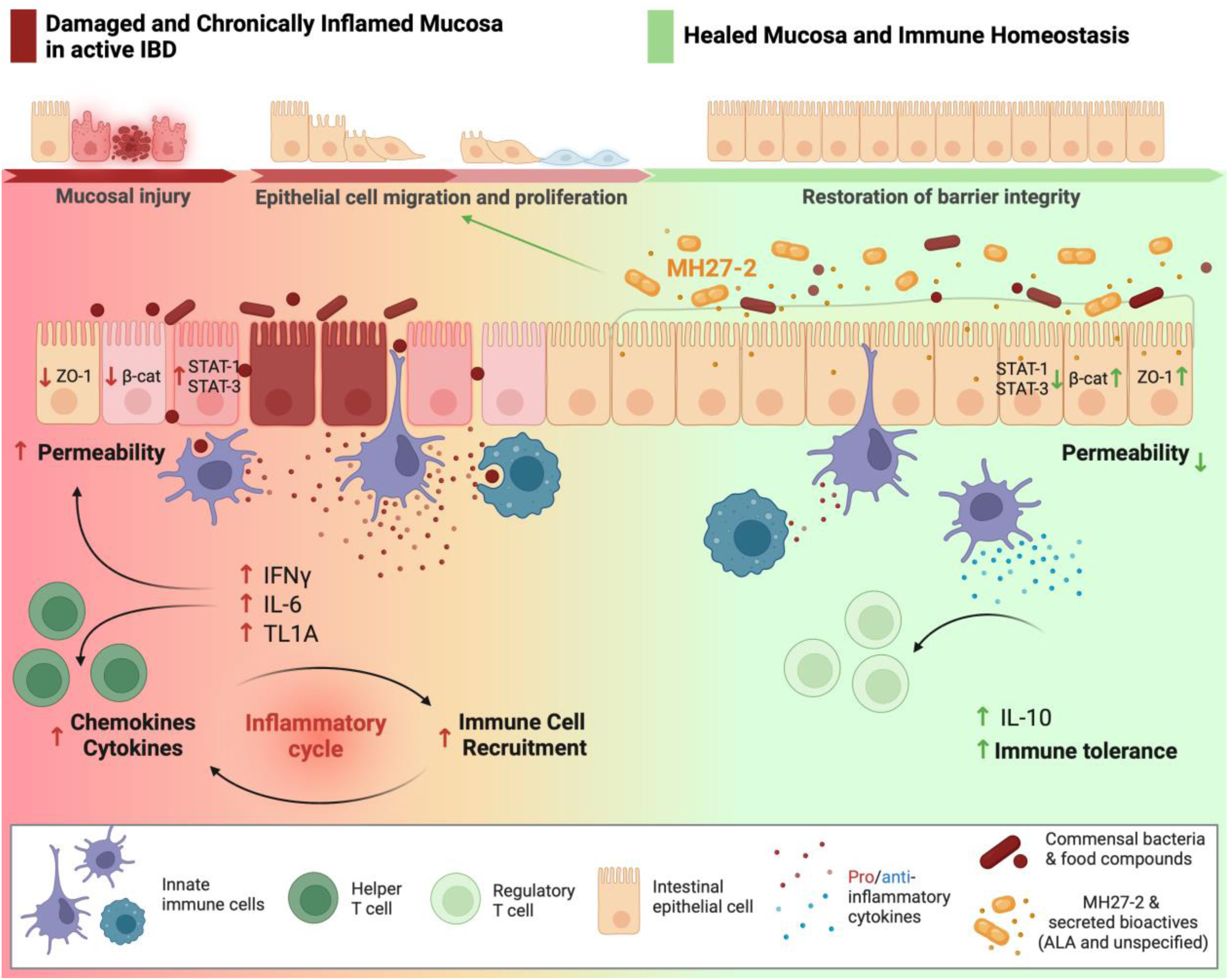
Schematic summarizing the proposed therapeutic effects and mode of action of MH27-2 in the context of IBD. Wound healing in the gut involves the migration of epithelial cells to cover the injury site, followed by their proliferation to restore the integrity and functionality of the intestinal barrier. MH27-2 promotes both the migration and proliferation of injured gut epithelial cells. In IBD, gut barrier function is compromised due to disrupted epithelial integrity, increased permeability, and impaired tight junctions, leading to heightened immune activation and chronic inflammation. MH27-2, secretes both known (ALAs = aromatic lactic acids) and unknown bioactives, that influence several of these processes to promote restoration of gut barrier integrity and immune homeostasis.

We evaluated the efficacy of MH27-2 against two immunosuppressants, prednisone and cyclosporine, which have been used in clinical practice for treating severe IBD flares^7^. In murine models of acute colitis, MH27-2 displayed comparable efficacy to that of prednisone, but as expected, was less effective than cyclosporine, a highly potent immunosuppressant that often shows strong side effects and is used as a rescue therapy for severe IBD. MAP 315 showed no adverse effects in either mice or humans. In a tolerability study with healthy mice, MAP 315 drug substance was well tolerated at doses up to 1 x 10^8^ CFU/day. On a CFU/kg basis, this equates to a human clinical dose of ∼2.5 x 10^11^ CFU/day providing a ∼25-fold safety margin over a typical clinical starting dose of 1 x 10^10^ CFU. Based on these results and given that *H. mulieris* is a natural component of the healthy human gut microbiome, we expect that MAP 315 will be a safe and effective long-term treatment for IBD, an important advantage over existing IBD drugs.

The gut microbiome is now widely recognized as a key risk factor underpinning the pathogenesis of IBD. This has spurred research in “restoration ecology” approaches to reconstitute a healthy gut microbiome. Among these approaches, fecal microbiota transplants (FMT) have shown efficacy for treating IBD, particularly UC, highlighting the potential of gut microbiome derived drugs. However, FMTs have limitations, including variable donor microbiota, safety concerns, the complexity of treatment regimes, or inability of the transferred microbes to colonize the inflamed gut^73,74^. This underscores the need for more precise, controlled and reproducible live bacterial therapies, which has driven interest in developing microbial consortium based drugs^75–77^. While these consortium-based therapies show promise, advances in high-resolution microbial profiling now enable identification of specific bacterial strains, with potentially disease modulating activities. This supports the development of defined microbial drugs, as an alternative to microbiome replacement therapies. Single-strain therapeutics offer notable advantages over the significant challenges associated with commercial-scale production of microbial consortia^78^, including simplified manufacturing and quality control, improved stability and safety, and simplified regulatory and commercial complexity^79,80^. Using mouse models of colitis and ileitis we previously demonstrated that rationally selected single strain LBP candidates exhibited therapeutic effects comparable to a consortium containing these same strains, indicating no additional benefit from the combination therapy^38^. Consistent with this observation, numerous independent studies have demonstrated the efficacy of single strains in animal models of IBD (e.g. *Faecalibacterium prausnitzii*^70^, *Bacteroides thetaiotaomicron*^81^, *Bacteroides ovatus*^82^), with *F. prausnitzii* and *B. thetaiotaomicron* already being evaluated for the treatment of CD in human studies^83,84^. *H. mulieris* MH27-2 is a promising new addition to these single strain LBPs. As a clear difference to *F. prausnitzii*, MH27-2 promotes gut barrier function, as measured by the TEER and ZO1 assays, whereas *F. prausnitzii* does not show any activity in these assays^38,85–89^. However, several open questions remain.

First and foremost, the clinical efficacy of MAP 315 in UC patients is yet to be determined, although current data from animal studies and human cell-based assays are highly promising, and safety has been established in healthy volunteers. Moreover, it is unknown whether MAP 315 will be equally effective across diverse IBD patient populations, including Crohn’s disease, considering variations in microbiome composition, host genetics, and dietary patterns, or in other conditions with clinically relevant gut inflammation (e.g. graft versus host disease, immune checkpoint inhibitor induced colitis). Whether the strain can successfully engraft in the diseased gut also needs to be assessed, however, engraftment is likely not required for therapeutic efficacy of MAP 315 given the rapid onset of action observed in the animal models. The clinically relevant bioactive compounds of MH27-2 and their modes of action remain to be determined with the exception of aromatic lactic acids^65,90–92^, which we found can mitigate IFNγ and IL-6-induced reductions in TEER and ZO-1 expression. Fractionation of the MH27-2 culture supernatant revealed that multiple independent fractions, lacking the aromatic lactic acids, modulated ZO-1 expression suggesting it produces a diverse array of low molecular weight bioactive compounds. From an ecological perspective, the highly redundant ability of MH27-2 to support gut barrier integrity suggests that this is a critical function for the bacterium. These bioactives may exert additive or synergistic effects and attempts to recapitulate MAP 315 clinical efficacy with a specific bioactive could result in reduced efficacy. Understanding which bacterial compounds confer beneficial activities, along with their mechanism of action, may allow the development of new small molecule-based drugs for the treatment of IBD.

In summary, we present a promising new drug candidate for the treatment of IBD that is well tolerated, orally delivered, and addresses the important clinical gaps of mucosal healing and immune tolerance. Industrial scale anaerobic manufacturing processes are in place and the candidate is ready to be progressed to a Phase 2 clinical trial. Our results highlight the value of a data-driven approach to identify safe and effective human microbiome derived therapeutics.

## Data Availability

All data produced in the present study are available upon reasonable request to the authors

## Acknowledgements

We thank Quinn Cao for assisting with the graphical design of the figures. We thank Thomas Kryza, Mark Scott and Cameron Flegg for assistance with the IncuCyte system, and Mark Scott and Cameron Flegg also for assistance with the spinning disc confocal microscope.

## Author contributions

LK, PÓC, PH, GWT conceived the original idea, and PÓC, TM and LK planned the experiments and supervised the project. NA, DLAW, LR and BW supervised aspects of the project. LK identified lead species and JB, CV, JJ, JZ, ER and PÓC did the isolation and characterization. AK, JKL, MN, AR, JZ and MB assessed therapeutic potential. SC, HMC and JS performed animal experiments under the supervision of SK. HC provided regulatory guidance. LF, SMD, NA, JB, RN, DLAW and LK did the sequencing and bioinformatics analysis. JB, MN and LK did the data analysis, machine learning and associated statistics. SK, JB and IHF provided clinical insight. All authors contributed to the interpretation of the results. LK, PÓC, PH, AK, JKL and JB drafted the manuscript. All authors read and approved the final version of the manuscript.

**Supplementary Table 1:** Association with inflammatory conditions.

| Condition | P (Fisher's exact<br>test) | FDR | Prevalence (%) |
| --- | --- | --- | --- |
| Healthy |  |  | 82.1 |
| Inflammatory bowel disease | $8.1 \times 10^{-16}$ | $2.99 \times 10^{-13}$ | 42.6 |
| Crohn's disease | $1.1 \times 10^{-8}$ | $7.47 \times 10^{-7}$ | 39.5 |
| Ulcerative colitis | $1.6 \times 10^{-11}$ | $2.36 \times 10^{-9}$ | 43.9 |
| Non-alcohol fatty liver<br>disease | 0.00071 | 0.0126 | 57.5 |
| Rheumatoid arthritis | $2.0 \times 10^{-4}$ | 0.00719 | 60.9 |
| Asthma | 0.00037 | 0.00243 | 68.3 |
| Fatty liver | 0.019 | 0.0904 | 68.7 |
| Systemic autoimmune<br>disease | 0.007 | 0.0319 | 70.2 |

The prevalence of *H. mulieris* in various disease cohorts compared to a healthy cohort. Statistical significance was assessed using the Fisher's Exact Test, with False Discovery Rate (FDR) corrected p-values reported to account for multiple testing.

**Supplementary Table 2:**
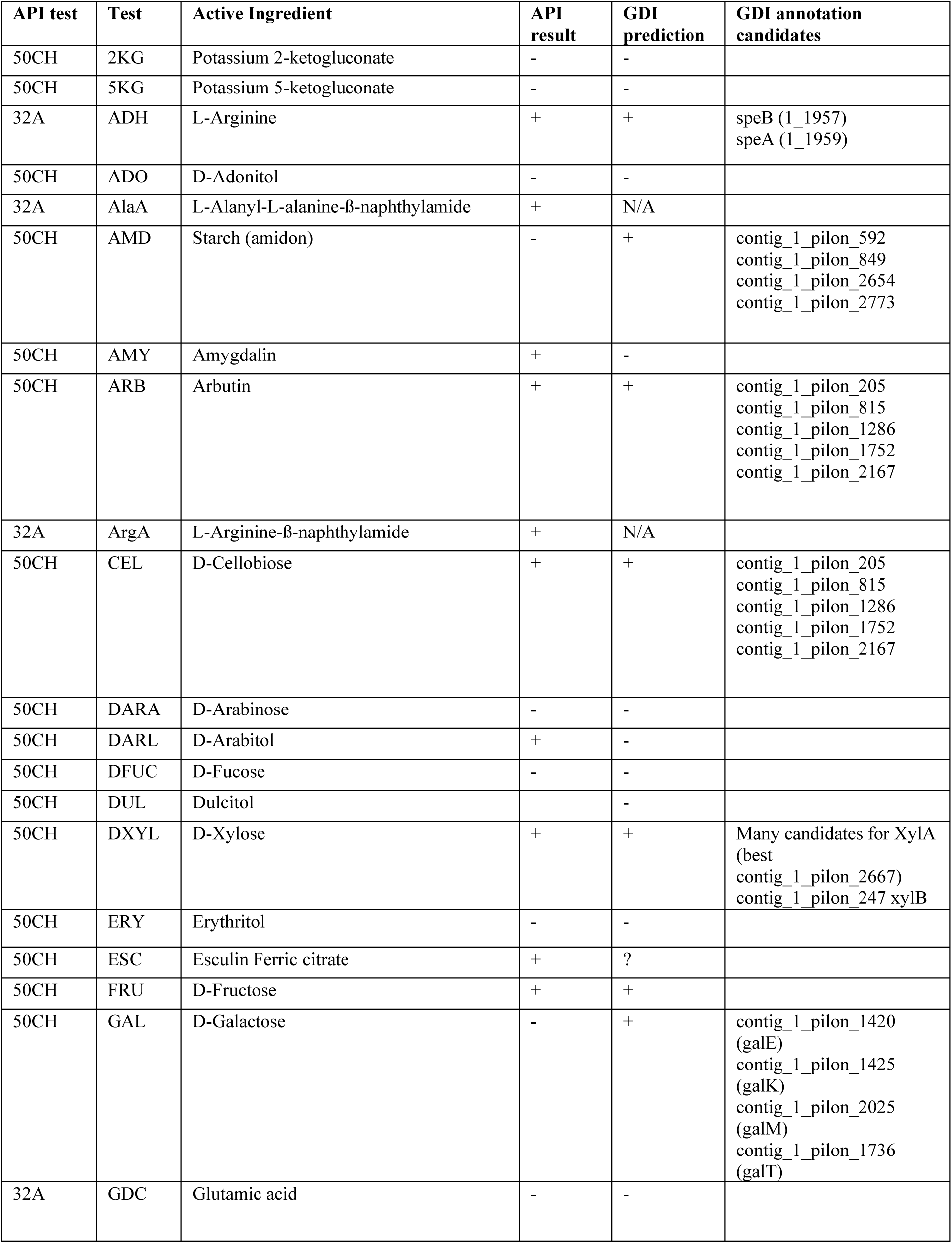
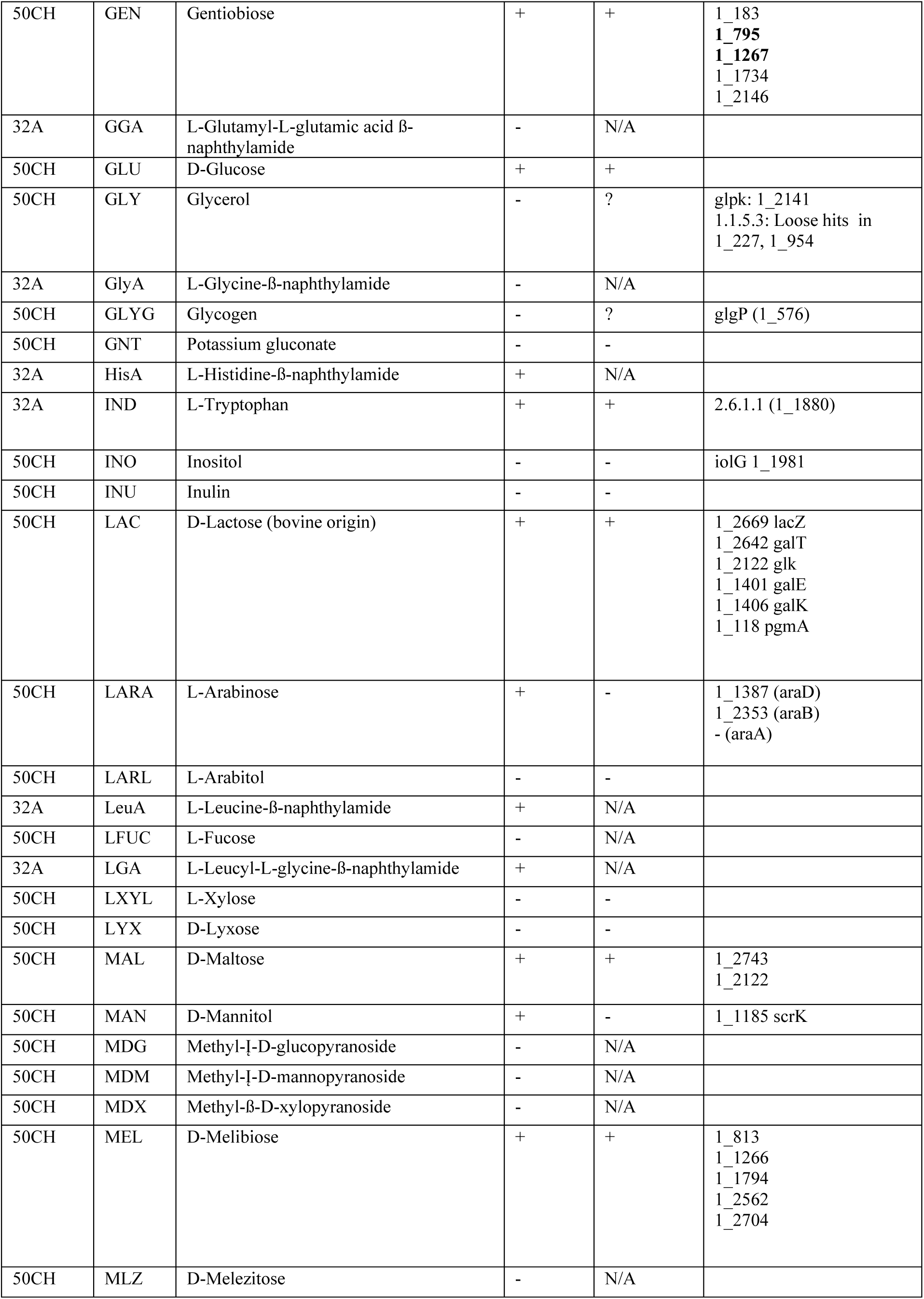
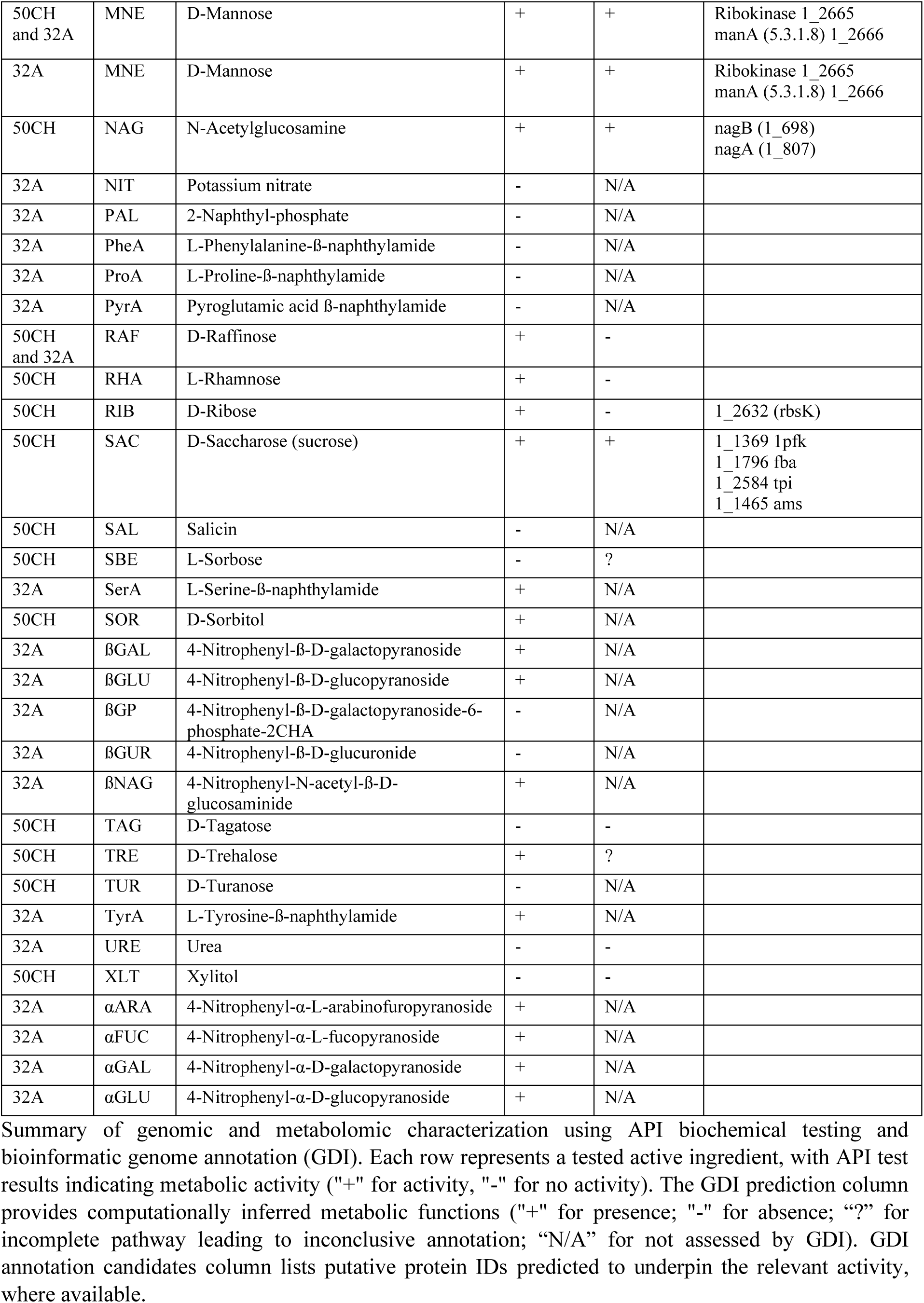
Genomic and metabolomic characterization.

**Supplementary Table 3:** MH27-2 production of aromatic lactic acids.

| Medium | ILA |  | PLA |  | HPLA |  |
| --- | --- | --- | --- | --- | --- | --- |
|  | YG/V | YG/V + IPyA | YG/V | YG/V + PPyA | YG/V | YG/V + HPPyA |
| Concentration of respective ALA in base medium (mM) | <LLOQ | <LLOQ | 19.79<br>±2.01 | 23.04<br>±1.97 | <LLOQ | <LLOQ |
| Concentration of respective ALA in medium following growth of MH27-2 (mM) | 2.01<br>±0.06 | 456.72<br>±38.69 | 42.09<br>±0.06 | 508.12<br>±38.72 | 41.56<br>±3.82 | 268.1<br>±21.59 |

MH27-2 was found to produce high quantities of ILA, PLA and HPLA when supplemented with 1 mM of the respective aromatic pyruvic acids. ALA = Aromatic lactic acid; IPyA = Indolepyruvic acid; ILA = Indolelactic acid; PPyA = Phenylpyruvic acid; PLA = Phenyllactic acid; HPPyA = 4-Hydroxyphenylpyruvic acid; HPLA = 4-Hydroxyphenyllactic acid; LLOQ = The lowest level of calibration (< 1mM for ILA, PLA and HPLA).

**Supplementary Table 4:**
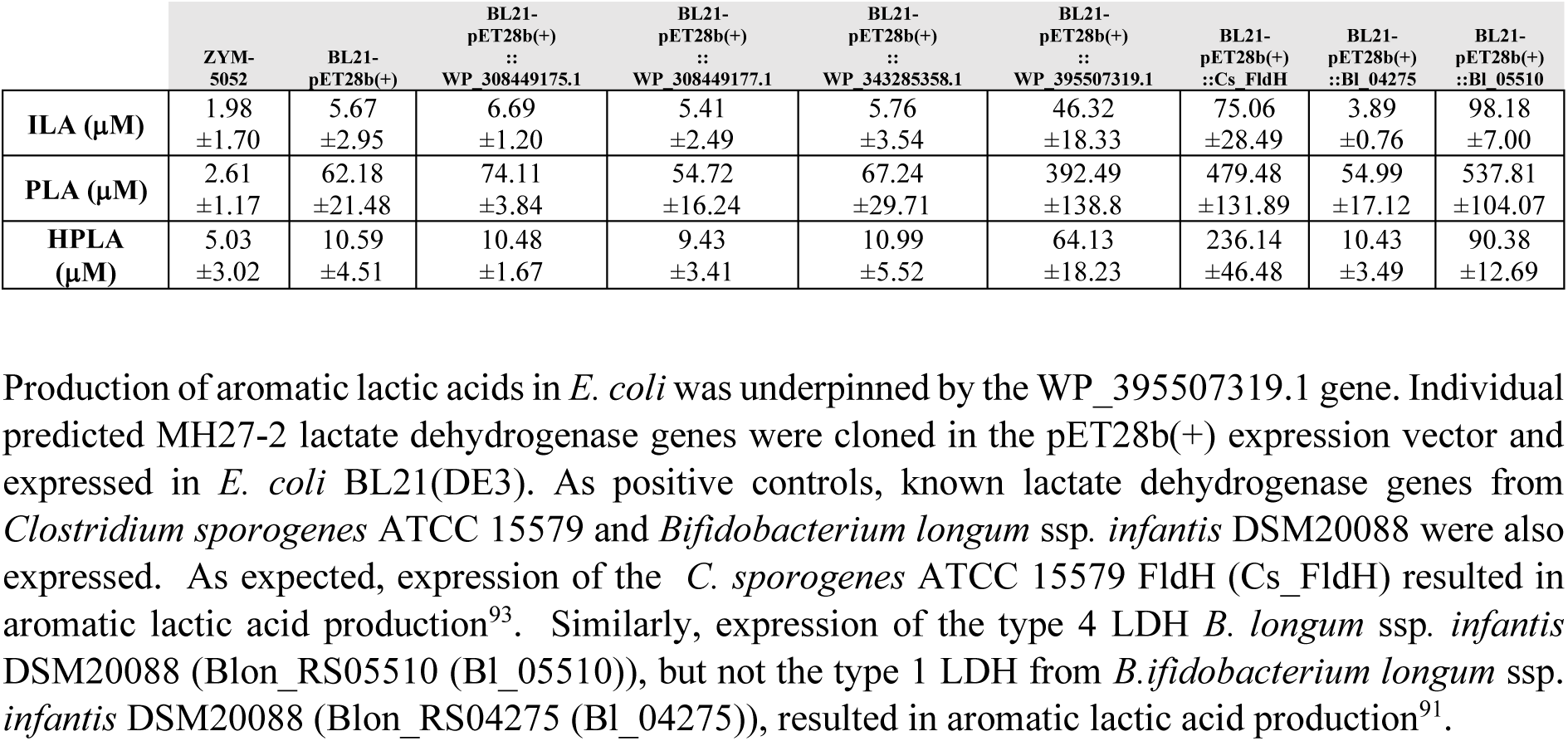
Production of aromatic lactic acids by heterologous expression of recombinant MH27-2 lactate dehydrogenases in *Escherichia coli* BL21(DE3).

**Supplementary Table 5:** *In silico* and *in vitro* safety.

| Category | Protein ID | Annotation | Function |
| --- | --- | --- | --- |
| AMR | WP_002586627.1 | tet(W) | Tetracycline resistance |
|  | WP_308448114.1 | ABC-F type ribosomal protection protein | Macrolide resistance |
|  | WP_308449231.1 | ant(6) | Streptomycin resistance |
| VFDB | WP_176820117.1 | dnaK chaperone protein DnaK | Adherence VFC0001 |
|  | WP_176820779.1 | plr/gapA type I glyceraldehyde-3-phosphate dehydrogenase | Adherence VFC0001 |
|  | WP_176821231.1 | tuf elongation factor Tu | Adherence VFC0001 |
|  | WP_308448692.1 | groEL chaperonin GroEL | Adherence VFC0001 |
|  | WP_395506674.1 | eno phosphopyruvate hydratase | Adherence VFC0001 |
|  | WP_395506914.1 | CPE_RS09090 type IV pilus twitching motility protein PilT | Adherence VFC0001 |
|  | WP_178200657.1 | cylR2 cytolysin regulator R2 | Exotoxin VFC0235 |
|  | WP_176819707.1 | KPN2242_RS16175 glycosyltransferase family 2 protein | Immune modulation VFC0258 |
|  | WP_176821290.1 | galE UDP-glucose 4-epimerase GalE | Immune modulation VFC0258 |
|  | WP_308448224.1 | rfbA glucose-1-phosphate thymidyltransferase RfbA | Immune modulation VFC0258 |
|  | WP_308448225.1 | rfbB dTDP-glucose 4,6-dehydratase | Immune modulation VFC0258 |
|  | WP_308449692.1 | glf UDP-galactopyranose mutase | Immune modulation VFC0258 |
|  | WP_316687475.1 | galU UTP--glucose-1-phosphate uridylyltransferase GalU | Immune modulation VFC0258 |
|  | WP_395507109.1 | wbuZ glycosyl amidation-associated protein WbuZ | Immune modulation VFC0258 |
|  | WP_316519009.1 | pyrB aspartate carbamoyltransferase | Nutritional/Metabolic factor VFC0272 |
|  | WP_395506614.1 | sugC sn-glycerol-3-phosphate ABC transporter ATP-binding protein UgpC | Nutritional/Metabolic factor VFC0272 |
|  | WP_176821223.1 | sigA/rpoV RNA polymerase sigma factor | Regulation VFC0301 |
|  | WP_316519315.1 | clpP ATP-dependent Clp protease proteolytic subunit | Stress survival VFC0282 |
|  | WP_395506374.1 | clpC endopeptidase Clp ATP-binding chain C | Stress survival VFC0282 |
|  | WP_395507292.1 | clpE ATP-dependent protease | Stress survival VFC0282 |
|  | WP_395507364.1 | flmH short chain dehydrogenase/reductase family oxidoreductase | Motility VFC0204 |
| Biogenic amines | N/D | N/D | N/D |

Genomic annotations related to antimicrobial resistance (AMR) genes and virulence factors identified using the virulence facto database (VFDB) in the dataset. The "Protein ID" column lists unique identifiers for proteins within MAP315, "Annotation" lists the predicted function, and "Risk" column categorizes proteins into resistance mechanisms or virulence-associated pathways.

## FIGURE LEGENDS

**Figure S1.**
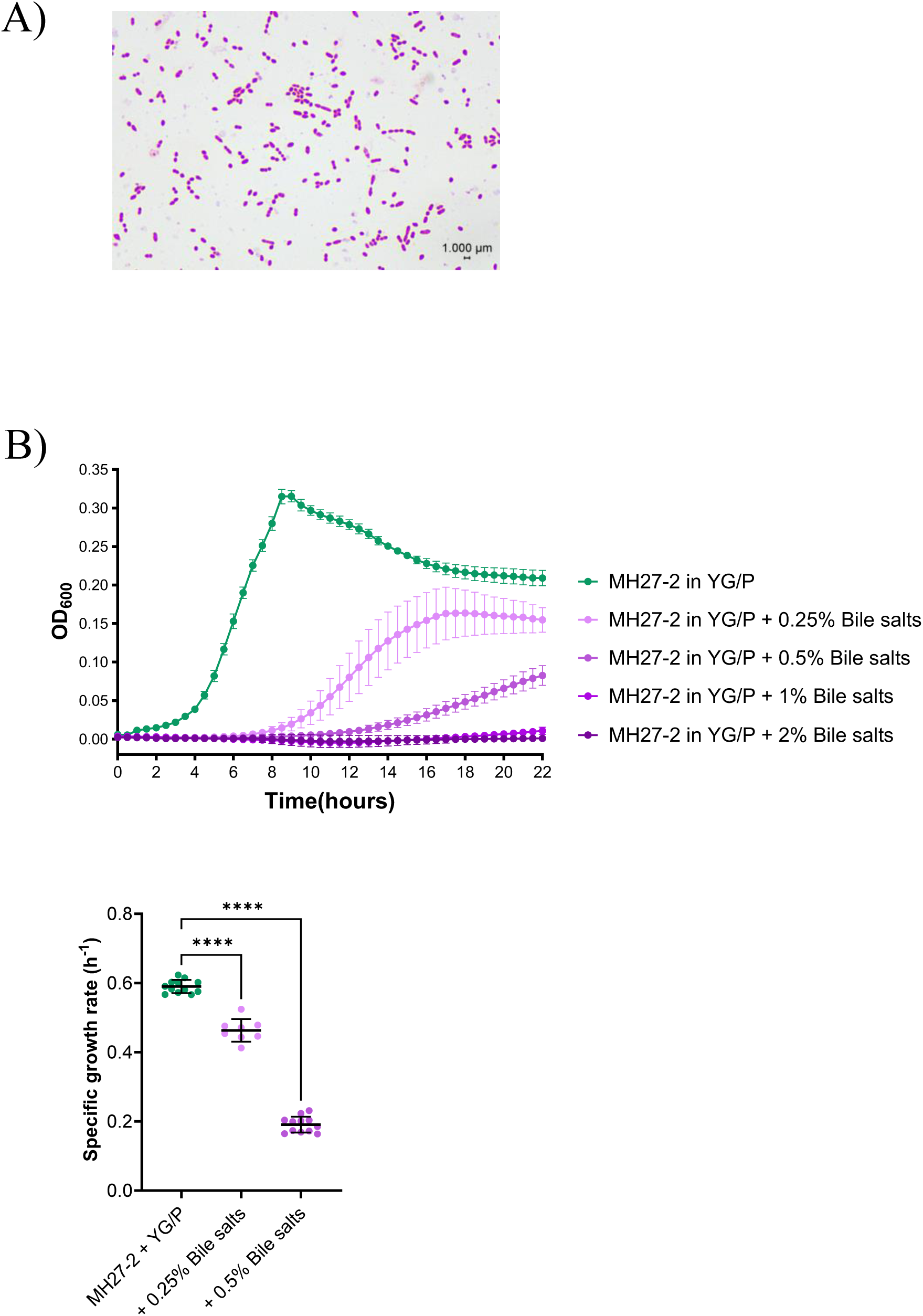
**A**. Microscopy image of Gram stained MH27-2 cells. **B.** MH27-2 growth in YG/P is affected by increasing concentrations of bile salts. The specific growth rate of MH27-2 is significantly higher in YG/P as compared to YG/P supplemented with 0.25 or 0.5% bile salts. All data are presented as average and standard deviation. Significance was determined using the ordinary one-way ANOVA. ****, *p* < 0.0001.

**Figure S2.**
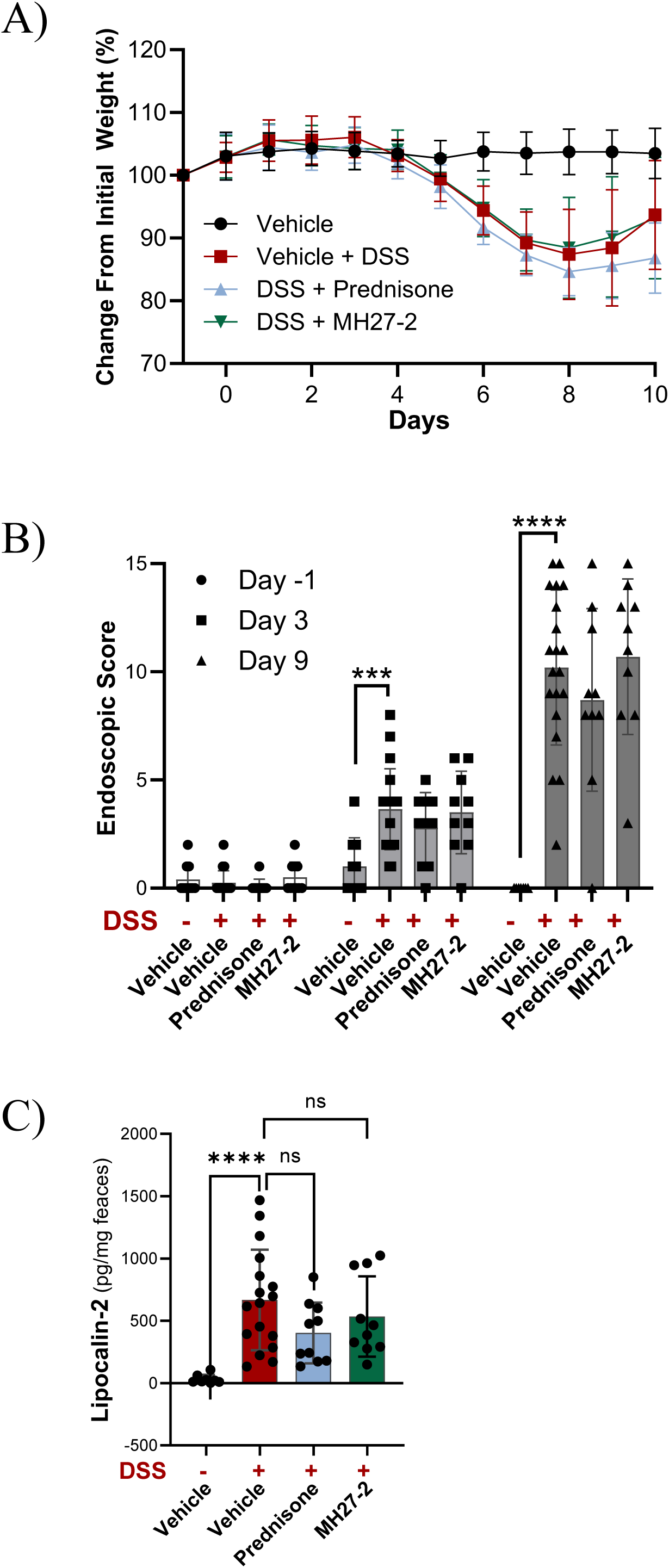
**A**. In a therapeutic model of DSS-induced acute murine colitis (Figure 2A), the body weight changes of mice were monitored over the course of the experiment. The body weight over time in mice treated with MH27-2 and DSS was comparable to that of mice receiving DSS alone. **B.** Endoscopy was performed on Day –1 for a baseline pre-DSS score, Day 3 one day prior to the commencement of LBP treatments, and Day 9 after six days of LBP treatment. For each time point, the different groups were compared to DSS + Vehicle group, using an uncorrected Brown-Forsythe and Welch ANOVA test with multiple comparisons. Data presented as mean with standard deviation. For all data, ns: not significant; *, p < 0.05; **, p < 0.01; ***, p < 0.001; ****, p < 0.0001. **C.** Lipocalin-2/NGAL ELISA was performed on fecal protein isolates collected at post-mortem and is presented as the concentration of Lipocalin-2/NGAL (pg) to weight of stool (mg). All groups were compared directly to the DSS + Vehicle group using the uncorrected Brown-Forsythe and Welch ANOVA test with multiple comparisons. Data presented as mean ± standard deviation. For all data, ns: not significant; *, p < 0.05; **, p < 0.01; ***, p < 0.001; ****, p < 0.0001.

**Figure S3.**
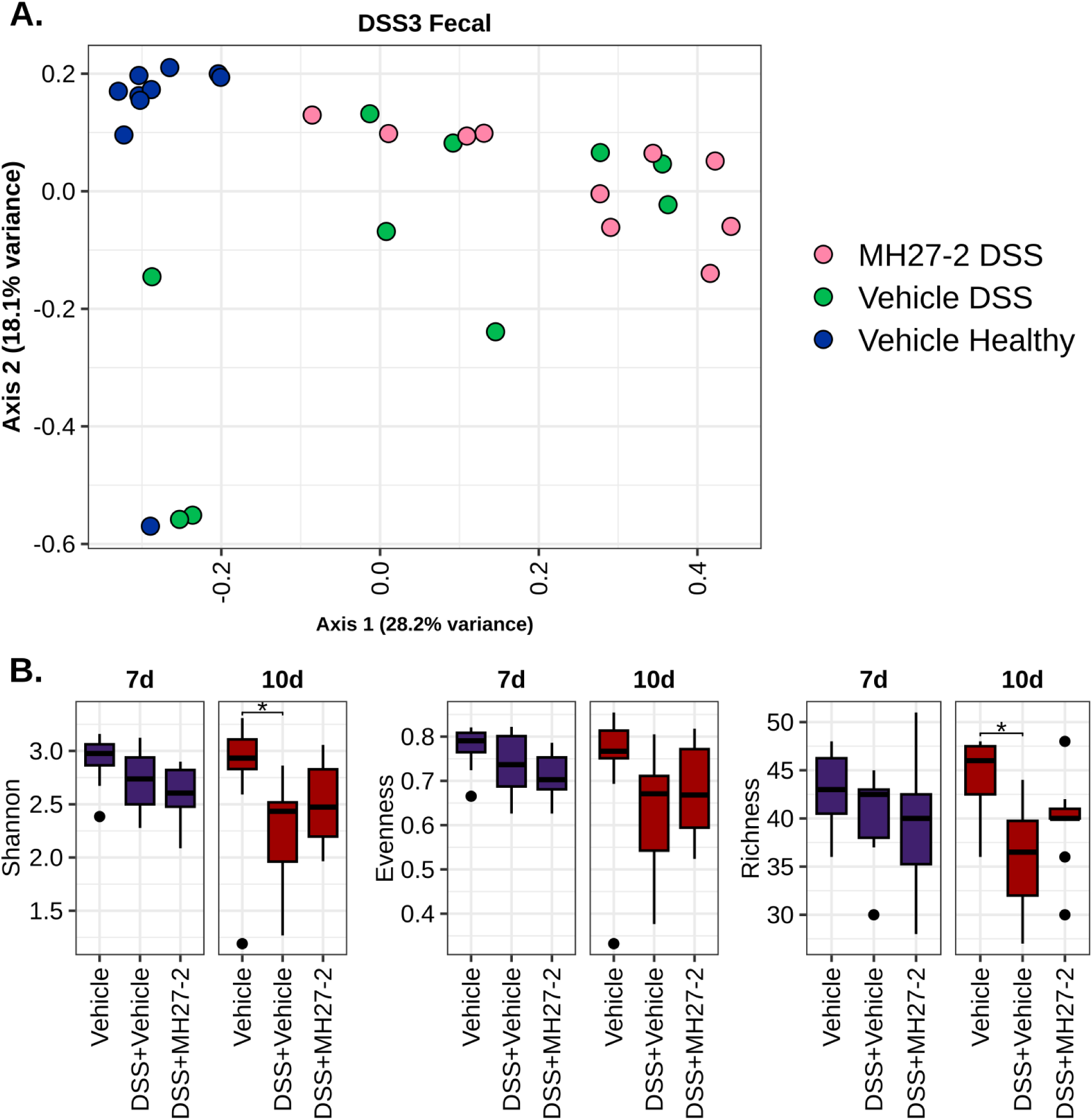
**A**. Effects on gut microbial community composition in a therapeutic model of DSS-induced acute murine colitis (Figure 2A). Principal Coordinates Analysis (PCoA) of fecal microbial profiles based on Bray-Curtis distances. Fecal pellet samples were collected from mice in three experimental groups: Vehicle (n=10, dark blue), DSS + Vehicle (n=10, green), and DSS + MH27-2 (n=10, pink). **B.** Microbial Shannon diversity (left), evenness (middle), and richness (right) at days 7 and 10 post-DSS treatment initiation. Statistical differences were assessed using t-tests with multiple test correction, where * indicates p < 0.05.

**Figure S4.**
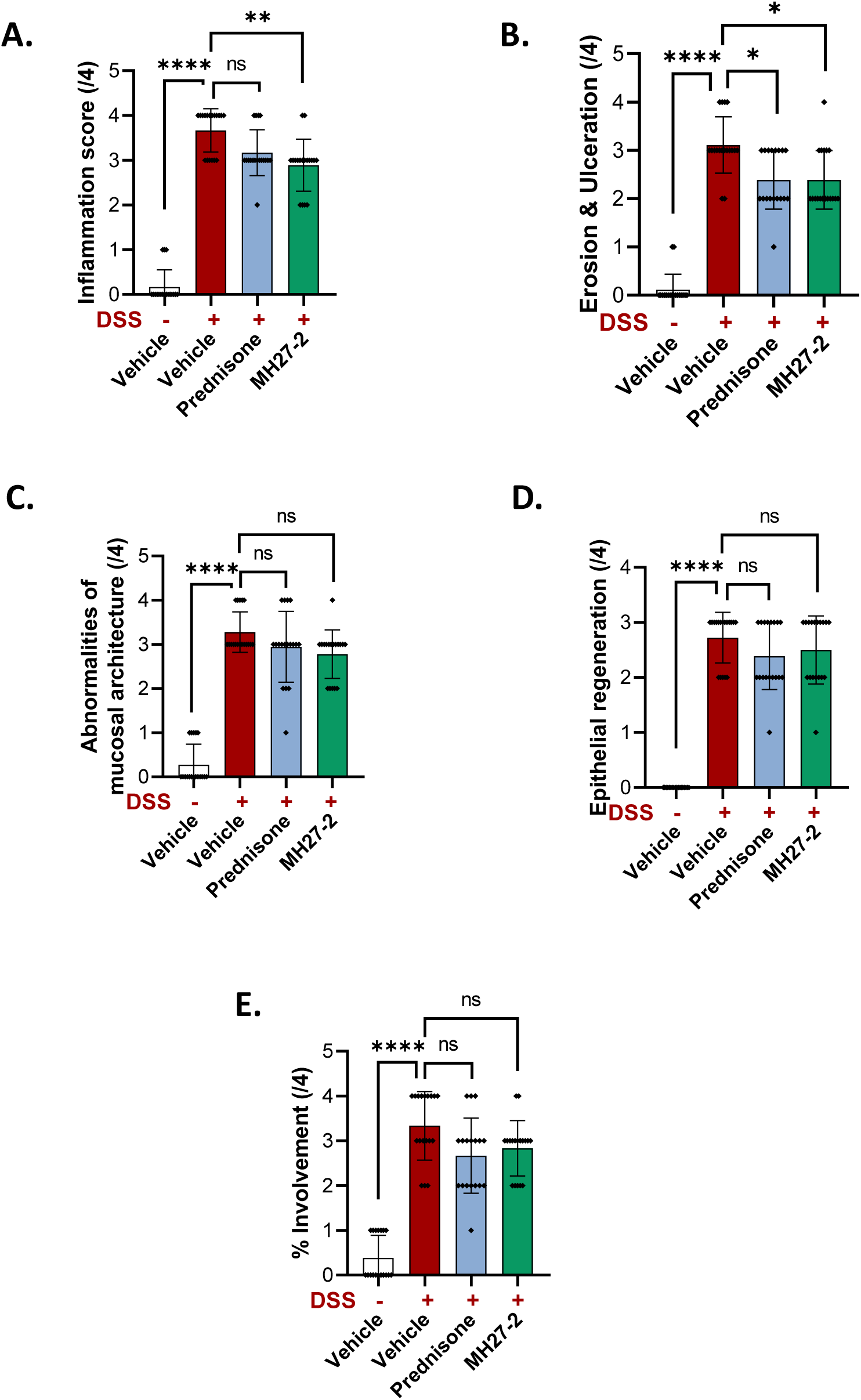
The prophylactic treatment of MH27-2 was evaluated in the DSS mouse model as presented in Figure 2F. In mice challenged with DSS, the co-treatment with live MH-27-2 significantly ameliorated inflammation (**A.**), as well as erosion and ulceration (**B.**), compared to mice exposed to DSS only. Although improvements in abnormalities of mucosal architecture (**C.**), epithelial regeneration (**D.**) and the percentage of tissue involved (**E.**) were also observed in mice treated with both DSS and MH27-2, these changes were not statistically significant. For all data, ns: not significant; *, p < 0.05; **, p < 0.01; ***, p < 0.001; ****, p < 0.0001; presented is the mean and standard deviation. The Kruskal-Wallis test with uncorrected Dunn’s multiple comparisons was used for statistical testing.

**Figure S5.**
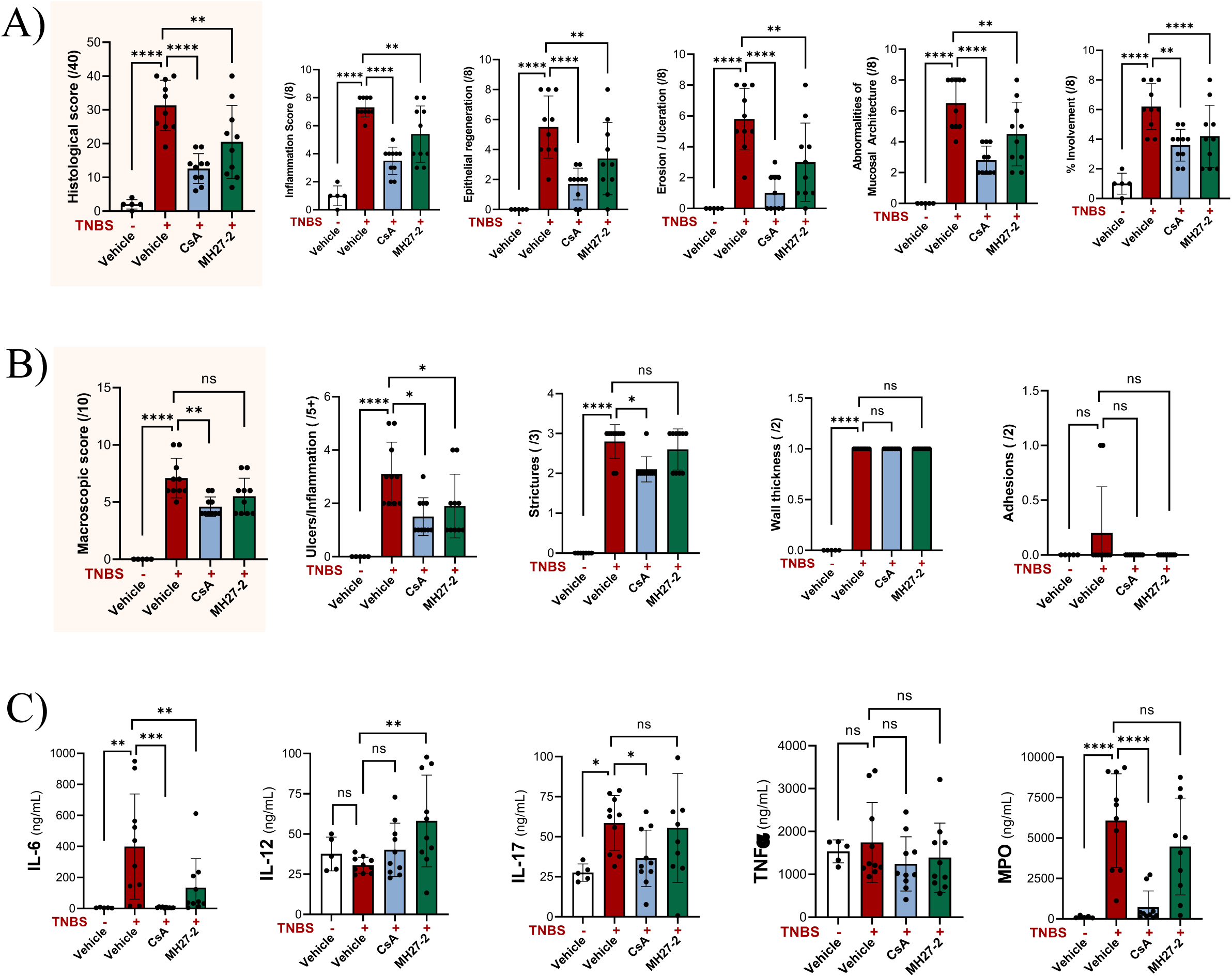
Prophylactic effects of MH27-2 were assessed in a mouse model of acute murine colitis that uses the haptenizing agent 2,4,6-Trinitrobenzene sulfonic acid (TNBS) to cause predominately Th1-driven pro-inflammatory responses in the murine colon characteristic of CD in humans (experimental setup presented in Figure 2K). **A.** MH27-2 ameliorated TNBS induced colitis as evidenced by the total histological score (total colitis index), that is the sum of five assessed sub-scores including inflammation, epithelial regeneration, erosion/ulceration, abnormalities of mucosal architecture, and lastly, the percentage involvement. One-way ANOVA with uncorrected Fisher’s LSD test for multiple comparison. **B.** The macroscopic score is composed of the four sub-scores ulcers/inflammation, strictures, wall thickness, and adhesions. Treatment with MH27-2 significantly improved the occurrence of ulcers and inflammation, but had no effect on the other three macroscopic readouts. Kruskal-Wallis test with uncorrected Dunn’s multiple comparisons. **C.** Compared to the DSS vehicle control, MH27-2 reduced levels of pro-inflammatory cytokine interleukin-6 (IL-6) in colonic tissue, had no effect on IL-17, TNF, MPO, and increased levels of IL-12. Kruskal-Wallis test with uncorrected Dunn’s multiple comparisons was used for IL-6, and for the remaining cytokines a one-way ANOVA with uncorrected Fisher’s LSD test for multiple comparison. For all data in Figure S5, ns: not significant; *, p < 0.05; **, p < 0.01; ***, p < 0.001; ****, p < 0.0001; presented is the mean and standard deviation.

**Figure S6.**
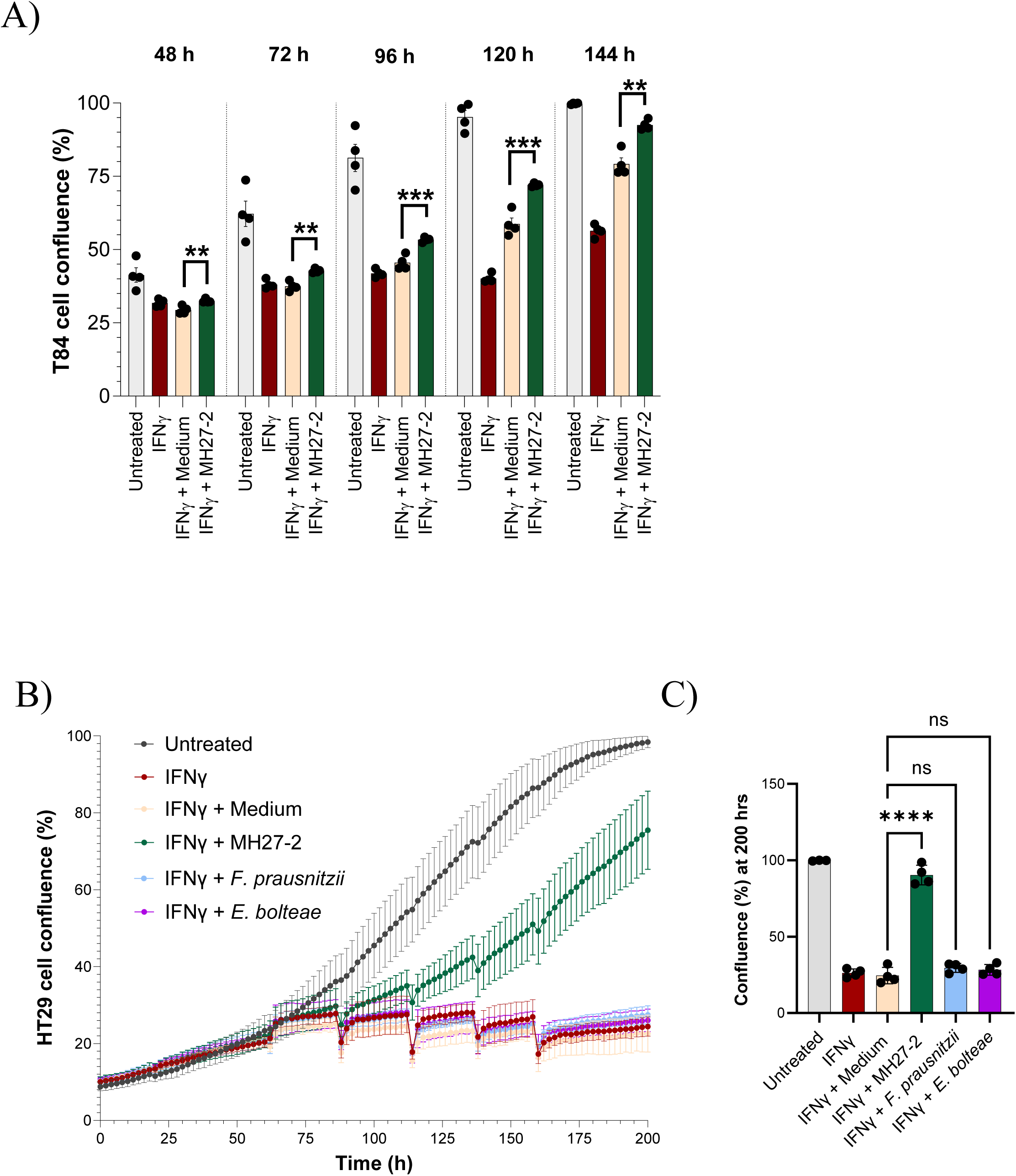
**A**. Proliferation of T84 cells is inhibited by the pro-inflammatory stimulus IFNγ and ameliorated by treatment with MH27-2 extract relative to the medium control extract. **B.** Proliferation of HT29 cells is inhibited by IFNγ and this effect is ameliorated by treatment MH27-2 extract but not medium control. The bacterial controls, *Enterocloster* (previously *Clostridium*) *bolteae* (strain ATCC BAA-613), a bacterium positively associated with IBD^38^, and *Faecalibacterium prausnitzii* (strain A2-165), a well-known anti-inflammatory gut bacterium depleted in IBD and currently evaluated as LBP candidate, did not promote proliferation. **C.** Endpoint analysis (200 h) for the proliferation data presented in **B.** with statistical analysis; Dunnett’s multiple comparisons test medium control versus bacterial extracts, ****, p < 0.0001.

**Figure S7.**
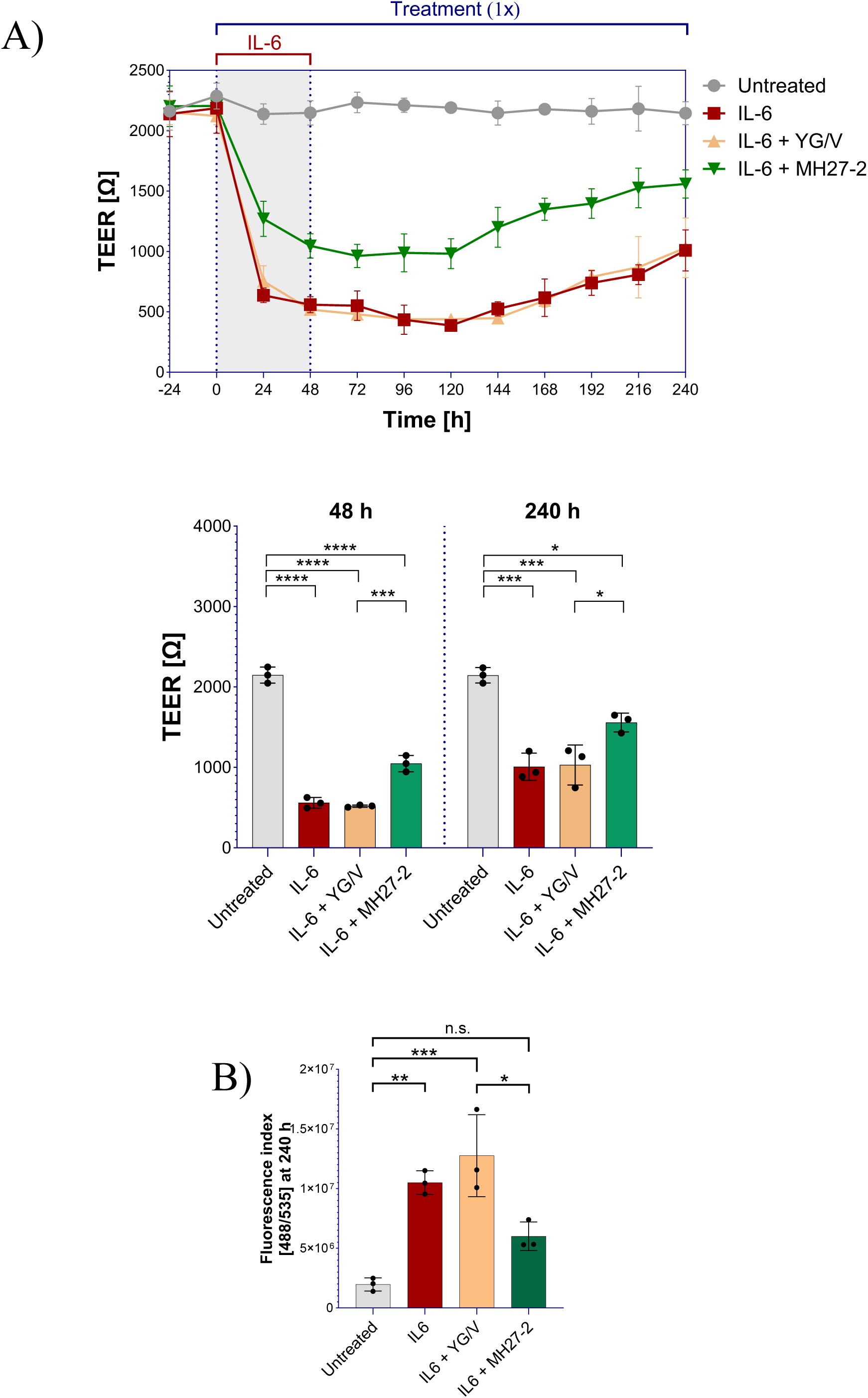
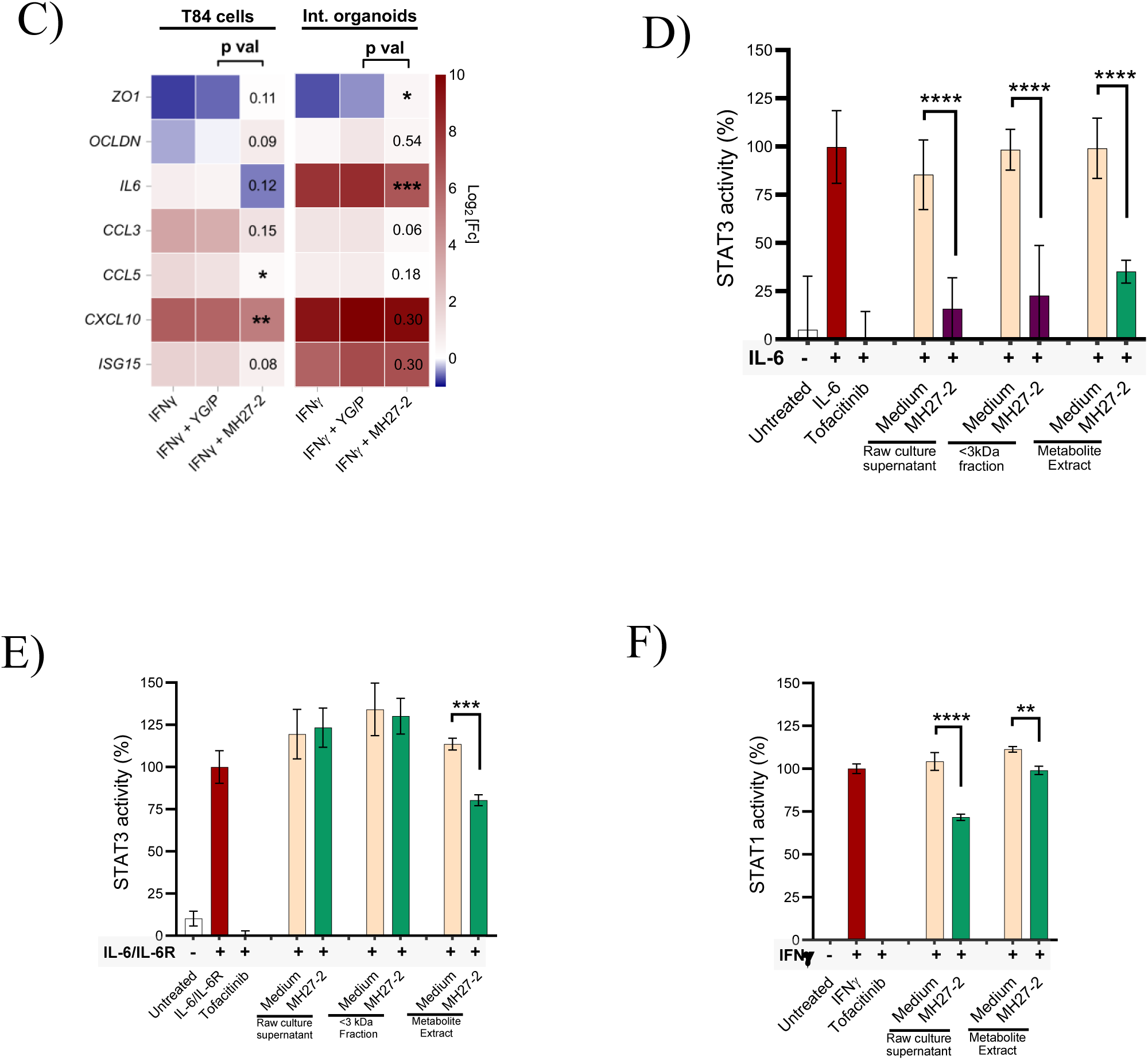
**A**. The effect of MH27-2 on barrier function of T84 gut epithelial cells was assessed in a trans-epithelial electrical resistance (TEER) model. For this, IL-6 was used as a barrier disruptor which, after 48 hours, led to an approximately 75% reduction in TEER, indicative of an increased barrier permeability, which naturally and gradually increased upon removal of the stimulus. T84 cells were treated with MH27-2 extract (1X) for one hour prior to challenge with IL-6 (100 ng/mL), and MH27-2 was replenished every 24 hours throughout the duration of the experiment. MH27-2 mitigated IL-6 mediated reduction in battier integrity, and promoted recovery compared to the medium control extract. One-way ANOVA at 48 and 240 hours. **B.** At experimental endpoint of the TEER assays presented in Figure S7A, cell permeability was assessed by paracellular translocation of FITC-labelled dextran. Compared to the medium control, MH27-2 extract significantly reduced the flux of FITC-dextran across the T84 cell monolayer as assessed through One-way ANOVA. **C.** In T84 cells and mouse intestinal organoids, MH27-2 metabolite extract (1X) mitigates IFNγ (100 ng/mL) mediated transcriptional changes of ZO1, OCLDN, and select pro-inflammatory drivers after 24 hours. Experiments performed in biological triplicates, two technical replicates each; paired t-test comparing IFNγ + YG/P medium control to IFNγ + MH27-2. **D.** MH27-2 culture supernatant (raw and <3kDa filtered), and MH27-2 extract (1X) significantly mitigated IL-6 mediated STAT3 activity in a HEK-Blue IL-6 reporter cell line. **E.** MH27-2 extract (1X) significantly mitigated IL-6/IL6R mediated STAT3 activity in a HEK-Blue IL-6 reporter cell line. **F.** Raw MH27-2 culture supernatant and MH27-2 extract (1X) significantly mitigated IFNγ-mediated STAT1 activity in a HEK-Blue IFNγ reporter cell line.

**Figure S8.**
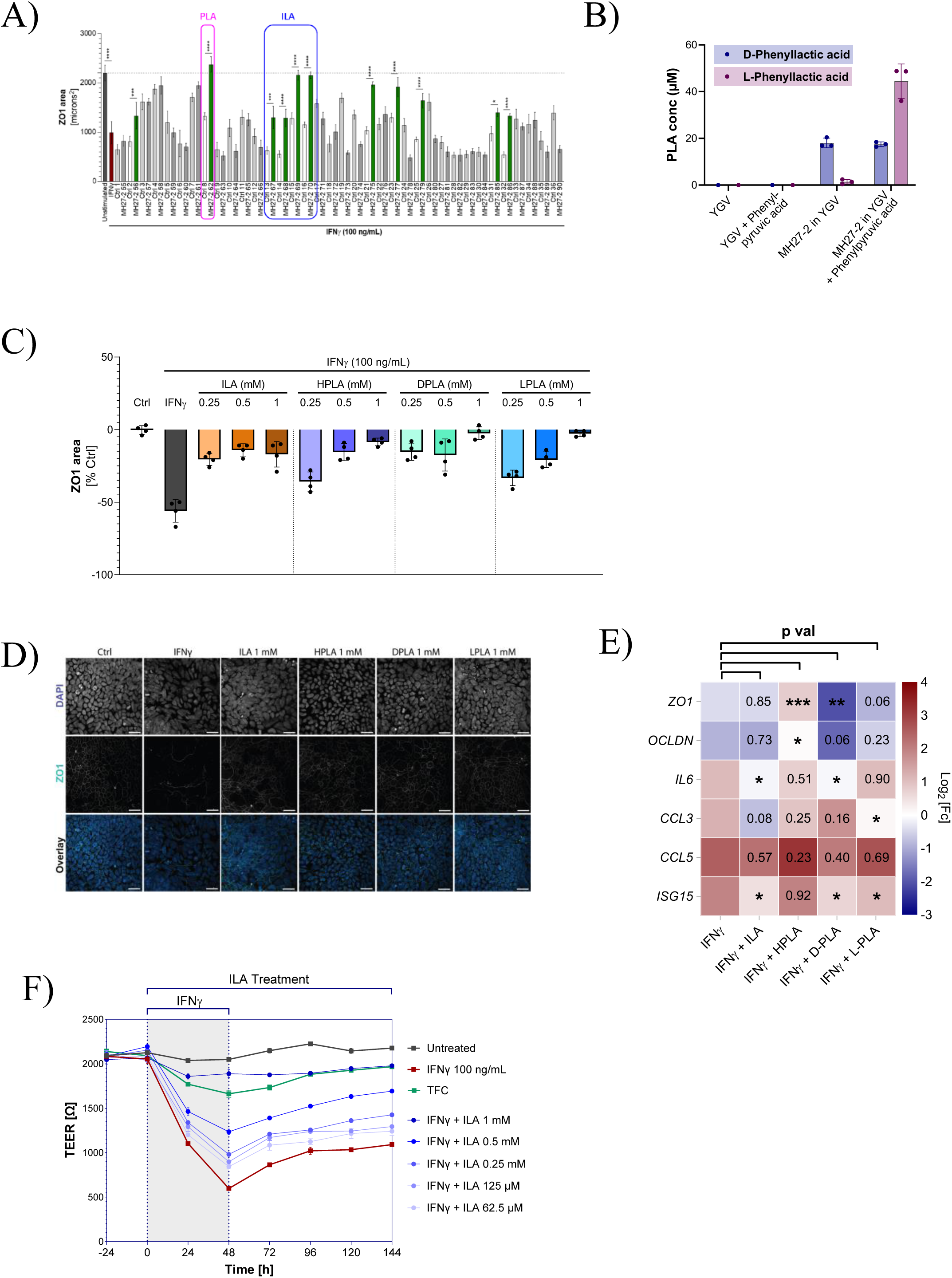
**A**. As assessed through confocal microscopy, one hour pre-treatment with select fractions of MH27-2 extract, known to contain PLA and ILA respectively, significantly mitigated IFNγ-mediated reduction in ZO-1 protein expression following 24 hours stimulation. **B.** As assessed by HPLC, MH27-2 principally produces D-PLA when grown in YG/V. Supplementation with phenylpyruvic acid results in increased L-PLA, but not D-PLA, production. **C.** As assessed through confocal microscopy, one hour pre-treatment with the aromatic lactic acids ILA, HPLA, D-PLA, and L-PLA followed by 24 hours stimulation with IFNγ resulted in dose-dependent protective effects on ZO-1 protein expression. **D.** Representative images of effect of aromatic lactic acids on ZO-1 protein expression. **E.** As assessed through RT-qPCR, one hour pre-treatment with ILA, HPLA, D-PLA, or L-PLA, followed by 24-hour stimulation with IFNγ significantly mitigated transcriptional changes of *ZO1* (HPLA and D-PLA) and *OCLDN* (HPLA), *IL6* (ILA and D-PLA), *CCL3* (L-PLA), and *ISG15* (ILA, D-PLA, and L-PLA). **F.** The effect of various concentrations of ILA on barrier function of T84 gut epithelial cells was assessed in a trans-epithelial electrical resistance (TEER) model. T84 cells were pre-treated with ILA of Tofacitinib (TFC; 80µM) for one hour prior to challenge with IFNγ (100 ng/mL), and treatments were replenished every 24 hours throughout the duration of the experiment. ILA mitigated IFNγ-mediated loss of barrier integrity in a dose dependent manner.

**Figure S9.**
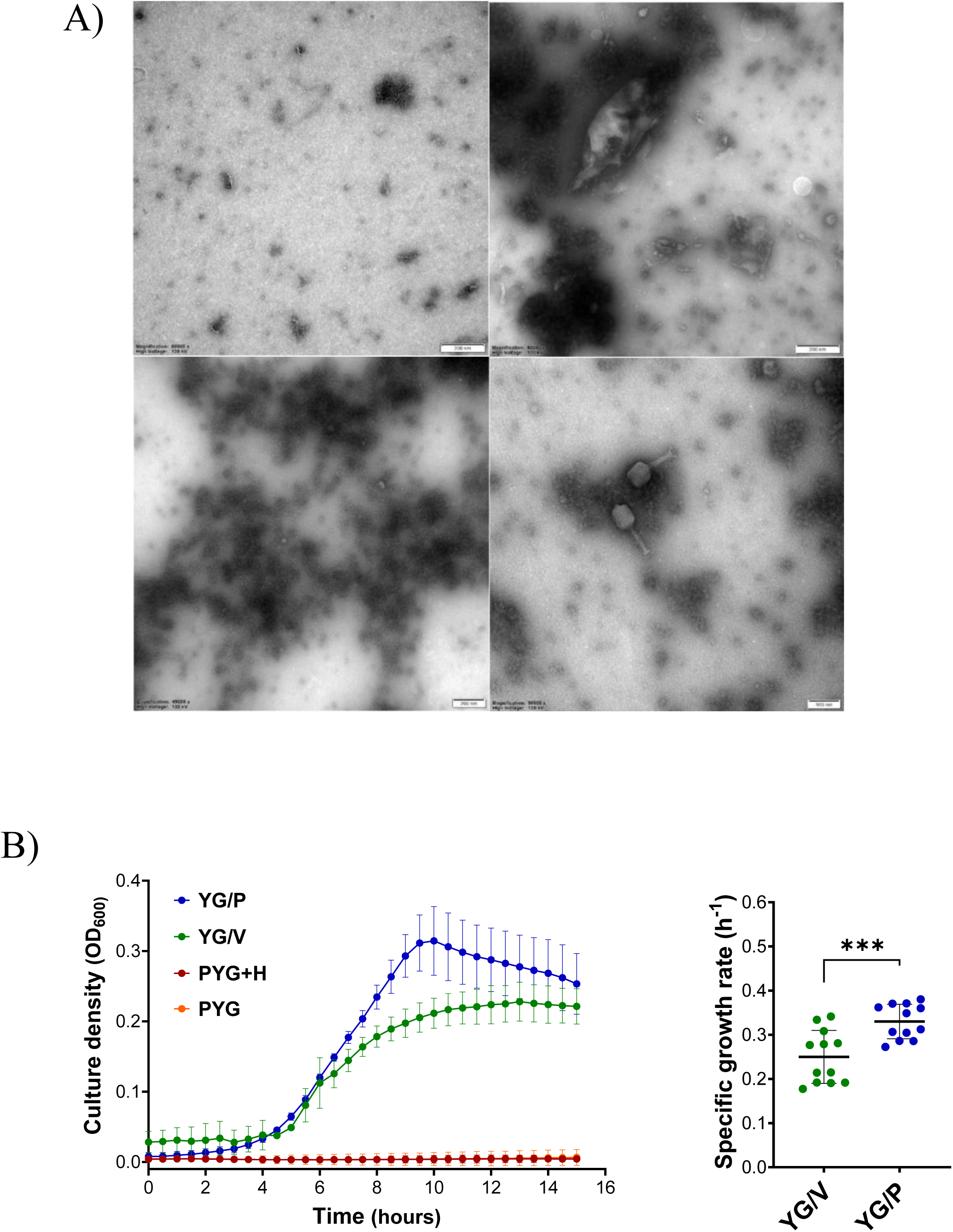
**A**. Testing for virulent phage and for inducible prophage presence. First round of enrichment of virulent phage test (left upper panel), second round of enrichment of virulent phage test (right upper panel), and test for inducible prophage (left lower panel) are represented by typical field of view. Positive control is right lower panel. **B.** A specific growth rate of 0.33 h^-1^ was measured and maximum culture density was achieved after ten hours when MH27-2 was cultured in YG/P animal component-free medium. In comparison, a growth rate of 0.25 h^-1^ and maximum culture density after 13 hours was reached in YG/V animal component-free medium. No growth was observed in PYG medium containing casein-derived tryptone (**Figure S10B**).

**Figure S10.**
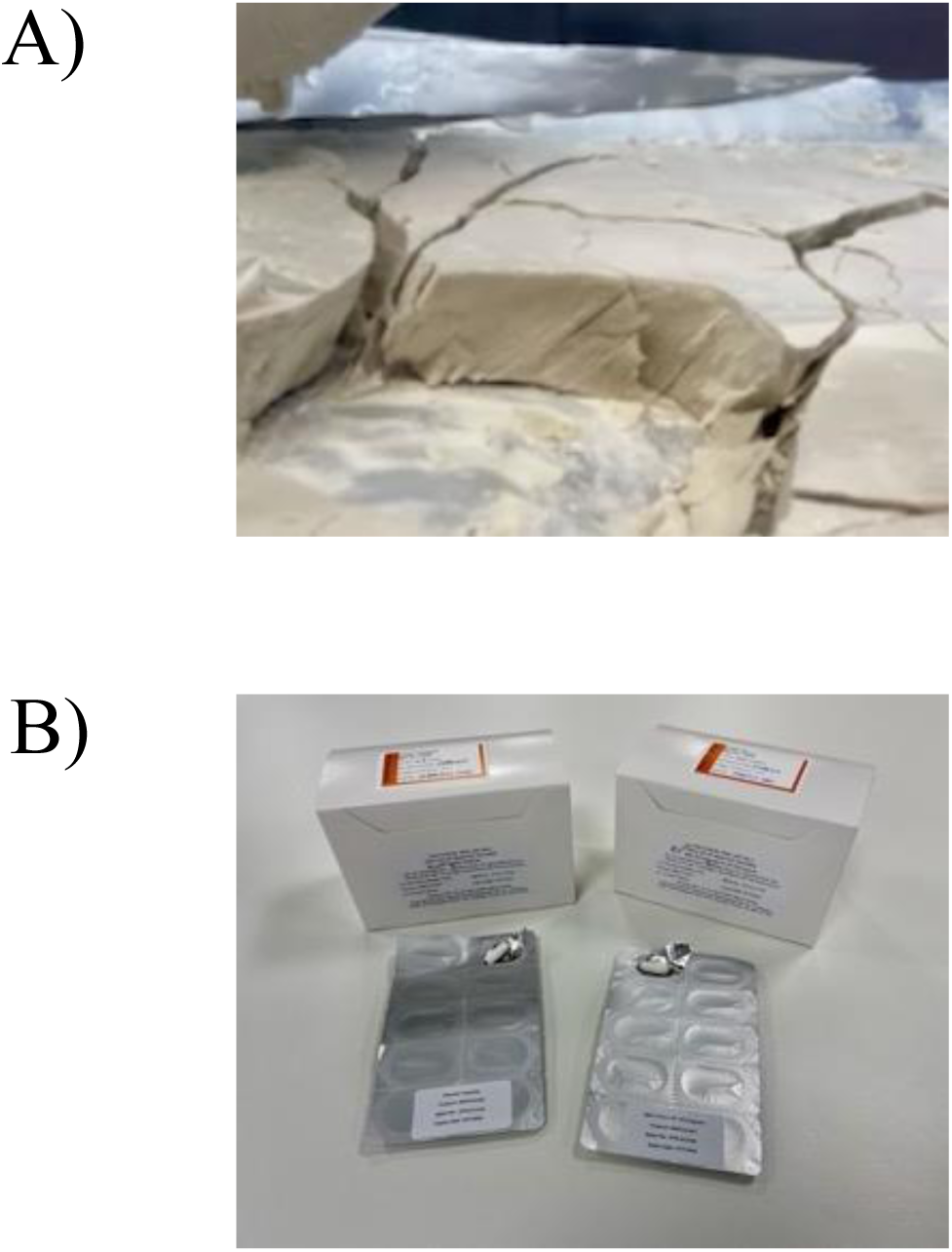
Images of MAP 315 drug substance (**A.**) and drug product (**B.**)

**Figure S11.**
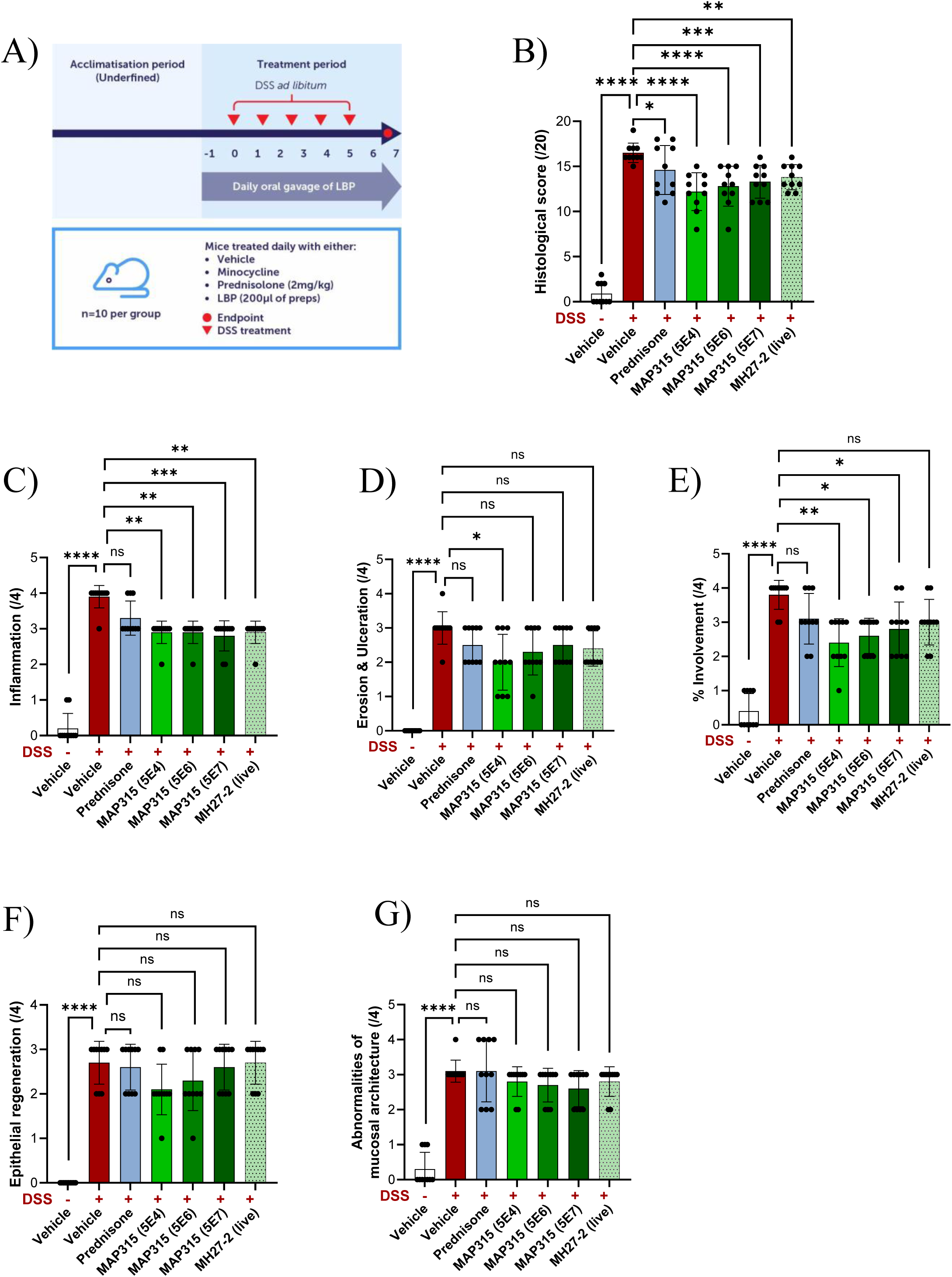
**A**. The therapeutic efficacy of MAP 315 drug substance in three different doses was assessed in a prophylactic model of DSS-induced murine colitis, and compared to freshly grown live MH27-2. Both the drug substance and freshly grown MH27-2 significantly improved the total histopathological score (**B.**), which includes the five sub-scores inflammation (**C.**), erosion and ulceration (**D.**), percentage of the colonic epithelium affected (**E.**), epithelial regeneration (**F.**) and abnormalities of mucosal architecture (**G.**). There was no significant difference in the efficacy of the MAP 315 drug substance and freshly grown MH27-2 cells, indicating that the drug substance manufacturing process did not negatively impact therapeutic efficacy (Total histology score MH27 live vs. MAP315 (5E7) p=0.56; MAP315 (5E6) p=0.25; MAP315 (5E4) p = 0.07; For the subscores, all comparisons p>0.05 except for epithelial regeneration MH27-2 live vs MAP 315 5e4 p=0.02). The statistical test for the total histology score was a one-way ANOVA with uncorrected Fisher’s LSD test for multiple comparisons. For the five subscores, a Kruskal-Wallis test with uncorrected Dunn’s multiple comparisons was used. Data is presented as the mean and SD, and ns: not significant; *, p < 0.05; **, p < 0.01; ***, p < 0.001; ****, p < 0.0001.

**Figure S12.**
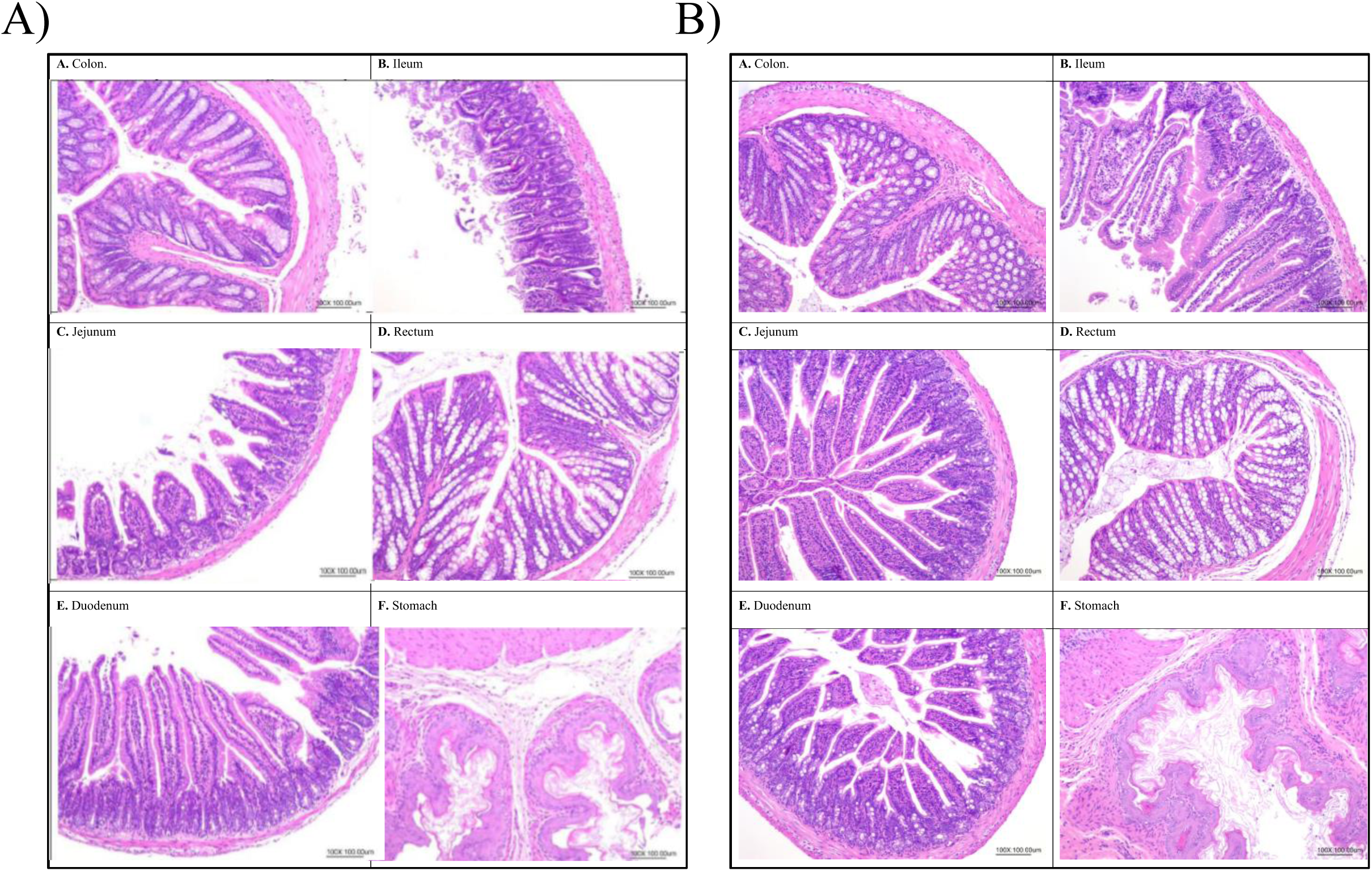
**A**. Representative images of histopathologic findings from the murine tolerability study. Healthy C57BL/6 mice were treated with 5 x 10^6^, 5 x 10^7^ or 1 x 10^8^ CFU/mouse/day of MAP 315 for 14 days by oral administration to evaluate safety and tolerability. The gastrointestinal tracts, spleens and mesenteric lymph nodes of the treated mice were evaluated by histopathology and there were no atypical findings. Representative pathology images from the colon, ileum, jejunum, rectum, duodenum, and stomach of the (**A.**) vehicle group and (**B.**) high dose groups (1 x 10^8^ CFU/mouse/day of MAP 315) are shown.

**Figure S13.**
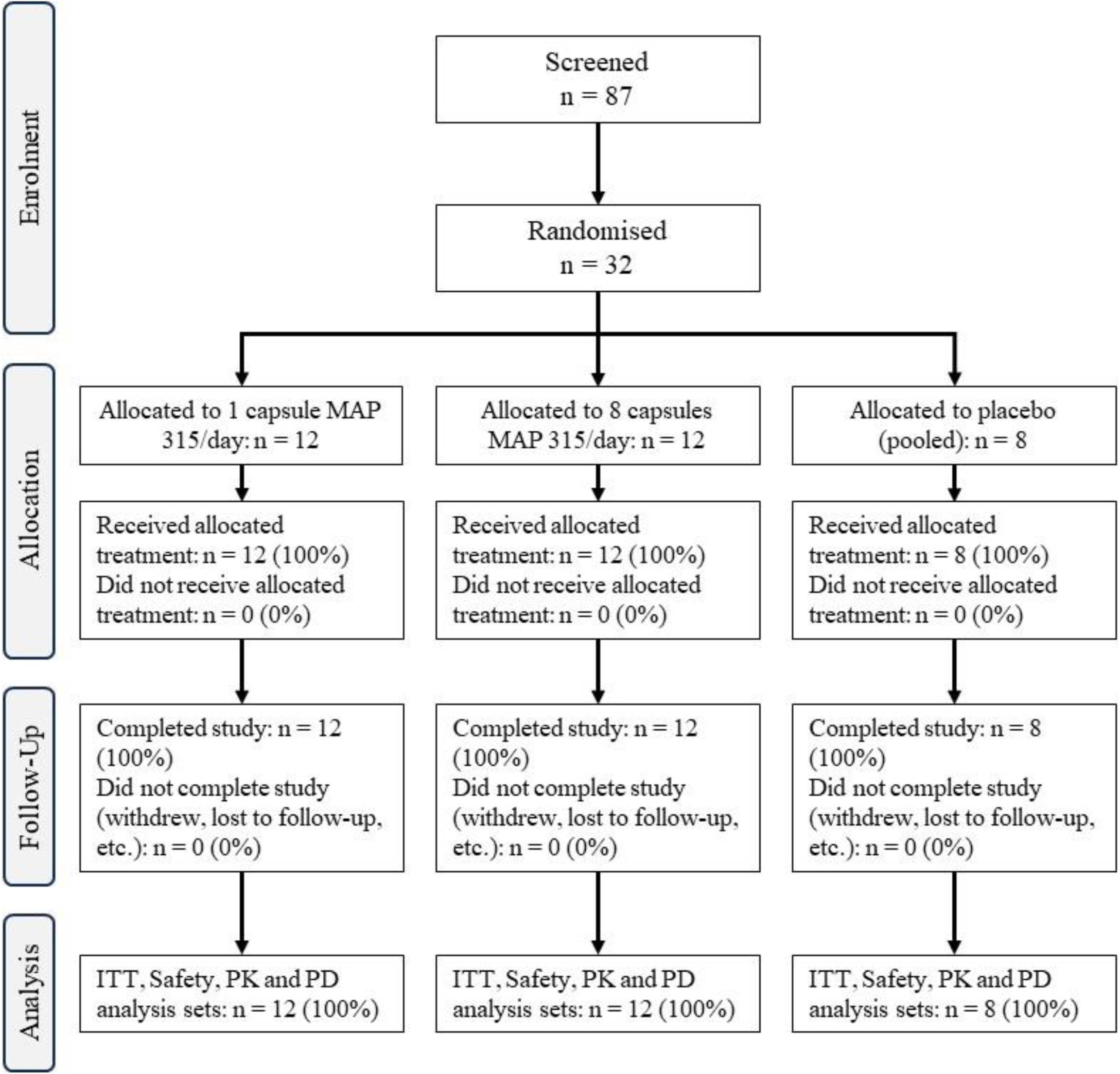
A Phase 1 clinical study was conducted to assess the safety and tolerability of the MAP 315 drug product in humans. This single-center, randomized, double-blind, placebo-controlled study involved multiple doses administered to 32 healthy adults. Participants were randomized in a 3:1 ratio to receive either MAP 315 or a matching placebo for 14 consecutive days and were divided into two cohorts based on dosage: a low-dose group (one MAP 315 or placebo capsule daily) and a high-dose group (eight MAP 315 or placebo capsules daily). The study was completed as planned, with all participants finishing the trial.

## References

1. Saez A, Herrero-Fernandez B, Gomez-Bris R, Sánchez-Martinez H, Gonzalez-Granado JM. Pathophysiology of inflammatory bowel disease: innate immune system. International Journal of Molecular Sciences. 2023;24(2):1526.

2. Xavier RJ, Podolsky DK. Unravelling the pathogenesis of inflammatory bowel disease. Nature. 2007;448(7152):427–434. doi:10.1038/nature06005

3. Wang R, Li Z, Liu S, Zhang D. Global, regional and national burden of inflammatory bowel disease in 204 countries and territories from 1990 to 2019: a systematic analysis based on the Global Burden of Disease Study 2019. BMJ open. 2023;13(3):e065186.

4. Singh S, Blanchard A, Walker JR, Graff LA, Miller N, Bernstein CN. Common symptoms and stressors among individuals with inflammatory bowel diseases. Clinical Gastroenterology and Hepatology. 2011;9(9):769–775.

5. Imhann F, Vich Vila A, Bonder MJ, et al. Interplay of host genetics and gut microbiota underlying the onset and clinical presentation of inflammatory bowel disease. Gut. 2018;67(1):108–119. doi:10.1136/gutjnl-2016-312135

6. Higashiyama M, Hokari R. New and emerging treatments for inflammatory bowel disease. Digestion. 2023;104(1):74–81.

7. Cai Z, Wang S, Li J. Treatment of Inflammatory Bowel Disease: A Comprehensive Review. Front Med (Lausanne). 2021;8:765474. doi:10.3389/fmed.2021.765474

8. Peyrin-Biroulet L, Sandborn W, Sands BE, et al. Selecting Therapeutic Targets in Inflammatory Bowel Disease (STRIDE): Determining Therapeutic Goals for Treat-to-Target. Am J Gastroenterol. 2015;110(9):1324–1338. doi:10.1038/ajg.2015.233

9. Le Berre C, Peyrin-Biroulet L, Sandborn WJ, et al. Selecting End Points for Disease-Modification Trials in Inflammatory Bowel Disease: the SPIRIT Consensus From the IOIBD. Gastroenterology. 2021;160(5):1452–1460.e21. doi:10.1053/j.gastro.2020.10.065

10. Ungaro R, Colombel JF, Lissoos T, Peyrin-Biroulet L. A Treat-to-Target Update in Ulcerative Colitis: A Systematic Review. Am J Gastroenterol. 2019;114(6):874–883. doi:10.14309/ajg.0000000000000183

11. Neurath MF, Travis SPL. Mucosal healing in inflammatory bowel diseases: a systematic review. Gut. 2012;61(11):1619–1635. doi:10.1136/gutjnl-2012-302830

12. Colombel JF, Rutgeerts P, Reinisch W, et al. Early mucosal healing with infliximab is associated with improved long-term clinical outcomes in ulcerative colitis. Gastroenterology. 2011;141(4):1194–1201. doi:10.1053/j.gastro.2011.06.054

13. Rath T, Atreya R, Bodenschatz J, et al. Intestinal barrier healing is superior to endoscopic and histologic remission for predicting major adverse outcomes in inflammatory bowel disease: the Prospective ERIca trial. Gastroenterology. 2023;164(2):241–255.

14. Elhag DA, Kumar M, Saadaoui M, et al. Inflammatory bowel disease treatments and predictive biomarkers of therapeutic response. International journal of molecular sciences. 2022;23(13):6966.

15. Jacobse J, Li J, Rings EH, Samsom JN, Goettel JA. Intestinal regulatory T cells as specialized tissue-restricted immune cells in intestinal immune homeostasis and disease. Frontiers in Immunology. 2021;12:716499.

16. Zaiatz Bittencourt V, Jones F, Doherty G, Ryan EJ. Targeting immune cell metabolism in the treatment of inflammatory bowel disease. Inflammatory bowel diseases. 2021;27(10):1684–1693.

17. Hashash JG, Abou Fadel CG, Rimmani HH, Sharara AI. Biologic monotherapy versus combination therapy with immunomodulators in the induction and maintenance of remission of Crohn’s disease and ulcerative colitis. Annals of Gastroenterology. 2021;34(5):612.

18. Atreya R, Neurath MF. Current and future targets for mucosal healing in inflammatory bowel disease. Visceral medicine. 2017;33(1):82–88.

19. Ghosh S, Whitley CS, Haribabu B, Jala VR. Regulation of intestinal barrier function by microbial metabolites. Cellular and molecular gastroenterology and hepatology. 2021;11(5):1463–1482.

20. Zheng D, Liwinski T, Elinav E. Interaction between microbiota and immunity in health and disease. Cell research. 2020;30(6):492–506.

21. Morgan XC, Tickle TL, Sokol H, et al. Dysfunction of the intestinal microbiome in inflammatory bowel disease and treatment. Genome Biology. 2012;13(9):R79. doi:10.1186/gb-2012-13-9-r79

22. Franzosa EA, Sirota-Madi A, Avila-Pacheco J, et al. Gut microbiome structure and metabolic activity in inflammatory bowel disease. Nat Microbiol. 2019;4(2):293–305. doi:10.1038/s41564-018-0306-4

23. Halfvarson J, Brislawn CJ, Lamendella R, et al. Dynamics of the human gut microbiome in inflammatory bowel disease. Nat Microbiol. 2017;2:17004. doi:10.1038/nmicrobiol.2017.4

24. Johansson MEV, Gustafsson JK, Holmén-Larsson J, et al. Bacteria penetrate the normally impenetrable inner colon mucus layer in both murine colitis models and patients with ulcerative colitis. Gut. 2014;63(2):281–291. doi:10.1136/gutjnl-2012-303207

25. Qiu P, Ishimoto T, Fu L, Zhang J, Zhang Z, Liu Y. The Gut Microbiota in Inflammatory Bowel Disease. Frontiers in Cellular and Infection Microbiology. 2022;12. Accessed September 22, 2022. https://www.frontiersin.org/articles/10.3389/fcimb.2022.733992

26. Zhang T, Li X, Li J, Sun F, Duan L. Gut microbiome-targeted therapies as adjuvant treatments in inflammatory bowel diseases: a systematic review and network meta-analysis. Journal of Gastroenterology and Hepatology. 2025;40(1):78–88.

27. Ghoshal UC, Gwee K, Holtmann G, et al. The role of the microbiome and the use of probiotics in gastrointestinal disorders in adults in the Asia-Pacific region-background and recommendations of a regional consensus meeting. Journal of gastroenterology and hepatology. 2018;33(1):57–69.

28. Pribyl AL, Hugenholtz P, Cooper MA. Advances in human gut microbiome-derived live biotherapeutics: Learnings from the last decade of research and development. Nature Microbiology (in press).

29. Naegeli AN, Hunter T, Dong Y, et al. Full, partial, and modified permutations of the Mayo score: characterizing clinical and patient-reported outcomes in ulcerative colitis patients. Crohn’s & Colitis 360. 2021;3(1):otab007.

30. Alavinejad P, Hashemi SJ, Behl N, et al. Inflammatory bowel disease evolution in the past two decades: a chronological multinational study. EClinicalMedicine. 2024;70.

31. Ko CW, Singh S, Feuerstein JD, et al. AGA Clinical Practice Guidelines on the Management of Mild-to-Moderate Ulcerative Colitis. Gastroenterology. 2019;156(3):748–764. doi:10.1053/j.gastro.2018.12.009

32. Ham M, Moss AC. Mesalamine in the treatment and maintenance of remission of ulcerative colitis. Expert review of clinical pharmacology. 2012;5(2):113–123.

33. Ford AC, Achkar JP, Khan KJ, et al. Efficacy of 5-aminosalicylates in ulcerative colitis: systematic review and meta-analysis. Official journal of the American College of Gastroenterology| ACG. 2011;106(4):601–616.

34. Sutherland LR, MacDonald JK. Oral 5-aminosalicylic acid for maintenance of remission in ulcerative colitis. Cochrane Database of Systematic Reviews. 2006;(2).

35. Mikami Y, Tsunoda J, Suzuki S, Mizushima I, Kiyohara H, Kanai T. Significance of 5-aminosalicylic acid intolerance in the clinical management of ulcerative colitis. Digestion. 2023;104(1):58–65.

36. Zheng J, Fan Z, Li C, Wang D, Zhang S, Chen R. Predictors for colectomy in patients with acute severe ulcerative colitis: a systematic review and meta-analysis. BMJ Open Gastroenterology. 2024;11(1):e001587.

37. Zhou J, Boyd JA, Nyeverecz B, et al. Draft genome sequence of two “Candidatus Intestinicoccus colisanans” strains isolated from faeces of healthy humans. BMC Research Notes. 2023;16(1):174.

38. Krause L, Boyd J, Vivian C, et al. A rational approach for the targeted discovery and characterisation of microbiome-derived therapeutics. bioRxiv. Published online 2024:2024-10.

39. Afrizal A, Hitch TC, Viehof A, et al. Anaerobic single-cell dispensing facilitates the cultivation of human gut bacteria. Environmental Microbiology. 2022;24(9):3861–3881.

40. Lagkouvardos I, Pukall R, Abt B, et al. The Mouse Intestinal Bacterial Collection (miBC) provides host-specific insight into cultured diversity and functional potential of the gut microbiota. Nature microbiology. 2016;1(10):1–15.

41. Zgheib R, Ibrahim A, Anani H, et al. N eglectibacter timonensis gen. nov., sp. nov. and Scatolibacter rhodanostii gen. nov., sp. nov., two anaerobic bacteria isolated from human stool samples. Archives of Microbiology. 2022;204:1–9.

42. Tyson GW, Lo I, Baker BJ, Allen EE, Hugenholtz P, Banfield JF. Genome-directed isolation of the key nitrogen fixer Leptospirillum ferrodiazotrophum sp. nov. from an acidophilic microbial community. Appl Environ Microbiol. 2005;71(10):6319–6324. doi:10.1128/AEM.71.10.6319-6324.2005

43. Eichele DD, Kharbanda KK. Dextran sodium sulfate colitis murine model: An indispensable tool for advancing our understanding of inflammatory bowel diseases pathogenesis. World J Gastroenterol. 2017;23(33):6016–6029. doi:10.3748/wjg.v23.i33.6016

44. Bruscoli S, Febo M, Riccardi C, Migliorati G. Glucocorticoid therapy in inflammatory bowel disease: mechanisms and clinical practice. Frontiers in immunology. 2021;12:691480.

45. Yamamoto A, Itoh T, Nasu R, Nishida R. Effect of sodium alginate on dextran sulfate sodium– and 2,4,6-trinitrobenzene sulfonic acid-induced experimental colitis in mice. Pharmacology. 2013;92(1-2):108–116. doi:10.1159/000353192

46. Bu F, Ding Y, Chen T, et al. Total flavone of Abelmoschus Manihot improves colitis by promoting the growth of Akkermansia in mice. Scientific Reports. 2021;11(1):20787.

47. Hugenholtz F, de Vos WM. Mouse models for human intestinal microbiota research: a critical evaluation. Cellular and Molecular Life Sciences. 2018;75:149–160.

48. Antoniou E, Margonis GA, Angelou A, et al. The TNBS-induced colitis animal model: An overview. Annals of medicine and surgery. 2016;11:9–15.

49. Molnár T, Farkas K, Szepes Z, et al. Long-term outcome of cyclosporin rescue therapy in acute, steroid-refractory severe ulcerative colitis. United European gastroenterology journal. 2014;2(2):108–112.

50. Evirgen S, İliaz R, Akyüz F, et al. Cyclosporine therapy as a rescue treatment in steroid refractory acute severe ulcerative colitis: a real life data from a tertiary center. The Turkish Journal of Gastroenterology. 2022;33(6):463.

51. Xiao C, Li G, Li X, et al. A topical thermosensitive hydrogel system with cyclosporine A PEG-PCL micelles alleviates ulcerative colitis induced by TNBS in mice. Drug Delivery and Translational Research. Published online 2023:1–16.

52. Otte ML, Tamang RL, Papapanagiotou J, Ahmad R, Dhawan P, Singh AB. Mucosal healing and inflammatory bowel disease: Therapeutic implications and new targets. World Journal of Gastroenterology. 2023;29(7):1157.

53. Colosimo DA, Kohn JA, Luo PM, et al. Mapping Interactions of Microbial Metabolites with Human G-Protein-Coupled Receptors. Cell Host Microbe. 2019;26(2):273–282.e7. doi:10.1016/j.chom.2019.07.002

54. Piscotta FJ, Hoffmann HH, Choi YJ, et al. Metabolites with SARS-CoV-2 inhibitory activity identified from human microbiome commensals. Msphere. 2021;6(6):e00711–21.

55. You J, Nguyen AV, Albers CG, Lin F, Holcombe RF. Wnt pathway-related gene expression in inflammatory bowel disease. Digestive diseases and sciences. 2008;53:1013–1019.

56. Moparthi L, Koch S. Wnt signaling in intestinal inflammation. Differentiation. 2019;108:24–32.

57. Shahini A, Shahini A. Role of interleukin-6-mediated inflammation in the pathogenesis of inflammatory bowel disease: Focus on the available therapeutic approaches and gut microbiome. Journal of Cell Communication and Signaling. 2023;17(1):55–74.

58. Langer V, Vivi E, Regensburger D, et al. IFN-γ drives inflammatory bowel disease pathogenesis through VE-cadherin–directed vascular barrier disruption. The Journal of clinical investigation. 2019;129(11):4691–4707.

59. Zihni C, Mills C, Matter K, Balda MS. Tight junctions: from simple barriers to multifunctional molecular gates. Nat Rev Mol Cell Biol. 2016;17(9):564–580. doi:10.1038/nrm.2016.80

60. Chelakkot C, Ghim J, Ryu SH. Mechanisms regulating intestinal barrier integrity and its pathological implications. Experimental & molecular medicine. 2018;50(8):1–9.

61. Suzuki T. Regulation of intestinal epithelial permeability by tight junctions. Cellular and molecular life sciences. 2013;70:631–659.

62. Neurath MF, Artis D, Becker C. The intestinal barrier: a pivotal role in health, inflammation, and cancer. The Lancet Gastroenterology & Hepatology. Published online 2025.

63. Atreya R, Neurath MF. Involvement of IL-6 in the pathogenesis of inflammatory bowel disease and colon cancer. Clin Rev Allergy Immunol. 2005;28(3):187–196. doi:10.1385/CRIAI:28:3:187

64. Peters A, Krumbholz P, Jäger E, et al. Metabolites of lactic acid bacteria present in fermented foods are highly potent agonists of human hydroxycarboxylic acid receptor 3. PLoS genetics. 2019;15(5):e1008145.

65. Ehrlich AM, Pacheco AR, Henrick BM, et al. Indole-3-lactic acid associated with Bifidobacterium-dominated microbiota significantly decreases inflammation in intestinal epithelial cells. BMC microbiology. 2020;20:1–13.

66. Laukova M, Glatman Zaretsky A. Regulatory T cells as a therapeutic approach for inflammatory bowel disease. European Journal of Immunology. 2023;53(2):2250007.

67. Meng F, Jiang X, Wang X, et al. Tumor necrosis factor–like cytokine 1A plays a role in inflammatory bowel disease pathogenesis. Proceedings of the National Academy of Sciences. 2023;120(34):e2120771120.

68. Furfaro F, Alfarone L, Gilardi D, et al. TL1A: a new potential target in the treatment of inflammatory bowel disease. Current Drug Targets. 2021;22(7):760–769.

69. Wang J, Al-Lamki RS, Zhu X, Liu H, Pober JS, Bradley JR. TL1-A can engage death receptor-3 and activate NF-kappa B in endothelial cells. BMC nephrology. 2014;15:1–10.

70. Sokol H, Pigneur B, Watterlot L, et al. Faecalibacterium prausnitzii is an anti-inflammatory commensal bacterium identified by gut microbiota analysis of Crohn disease patients. Proc Natl Acad Sci U S A. 2008;105(43):16731–16736. doi:10.1073/pnas.0804812105

71. Foligne B, Nutten S, Grangette C, et al. Correlation between in vitro and in vivo immunomodulatory properties of lactic acid bacteria. World journal of gastroenterology: WJG. 2007;13(2):236.

72. Food and Drug Administration. Early clinical trials with live biotherapeutic products: chemistry, manufacturing, and control information. Guidance for Industry. Published online 2016.

73. Marrs T, Walter J. Pros and cons: Is faecal microbiota transplantation a safe and efficient treatment option for gut dysbiosis? Allergy. 2021;76(7):2312–2317.

74. Merrick B, Allen L, Zain NMM, Forbes B, Shawcross DL, Goldenberg SD. Regulation, risk and safety of faecal microbiota transplant. Infection prevention in practice. 2020;2(3):100069.

75. Seres Therapeutics. Seres Therapeutics Announces Initiation of Phase 1b Trial of SER-301 for the Treatment of Ulcerative Colitis.

76. Vedanta Biosciences I. Vedanta Biosciences Announces Positive Topline Data from Two Phase 1 Studies of VE202, a Rationally Defined Bacterial Consortium Being Advanced for Inflammatory Bowel Diseases (IBD).

77. Microbiotica. Microbiotica Receives Crohn’s & Colitis Foundation Funding to Develop Ulcerative Colitis Therapeutic – Microbiotica.

78. McChalicher CW, Aunins JG. Drugging the microbiome and bacterial live biotherapeutic consortium production. Curr Opin Biotechnol. 2022;78:102801. doi:10.1016/j.copbio.2022.102801

79. Rouanet A, Bolca S, Bru A, et al. Live Biotherapeutic Products, A Road Map for Safety Assessment. Front Med (Lausanne). 2020;7:237. doi:10.3389/fmed.2020.00237

80. Cordaillat-Simmons M, Rouanet A, Pot B. Live biotherapeutic products: the importance of a defined regulatory framework. Exp Mol Med. 2020;52(9):1397–1406. doi:10.1038/s12276-020-0437-6

81. Delday M, Mulder I, Logan ET, Grant G. Bacteroides thetaiotaomicron ameliorates colon inflammation in preclinical models of Crohn’s disease. Inflammatory bowel diseases. 2019;25(1):85–96.

82. Ihekweazu FD, Fofanova TY, Queliza K, et al. Bacteroides ovatus ATCC 8483 monotherapy is superior to traditional fecal transplant and multi-strain bacteriotherapy in a murine colitis model. Gut microbes. 2019;10(4):504–520.

83. Hansen R, Sanderson IR, Muhammed R, et al. A double-blind, placebo-controlled trial to assess safety and tolerability of (thetanix) bacteroides thetaiotaomicron in adolescent crohn’s disease. Clinical and translational gastroenterology. 2021;12(1):e00287.

84. ClinicalTrials.gov. A Phase 1, Multicentre, 2-Part, Randomised, Parallel-Arm, Placebo-Controlled, Partially Double-Blind Study to Evaluate the Safety and Target Engagement of EXL01 in the Maintenance of Steroid-Induced Clinical Response or Remission in Participants With Mild to Moderate Crohn’s Disease. Identifier NCT05542355.; 2022. https://clinicaltrials.gov/study/NCT05542355#publications

85. Ulluwishewa D, Anderson RC, Young W, et al. Live F aecalibacterium prausnitzii in an apical anaerobic model of the intestinal epithelial barrier. Cellular microbiology. 2015;17(2):226–240.

86. Zhang J, Huang YJ, Yoon JY, et al. Primary human colonic mucosal barrier crosstalk with super oxygen-sensitive Faecalibacterium prausnitzii in continuous culture. Med. 2021;2(1):74–98.

87. Zhang J, Huang YJ, Trapecar M, et al. An immune-competent human gut microphysiological system enables inflammation-modulation by Faecalibacterium prausnitzii. npj Biofilms and Microbiomes. 2024;10(1):31.

88. Maier E, Anderson RC, Roy NC. Live Faecalibacterium prausnitzii does not enhance epithelial barrier integrity in an apical anaerobic co-culture model of the large intestine. Nutrients. 2017;9(12):1349.

89. Huang YJ, Lewis CA, Wright C, et al. Faecalibacterium prausnitzii A2-165 metabolizes host-and media-derived chemicals and induces transcriptional changes in colonic epithelium in GuMI human gut microphysiological system. Microbiome Research Reports. 2024;3(3):N-A.

90. Yu K, Li Q, Sun X, et al. Bacterial indole-3-lactic acid affects epithelium–macrophage crosstalk to regulate intestinal homeostasis. Proceedings of the National Academy of Sciences. 2023;120(45):e2309032120.

91. Laursen MF, Sakanaka M, von Burg N, et al. Bifidobacterium species associated with breastfeeding produce aromatic lactic acids in the infant gut. Nature microbiology. 2021;6(11):1367–1382.

92. Wilck N, Matus MG, Kearney SM, et al. Salt-responsive gut commensal modulates TH17 axis and disease. Nature. 2017;551(7682):585–589.

93. Dodd D, Spitzer MH, Van Treuren W, et al. A gut bacterial pathway metabolizes aromatic amino acids into nine circulating metabolites. Nature. 2017;551(7682):648-652.

