## Supplementary material for "A human microbiome-derived therapeutic for ulcerative colitis promotes mucosal healing and immune homeostasis": Methods

***DNA extraction***. DNA from microbial cultures was processed for direct entry to library preparation according to the Illumina DNA Prep crude lysate protocol for NGS (Illumina, USA). DNA was extracted from feces using the DNeasy 96 PowerSoil Pro QIAcube HT Kit (Qiagen; 47021) in 2 mL deep well plate format as per the manufacturer’s instructions, with optimised volumes of extraction buffer in the initial processing step on the QIAcube HT DNA extraction system (Qiagen; 9001793). Mechanical lysis was performed with PowerBead Pro beads (Qiagen) in 2 cycles of processing correcting for any position effects, enabling more efficient lysis of bacteria and fungi. Resulting DNA was quantitated using a Quant-iT^TM^ high sensitivity dsDNA fluorometric assay (ThermoFisher; Q33120). A minimum of 0.2 ng/µL DNA yield was required for samples to pass quality control.

***Metagenomic library preparation***. Libraries were constructed using the Illumina DNA Prep (M) Tagmentation Kit (Illumina; 20018705) with IDT for Illumina DNA/RNA UD Index Sets A-D (Illumina; 20027213-16) as per the manufacturer’s instructions, with a modification of volumes to accommodate processing in a 384-well plate format. Resulting libraries were assessed using a Quant-iT^TM^ high sensitivity dsDNA fluorometric assay (ThermoFisher; Q33120), and individual libraries visualized through capillary gel electrophoresis using the QIAxcel DNA High Resolution Kit (Qiagen). Libraries were required to meet minimum criteria for average size, smallest and largest fragment gating and concentration.

***Metagenomic sequencing.*** Individual libraries were pooled in equimolar amounts, thus creating a sequencing pool to be assessed through a Quant-iT^TM^ high sensitivity dsDNA fluorometric assay (ThermoFisher) and subsequently visualized through capillary gel electrophoresis using the QIAxcel DNA High Resolution Kit (Qiagen). Pools were required to meet minimum criteria for average size, smallest and largest fragment gating and concentration. Sequencing pools were loaded and sequenced using NovaSeq^TM^ 6000 (Illumina) coupled with v1.5 300 bp PE sequencing reagents as per the manufacturer’s instructions. Sequence data were initially reviewed for general minimum performance requirements for yield and sequence quality. Data were then further assessed for performance requirements of known control samples included in each processing run. Moreover, data were required to meet minimum requirements for contribution of reads from reagents (background contamination), and appropriate reporting of known control sample contents as assessed by Hellinger distance and associated statistical measures.

***Metagenomic sequencing data quality control.*** Metagenomic sequencing data quality control was performed by Microba Life Sciences Limited (www.microba.com). Paired-end DNA sequencing data were demultiplexed and adaptor trimmed using Illumina BaseSpace Bcl2fastq2 (v2.20), accepting one mismatch in index sequences. Obtained reads were trimmed, and residual adaptors excluded using the software Trimmomatic v0.39^1^ and the following parameters: -phred33 LEADING:3 TRAILING:3 SLIDINGWINDOW:4:15 CROP:100000 HEADCROP:0 MINLEN:100. Human DNA was identified and removed by aligning reads to the human genome reference assembly 38 (GRCh38.p12, GCF_000001405) using bwa-mem v0.7.17^2^ with default parameters except minimum seed length set to 31 (-k 31). Human genome alignments were filtered using SAMtools v1.7^3^ with flags -ubh -f1 -F2304. Read pairs where at least one read mapped to the human genome with >95% identity over >90% of the entire read length were flagged as human DNA and excluded from analysis. All samples were randomly subsampled to a standard depth of seven million read pairs.

***The Microba Genome Database (MGDB).*** Microba has established an extensive, high quality genome database, the Microba Genome Database (MGDB), which is used as a reference for the identification of microbial species from metagenomic sequence data. The database covers all prokaryotic lineages although it has been enriched with metagenome-assembled genomes (MAGs) mined from proprietary and publicly available metagenomes. Specifically, MGDB comprises 400,971 high quality prokaryotic genomes from the following sources: the Genome Taxonomy Database (GTDB, including GenBank and RefSeq genomes), metagenome-assembled genomes (MAGs) mined from proprietary samples collected under the Microba Future Insight Programme study (HREC 2018-05-400), MAGs mined by Microba from sequence read archive (SRA) samples, and MAGs and isolate genomes from previously published data sets^4–8^.

All included microbial genomes were quality controlled through rigorous standards, with an average completeness of 92.43% and average contamination of 1.15% as estimated with CheckM^9^ Genomes were then clustered into Operational Species Clusters and representative genomes selected to maximize the phylogenetic diversity within each species cluster. The resulting MGDB database contains 73,646 representative genomes grouped into 28,246 species clusters, of which, ~5,000 species clusters are observed at appreciable levels in fecal samples. Of these clusters, 625 were exclusively obtained by Microba mining efforts. Species represented in MGDB were systematically named using an in-house version of the Genome Taxonomy Database (GTDB)^10^.

***The Microba Community Profiler (MCP).*** The Microba Community Profiler (MCP), is a highly accurate, diagnostically accredited metagenomics taxonomic species profiler^11,12^. MCP determines bacterial, archaeal and eukaryotic species presence and estimates cellular relative abundance by aligning and interpreting metagenomic reads against the Microba Genome Database (MGDB). For prokaryote species, the taxonomic profiles produced by MCP are based on rank normalized taxa defined by the Genome Taxonomy Database^10^ and the expansive species clusters defined within MGDB. Eukaryote species profiles are based on curated sets of eukaryotic genomes classified using NCBI taxonomy^13^.

***Phylogenetic trees.*** A genome tree was constructed using gtdbtk (v2.4.0) from high quality species representative genomes (≥90%, contamination and ≤5% contamination from checkM analysis) from the *Acutalibacteriaceae* family (MGDB v4.1.0) and the MH27-2 isolate. For each genome, a set of 122 bacteria-specific conserved marker genes were extracted using gtdbtk^14^ *identify*. Genes were then aligned to profile Hidden Markov Models (HMMs), concatenated to a single alignment with gtdbtk *align*, and a Maximum likelihood phylogenetic tree was constructed from the alignments using FastTree^15^ (v. 2.1.10) with gtdbtk *infer*. Non-parametric bootstrap values were inferred using GenomeTreetk (v. 0.1.6) from 1000 repetitions. The resulting phylogenetic tree was visualised in R using ggtree^16^. Bar plots aligned to the phylogenetic plots representing the difference in relative abundance between Healthy and IBD cohorts for each species were calculated from the Microba Discovery Database as described in Krause *et al.* 2024^17^ (MDD; v2023).

***Metabolic reconstruction*.** Protein coding sequences were predicted and annotated using the annotate function in Microba Metabolics Inference (Mimir) pipeline (0.6.2). Briefly, Mimir identifies protein coding sequences using prodigal (v2.6.3)^22^, and annotates them using emapper (v2.1.3)^23^ to obtain E.C., PFam, TCDB and eggnog classifications. Hmmer hmmsearches (v3.1b2)^24^ against Pfam (release 33.0)^25^, TIGRFAM (release 15.0)^26^, and dbCAN2^27^ (downloaded September 2019) are then used to annotate functional domains, key metabolic markers and carbohydrate activate (CAZy) enzymes, respectively. Metabolic pathways are identified using the classify function in enrichM, thereby assessing annotations and their genomic position against manually defined metabolic pathway definitions. A pathway was considered present in a genome if it encoded >80% of the required proteins and it passed all required synteny checks. Lastly, the automatically predicted pathways were manually assessed and gutSMASH (version 1.0.0)^28^ applied to identify common functions mediated by gut microbiomes.

***Bacterial strains, culture conditions, and analyses.*** Stool samples were collected from healthy human adults with no history of gastrointestinal disorders and mixed with an equal amount (w/v) sterile oxygen-free glycerol solution^10^. Donors had not consumed antibiotics in the three months leading up to sample collection. All samples were collected following informed consent and in accordance with ethical guidelines approved by Bellberry Limited (HREC2018-05-400-A-6).

To isolate *Hominenteromicrobium mulieris* MH27-2, a donor fecal sample with *H. mulieris* present at a relative abundance of 0.38% was inoculated into custom medium (Alanine 0.4 g/L, tryptophan 0.08 g/L, methionine 0.2 g/L, phenylalanine 0.2 g/L, tyrosine 0.2 g/L, histidine 0.125 g/L, maltose 1 g/L, salt solution 2^29^, 75 mL/L, salt solution 3^29^, 75 mL/L, sodium bicarbonate 8 g/L, resazurin (0.1% w/v) 1mL/L, L-cysteine 1 g/L¸ Vitamin solution^29^ (1000x)) and serially diluted to extinction. The dilution-to-extinction culture series was sequenced, and an enrichment with *H. mulieris* at a relative abundance of 72.6% identified. The enrichment was subsequently streaked on PYG agar, and an isolate with a coccoid cell morphology identified. Following purification and whole genome sequencing, the resulting Gram-positive staining coccoid isolate was termed *H. mulieris* MH27-2.

*H. mulieris* and *Faecalibacterium prausnitzii* were routinely processed in a Coy vinyl anaerobic chamber with an oxygen-free (85% N_2_, 10% CO_2_, 5% H_2_) atmosphere. *H. mulieris* was routinely cultured in YG/V (Vegetable tryptone 20 g/L, yeast extract 10 g/L, glucose 10 g/L, salt solution 2^29^, 38 mL/L, salt solution 3^29^, 38 mL/L, sodium bicarbonate 8 g/L, resazurin (0.1% w/v) 1mL/L, L-cysteine 1 g/L) or YG/P medium (Phytone peptone 20 g/L, yeast extract 10 g/L, glucose 10 g/L, salt solution 2^29^, 38 mL/L, salt solution 3^29^, 38 mL/L, sodium bicarbonate 8 g/L, resazurin (0.1% w/v) 1mL/L, L-cysteine 1 g/L), and *F. prausnitzii* was cultured in TY medium (Tryptone 10 g/L, yeast extract 2.5 g/L, glucose 4 g/L, cellobiose 1 g/L, maltose 1 g/L, haemin (500 mg/L) 10 mL/L, acetic acid 1.9 mL/L, salt solution 2^29^, 38 mL/L, salt solution 3^29^, 38 mL/L, sodium bicarbonate 8 g/L, resazurin (0.1% w/v) 1mL/L, L-cysteine 1 g/L) or YG/V, as indicated. Isolates were stocked by combining 3 mL of actively growing culture with an equal volume of glycerol solution and storing at −80°C. *E. coli* was routinely cultured using Luria Bertani medium. All bacteria were grown at 37°C.

***In-silico safety analysis.*** *P*rotein coding sequences from the isolate genomes were assessed for the presence of antimicrobial resistance (AMR) genes using AMRFinderPlus^30^. Protein sequences were also searched against the Virulence Factor Database^31^ (downloaded August 2021). Significant hits to AMR and Virulence factor databases were further annotated with CD-SEARCH^32^ to verify the annotations. E.C. pathway annotations of the protein coding genes from mimir were used to assess the presence of biogenic amine biosynthesis pathways. A pathway was considered complete if all enzymes were present and was considered partially complete if key enzymes were present and one or more other enzymes were missing from the pathway. Isolate genome sequences were searched for mobile genetic elements (MGEs) using MobileElementFinder^33^ using default settings. Further, oriTFinder^34^ was used to identify known AMR genes, virulence factors, conjugation machinery and other features associated with the replication of mobile genetic elements such as the origin of transfer (*oriT*). Plasmids were identified using the default PlasForest pipeline^35^. Regions designated as potential mobile genetic elements were checked for the presence of AMR and virulence genes to assess the risk of horizontal gene transfer.

***Preparation of bacterial strains for animal experimentation***. *H. mulieris* MH27-2 and *F. prausnitzii* were grown to early stationary phase in YG/V and TY medium, respectively. Cell densities of the individual cultures were calculated using a Helber Counting Chamber. To prepare bacterial gavage solutions, bacteria were pelleted under a layer of sterile heavy mineral oil at 5,000 *g* for 10 minutes, and the cell-free supernatant discarded. Bacteria were then washed in 1.5 mL of sterile anaerobic buffered diluent (38 mL/L each of salt solutions 2 and 3^29^, 1 mL/L of 0.1% (w/v) resazurin solution, 1 g/L L-cysteine), and re-pelleted as described above. Finally, the washed cell pellets were resuspended in half strength glycerol solution (15% v/v glycerol solution in anoxic buffered diluent) to a final concentration of 1 x 10^9^ cells/mL, aliquoted, and stored at -80°C until required. Cell viability was confirmed by thawing a single aliquot and streaking on appropriate agar. The identity and purity of the individual strain preparations were confirmed by whole genome sequencing.

***Therapeutic model of Dextran Sulfate Sodium (DSS)-induced colitis.*** Five to eight-week-old C57BL/6 female mice purchased from Animal BioResources (New South Wales, Australia) were randomized and then co-housed for at least seven days prior to experimentation. Day zero was defined as the start of disease induction. To induce colitis, mice received 2.5% DSS *ad libitum* in their drinking water for five days. Naïve age-matched control mice were offered DSS-free drinking water. Treatments were administered for seven consecutive days, beginning on the final day of DSS exposure. For treatment with MH27-2, mice were first anesthetized using isoflurane, then orally gavaged with 0.2 x 10^9^ cells/day bacterial suspension in 200 µL of vehicle (15% v/v glycerol solution in anoxic buffered diluent) or vehicle control only. As a positive pharmacological control, prednisone (2 mg/kg) was intraperitoneally injected to select animals following anesthetization. Ten mice were allocated to each treatment group.

Body weight of the animals was measured daily and was reported as percentage (%) weight change relative to initial body weight (Day -1) in grams. Feces were collected and stool consistency recorded daily. To assess the extent of colonic mucosal inflammation, animals were examined with a small animal endoscope (Karl Storz Endoskope, Tuttlingen, Germany) on days -1, 3, 6 and 9, as previously described^36^.

Animal euthanasia was performed on day ten by sodium pentobarbitone overdose (300 mg/mL) by intra peritoneal injection. Blood was collected from animals via cardiac puncture, and the colon was harvested to measure length and weight. Fecal matter was removed from the colon and the presence of blood in feces was measured using Luminol solution (Sigma), as previously described^37^. Other tissues and organs were dissected out, and weights recorded following removal of fat and connective tissue.

For colon histopatholgy, tissue was rolled from the distal end into a roll and placed into 4% formalin for fixation. After 24 hours colon rolls were transferred to 70% ethanol for paraffin embedding before sectioning. Tissue sections were stained with hematoxylin and eosin (H&E) to assess disease pathology and separate sections were stained with Alcian blue to assess mucin production. Slides were imaged using the Aperio digital imaging system (Leica Biosystems, Nußloch, Germany).

Histological scoring was performed in accordance with a published method^36^.To grade colitis severity, the extent of inflammation and epithelial injury in the tissue sections were graded semi-quantitatively using an established scoring system^36^. For this, samples were randomized and blinded assessment conducted by a trained gastrointestinal pathologist. All procedures were approved by the University of Newcastle Animal Ethics Committee (A-2020-021).

For mouse faecal metagenome analysis, fresh faecal pellets were collected on day seven and ten, immediately frozen on dry ice and then stored at -80°C until further processing. DNA extraction, library preparation and sequencing were performed as described above. Fecal pellet metagenomes were processed using the MCP to determine community structure. PCoAs were generated from species read counts that were Hellinger transformed and scaled. Bray-Curtis distances were calculated between untreated and treated samples using the vegdist from the vegan package. Shannon diversity, richness and evenness were determined from the Hellinger-transformed read counts using vegan. Bray-Curtis distances, diversity, richness and evenness were compared between untreated and treated samples using Wilcoxon test. P-values were corrected for multiple testing using FDR.

***Prophylactic model of DSS-induced colitis.*** Female C57BL/6 mice, bred under specific pathogen-free (SPF) conditions and aged 13-14 weeks, were provided by BioLASCO Taiwan under Charles River Laboratory Licensee. Mice were bred under specific pathogen-free (SPF) conditions. Pups from multiple litters were weaned at 3 weeks of age, pooled and then housed with 10-15 mice per cage. Seven-week-old mice were shipped to Eurofins (Taiwan) and after 4 days acclimatization period, groups of mice were randomly assigned to their cage. The animal care and use protocol was reviewed and approved by the Institutional Animal Care and Use Committee (IACUC) at Pharmacology Discovery Services Taiwan, Ltd. All mice, except those in the control group, received 2.5% DSS in the drinking water for six days. Live MH27-2 at 0.2 x 10^9^ cells/day were administered daily by oral gavage starting one day prior to initiation of DSS challenge, and up to day seven. Prednisone at 2 mg/kg was administered by oral gavage for the same time frame.

During the study period, body weight, fecal occult blood and stool consistency were recorded from Day -1 to Day 7. At the termination on Day 7, animals were sacrificed approximately four hours after the last dose. Blood was collected from all animals by cardiac puncture, plasma samples were processed and stored -20^o^C for further analyses. Cecum tissues were harvested, and contents from cecum were collected and stored at -80^o^C. Colons were harvested, rinsed, photographed, weighed and its length measured. The colon length and colon-to-body weight ratio were calculated. The colon was then cut longitudinally to two parts from 4 cm from the anus; one part to be snap frozen with liquid nitrogen for possible biomarker, and the other part to be fixed in 10% formalin and kept in 70% ethanol for histopathology (Swiss-roll sections). Results were combined from two independent experiments, each with n=10 per treatment group (n=20/group in total).

The experimental setup for testing the efficacy of MAP315 in the prophylactic model of DSS-induced colitis was as described above. Details regarding the production of drug substance is outlined below (‘MAP315-001, a Phase I clinical study in healthy volunteers’). In addition to testing live MH27-2, Bacthera MAP315 drug substance was tested at three different doses (0.5 x 10^5^, 0.5 x 10^7^, and 0.5 x 10^8^ cells/day).

***TNBS model of murine colitis.*** BALB/c mice were bred under specific pathogen-free (SPF) conditions at BioLasco Taiwan (under Charles River Laboratories Licensee). Pups from multiple litters were weaned at 3 weeks of age, pooled and then housed with 10-15 mice per cage. Seven-week-old mice were shipped to Eurofins (Taiwan) and after 4 days acclimatization period, groups of mice were randomly assigned to their cage. The animal care and use protocol was reviewed and approved by the IACUC at Pharmacology Discovery Services Taiwan, Ltd. To induce disease, 5 or 10 male BALB/c mice were fasted overnight (Day 1) before TNBS challenge on Day 2. Distal colitis was induced by intracolonic instillation of TNBS (2,4,6 -Trinitrobenzene sulfonic acid solution, 1 mg in 0.1mL 50% ethanol), after which animals were kept in a vertical position for 30 s to ensure that the solution remained in the colon. Test article, MH27-2 at 0.2 x 10^9^ cells/day and vehicle (sterile glycerol and phosphate salt solution) were administered by oral gavage (PO) once daily (QD) starting from Day 1 (i.e., 1 day before TNBS challenge) to Day 5 for a total of 5 consecutive days (Days 1-5). The positive control Cyclosporin A was given at 75 mg/kg by oral gavage (PO) once daily from Day 1 (i.e., 1 day before TNBS challenge) to Day 4 for a total of 4 consecutive days (Days 1-4). On the day of TNBS challenge, vehicle (sterile glycerol and phosphate salt solution), MH27-2, and Cyclosporine A were given 2 hours before TNBS.

On Day 5, the mice were euthanized by CO_2_ asphyxiation, and blood was collected from all animals by cardiac puncture. Colons were weighed, and their lengths measured. Furthermore, when the abdominal cavity was opened before removal of the colon, adhesions between the colon and other organs were noted, as was the presence of colonic ulceration after removal and weighing of each colon. Macroscopic scoring was performed, and photos of the intact colons retrieved. Each colon was removed, rinsed, macroscopically scored^38^, photographed, weighed and its length measured, and then cut to two parts from 4 cm from the anus; one part fixed in 10% formalin and kept in 70% ethanol for histopathology, another part snap frozen with liquid nitrogen for cytokine analysis (TNF, IL-6, IL-12(p40), IFNγ and IL-17A) and MPO detection. For histopathology, four micrometre tissues sections were cut and stained with hematoxylin and eosin (H&E) for histological analysis under the light microscope (Leica DM2700 M, USA) as previously described^39^. Cecum tissues were harvested, and contents from cecums collected and stored at -80°C.

***Safety and tolerability of MAP315 drug substance in mice.*** The safety and tolerability of MAP315 drug substance was evaluated in 8–9-week-old C57BL/6 male and female mice. Mice received a 0.2 mL oral suspension of MAP315 at doses of either 5 x 10^6^ or 5 x 10^7^, or a vehicle control, once or twice daily over a 14-day period. During this period, body weight was monitored, and faecal samples were collected daily. Additionally, animals were closely observed for the presence of symptoms (e.g. mortality, convulsions, tremors, muscle relaxation, sedation) and autonomic effects (e.g. diarrhea, salivation, lacrimation, vasodilation, piloerection). Further, behaviour was observed, including alertness, vocalization, startle response, body and tail elevation, gait, convulsion, as well as noting skin colour, respiration, palpebral size and piloerection. Mice were sacrificed on day 15 and organs were harvested for histopathological evaluation, weight measurement and bacterial translocation. Blood was collected for analysis of standard hematologic and clinical chemistry parameters (e.g. leukocyte counts, hemoglobin, mean corpuscular volume, albumin, glucose, triglycerides), as well as bacterial translocation.

***Preparation of H. mulieris* *MH27-2 culture supernatant.*** Cultures for preparation of supernatants were prepared by inoculating three independent colonies into broth and incubating for 48 h in a 37°C water bath. Following incubation, the bacterial cultures were transferred to microcentrifuge tubes and centrifuged at 17,000 *g* for five minutes. The cell-free supernatant was then collected and stored at -20°C, or further processed using Vivaspin® 500 centrifugal concentrators, and size-fractionated through 3kDa filters according to the manufacturer’s instructions (Sartorius AG). The supernatant samples and size fractionated samples were stored as single use aliquots. All data presented are representative of three independent supernatant samples.

***Preparation and fractionation of H. mulieris* *MH27-2 metabolite extract.*** MH27-2 metabolite extracts were generated to enrich and concentrate bioactive compounds present in MH27-2 culture supernatants. For this, bacterial extracts or paired medium controls were prepared using an Amberlite^TM^ XAD-7HP or XAD-16 polymeric adsorbent (Sigma-Aldrich) essentially as previously described^40^. Briefly, MeOH-activated resin was added to 400 mL 3kDa-filtered cell-free MH27-2 supernatant (10% w/v) or medium controls, and the slurry was then gently shaken overnight at 4°C. Next, the resin was collected, washed with 400 mL deionized H_2_O, and combined with 120 mL 100% MeOH. The solution was incubated at room-temperature for 2 hours with gentle shaking, after which the MeOH solution was collected. A further 120 mL 100% MeOH was then added, and the incubation repeated as before. The two collected MeOH solutions were pooled and dried to completion under vacuum using a rotary evaporator, and the obtained extracts resuspended in 100% DMSO to a concentration of 100x of the original culture supernatant, and stored at -80°C.

To fractionate the MH27-2 metabolite extracts, the extracts or paired medium controls were generated as described above using Amberlite^TM^ XAD-16 resin (Sigma-Aldrich) and reconstituted in 10% DMSO/H_2_O to a 100x concentration. Extracts were then fractionated using a 1200 HPLC system (Agilent Technologies) coupled with an Eclipse XDB-C18 column (5 µm, 4.6 x 150 mm). A 5-95% acetonitrile gradient with 0.1% formic acid was performed at a flow rate of 0.8 mL/minute across 21 minutes. Detection was set to 280 nm, and fractions were collected every 30 seconds. Obtained fractions were freeze-dried (Martin Christ, Germany) and stored at -20 °C. Prior to experimental application, fractions were reconstituted in sterile H_2_O.

***Cell lines and culture conditions.*** IL-6- and IL-6/IL-6R-mediated STAT3 activity was assessed using the HEK-Blue^TM^ IL-6 reporter cell line (InvivoGen). IFNγ-mediated STAT1 activity was assessed using the HEK-Blue^TM^ IFNγ reporter cell line (InvivoGen). Cells were initially grown in DMEM medium supplemented with 10% heat-inactivated FBS and selective antibiotics blasticidin, zeocin, and normocin as directed by the manufacturer. Routine culture in the absence of selection antibiotics was then performed in the presence of 1% Penicillin-Streptomycin. Human HCT116 gut epithelial cells were sourced from CellBank Australia and grown in McCoy’s 5A medium supplemented with 10% heat-inactivated FBS and 1% Penicillin-Streptomycin. T84 and HT29 gut epithelial cells were sourced from CellBank Australia and cultured in DMEM/F12 supplemented with 10% heat-inactivated FBS and 1% Penicillin-Streptomycin. TF-1 erythroblasts were sourced from Sigma-Aldrich and grown in RPMI 1640 medium supplemented with 10% heat-inactivated FBS, 1% Penicillin-Streptomycin, 1% Sodium Pyruvate, 2mM L-glutamine and 5 ng/mL human recombinant GM-CSF (Peprotech). For adherent cells, media was refreshed every second day, and for cells in suspension 5 mL fresh media was added every second day. All cell lines were maintained at 37 °C, 5% CO_2_.

***Characterization of IL-6, IL-6/IL-6R, and IFNγ-mediated STAT signaling activity.*** To assess the effect of MH27-2 on the activity of IL-6- and IL-6/IL-6R-mediated STAT3 signaling, and IFNγ-mediated STAT1 signaling, HEK-Blue^TM^ IL-6 (InvivoGen^®^) and HEK-Blue^™^ IFNγ reporter cell lines (InvivoGen^®^) were seeded into flat-bottomed 96-well plates at a density of 50,000 cells/well, and incubated overnight. Cell-free bacterial supernatant or corresponding medium controls were added to the cells to a final concentration of 10% v/v, and incubated for 1 hour. Tofacitinib citrate (10 µM, Sellechchem) was simultaneously added as a positive control. Following incubation, cells were stimulated with recombinant human IL-6 (2 ng/mL; R&D systems), recombinant human IL-6/IL-6R complex (400 ng/mL; R&D systems), or recombinant human IFNγ (10 ng/mL; R&D systems), and incubated for 16 hours. STAT3- or STAT1-regulated SEAP reporter activity was subsequently assessed using QUANTI-Blue^TM^ as per the manufacturer’s instructions (InvivoGen^®^). Results are representative of three (IL-6, IFNγ) and one (IL-6/IL-6R) independent experiments, each assessing the effect of bacterial supernatants as three biological replicates in technical duplicates. Cytotoxicity was assessed using the CellTiter-Glo® 2.0 Cell Viability Assay (Promega) as per manufacturer’s instructions.

***Cell migration analyses.*** To assess the effect of MH27-2 on cell motility, IncuCyte^®^ wound healing and Transwell^®^ migration assays were utilized. For this, bacterial extracts were prepared using an Amberlite XAD-7HP polymeric adsorbent (Sigma Aldrich) essentially as described above. For Transwell^®^ assays, 100 µL of HCT116 cell suspension at a density of 3.5 x 10^5^/mL was added to the top compartments of 24-well plates coupled with 6.5 mm TC-treated polycarbonate membrane inserts (8 µm; Corning Costar), and 600 µL culture medium was added to the lower compartments. Cells were then incubated at 37°C, 5% CO_2_ for 24 hours, washed once with DPBS, and media refreshed with reduced serum (0.5% FBS) DMEM to suppress proliferation. MH27-2 metabolite extract (0.5x) or medium control was added to the lower compartment, and cells were incubated for a further 16 hours. Following incubation, cells were washed with DPBS, and the cells attached to the top of the membrane gently removed using a cotton tip. Remaining cells were fixed in 70% ethanol for 10 minutes and stained with 0.25% crystal violet for 5 minutes. The Transwell^®^ membranes were then washed with water, dried, mounted with 50% glycerol onto glass slides, and immediately imaged (Olympus DP73, Brightfield sRGB, 10x). Experiments were performed as biological and technical triplicates, capturing two images of each replicate. Successfully migrated cells were quantitated using ImageJ (Version 1.54d) and displayed as the average number of migrated cells in two microscopic fields per well.

For the IncuCyte® scratch wound assay, 6.0 x 10^4^ T84 cells were plated in 100 µL media on poly-L-ornithine-coated IncuCyte® ImageLock 96-well plates (Essen BioScience) and incubated for 7 days. Following incubation, the IncuCyte® WoundMaker tool was used to generate a homogeneous scratch in the cell monolayer. The cells were washed twice with DPBS and incubated in 200 µL DMEM/F12 medium supplemented with 10% heat-inactivated FBS and 1% Penicillin-Streptomycin for 24 hours. Cells were then pre-treated with 1x bacterial metabolite extract or medium control for one hour, before being stimulated with recombinant human IFNγ (10 ng/mL; R&D systems) and transferred to the IncuCyte® live-cell analysis system. Media and treatments were refreshed every 24 hours, and images captured every two hours until the wound was completely closed. Data analysis was performed using the integrated analysis software (v2020C).

***Cell proliferation analyses.*** HT29 cells were seeded into 96-well plates at a density of 6,000 cells/well, and T84 cells were seeded at a density of 10,000 cells/well and incubated overnight. For HCT116 cells, MH27-2 metabolite extract (2x) or medium control (2x) was then added, and the plate placed into the IncuCyte for label-free proliferation analysis which measures the percentage of the image area covered by cells (confluence) every two hours by the IncuCyte® live-cell analysis system. For HT29 and T84 cells, cells were pre-treated with 1x bacterial metabolite extract or medium control for one hour, before being stimulated with recombinant human IFNγ (10 ng/mL; R&D systems) and transferred to the IncuCyte® live-cell analysis system. Media and treatments were refreshed every 24 hours, and images captured every two hours until the cells were completely confluent. Data analysis was performed using the integrated analysis software (v2020C).

***Trans-epithelial electrical resistance (TEER) analyses*.** T84 cells were seeded onto 24-well Millicell^®^ polycarbonate cell culture inserts with 0.4 µm pore size (Sigma-Aldrich; PSHT010R5) at a density of 60,000 cells/well in 400 µL medium. A further 22 mL medium was added to the single-well feeder tray. Media was refreshed every second day. After 7 days of culture, inserts were transferred from the feeder tray to a 24-well Millicell^®^ receiver tray (Sigma-Aldrich; PSMW010R5), and 800 µL media was added to each bottom compartment. TEER values were recorded every 24 hours using the Millicell^®^ ERS-2 Voltohmmeter, and experiments initiated once all wells reached stable TEER readings above 2000 Ω.

For experiments, 1x MH27-2 metabolite extract or medium control, or indole-3-lactic acid (ILA; 62.5, 125, 250, 500, 1000 µM) diluted in cell culture medium was added to select apical compartments as three biological replicates either 1 h prior to addition of, or upon removal of barrier disruptor. To disrupt barrier integrity, recombinant human IL-6 (100 ng/mL; R&D systems) or recombinant human IFNγ (100 ng/mL; R&D systems) was added basolaterally, and cells incubated for 48 hours. Cells were then washed twice with PBS, and fresh media alone, or media supplemented with metabolite extract or ILA added before the cells were returned to the incubator. TEER values were recorded, and the medium refreshed every 24 hours.

***FITC-Dextran permeability assay.*** The paracellular flux of 10kDa Fluorescein Isothiocyanate (FITC)-Dextran (Sigma-Aldrich; FD10S) across T84 monolayers was assessed at the TEER experiment end points. For this, cells were first washed once with Hanks' Balanced Salt Solution (HBSS), then incubated in HBSS for 30 minutes at 37°C in a 5% CO_2_ atmosphere. HBSS was then removed, and 500 µL/well fresh HBSS was added to the basolateral compartments, and 200 µL/well HBSS containing 1 mg/mL 10kDa FITC-Dextran added to the apical compartments. Following incubation for 1 hour, 200 µL HBSS was transferred from the basolateral compartments to a black 96-well plate, and the fluorescence measured at an excitation of 485 nm and emission of 535 nm.

***ZO-1 quantitation by immunofluorescence (IF) microscopy.*** T84 cells were seeded onto trans-wells in 24-well plates at a density of 60,000 cells/well in 400 µL cell culture medium. Cells were incubated for ten days, and medium refreshed every second day. To induce loss of ZO-1, cells were stimulated with recombinant human IFNγ (100 ng/mL; R&D systems) in the absence or presence of 1x MH27-2 metabolite extract or medium control, Indole-3-lactic acid (ILA; 0.25, 0.5, 1 mM), Hydroxyphenyllactic acid (HPLA; 0.25, 0.5, 1 mM), D-(+)-3-phenyllactic acid (DPLA; 0.25, 0.5, 1 mM), L-(-)-3-phenyllactic acid (LPLA; 0.25, 0.5, 1 mM), or equivalent volume solvent control. Cells were incubated for 48 hours, then washed twice with PBS and fixed with 4% paraformaldehyde for 15 minutes. Cells were washed with PBS three times and stained while protected from light.

For staining, cells were permeabilized with 0.1% Triton X-100 in 5% BSA/PBS for 15 minutes, then washed three times with PBS and incubated for 2 hours in 5% BSA/PBS. Next, membranes were incubated with primary rabbit anti-human ZO-1 antibodies (5 µg/mL; ThermoFisher Scientific; 61-7300) in 5% BSA/PBS for 2 hours, washed three times with PBS, and incubated with Alexa Fluor® 488-conjugated goat anti-rabbit IgG H&L secondary antibody (1:2000; ThermoFisher Scientific; A-11008) and 4',6-diamidino-2-phenylindole, dihydrochloride (DAPI) (1:1000; ThermoFisher Scientific; 62247) in 5% BSA/PBS for 1 hour. Membranes were washed three times with PBS and once with ultrapure water, then excised and mounted under coverslips using ProLong Gold Antifade Mountant (ThermoFisher Scientific; P36930). Image acquisition was performed using an inverted and fully motorized Nikon/Spectral Spinning Disc Confocal microscope (X-1 Yokogawa spinning disc with Borealis modification) with a 40x NA 1.49 Plan Apochromat oil immersion objective lens (Nikon). Images were acquired across the cell using a coupled device (CCD) camera (Andow Clara) and the Nikon elements imaging software (Nikon, Version 4.40). For each condition, four separate images were captured. Maximum projection images were assembled, and relative brightness of stained cells quantified for each image using Fiji ImageJ (Version 1.53t) and normalized to unstimulated control cells.

***Generating murine colonic organoids.*** Ten-week-old C57BL/6 male mice (Australian Resource Centre) were housed under pathogen-free conditions, and sacrificed by CO_2_ asphyxiation. Proximal colons were harvested, flushed with cold DPBS, and cut open lengthwise. To isolate colonic crypts, colons were first washed three times with cold DPBS, then cut into 2 mm wide pieces into 50 mL tubes containing 15 mL cold DPBS. Using a 10 mL serological pipette pre-wetted with anti-adherence rinsing solution (STEMCELL Technologies; 07010), the tissue pieces were gently pipetted up and down three times. The pieces were then allowed to settle by gravity, the supernatant removed, and replaced with 15 mL fresh cold DPBS. Washing of the tissue pieces was repeated as described above 15-20 times until the supernatant ran clear. Following the last wash, tissue pieces were resuspended in 25 mL gentle cell dissociation media (STEMCELL Technologies; 100-0485), and incubated at room-temperature on a rocking platform for 15 minutes. The tissue pieces were allowed to settle by gravity, after which the dissociation media was removed, and the tissues resuspended in 10 mL cold DPBS/0.1% Bovine Serum Albumin (BSA; Sigma Aldrich). Tissues were pipetted up and down three times using a 10 mL serological pipette pre-wetted with anti-adherence rinsing solution, the DPBS/0.1% BSA recovered, and passed through a 70 µm filter (PluriSelect) into a 50 mL tube, thereby generating fraction one. Collection was repeated three more times using fresh DPBS/0.1% BSA, filters, and tubes, generating fractions two to four. Tubes were then centrifuged at 4°C, 200 *g* for 3 minutes, supernatants discarded, and pellets resuspended in 10 mL cold DMEM/F12 (Gibco). The fraction with the highest number of intact isolated crypts as assessed through microscopy was selected for organoid generation.

To generate intestinal organoids, crypts were centrifuged at 4°C, 200 *g* for 3 minutes, the supernatant discarded, and crypts resuspended in Intesticult^TM^ Organoid Growth Medium (STEMCELL Technologies; 06005) and Geltrex^TM^ LDEV-free reduced growth factor basement membrane matrix (Gibco; A1413301) (1/9 ratio) to a concentration of 5,000 crypts/mL. The crypt suspension was then seeded into a pre-warmed 24-well cell culture plate (Corning) at a volume of 50 µL/well using pipette tips pre-wetted with anti-adherence rinsing solution, and the plate incubated upside down at 37°C, 5% CO_2_ for 10 minutes. To each well, 750 µL Intesticult^TM^ Organoid Growth Medium was then added, and crypts incubated for a further 7-10 days until fully mature organoids. Media was refreshed every second day.

***Gene expression analyses.*** T84 cells (1.5 x 10^5^/well) were seeded into 24-well plates, incubated at 37°C, 5% CO_2_ for ten days, with media refreshed every second day. Cells were then stimulated with recombinant human IFNγ (100 ng/mL; R&D systems) in the presence or absence of 1x MH27-2 metabolite extract or medium control, ILA (0.5 mM), HPLA (0.5 mM), DPLA (0.5 mM), LPLA (0.5 mM), or equivalent volume solvent control for 24 hours, then washed with DPBS. RNA was extracted using the RNeasy Micro kit (Qiagen; 74004) as per the manufacturer’s instructions, and RNA concentrations assessed using a NanoDrop Lite Spectrophotometer. To generate complimentary DNA (cDNA), 1,000 nanograms of RNA was reverse transcribed using iScript^TM^ cDNA synthesis kit (Bio-Rad; 1708891), and diluted 1:3 in Nuclease-free water (Ambion). mRNA expression was analyzed through Quantitative real-time PCR (QuantStudio 7 Flex, Applied Biosystems) in 384-well plates (Applied Biosystems), using SYBR Green Real Time Master Mix (Applied Biosciences) and gene-specific primers designed using the Integrated DNA Technologies tool (IDT). Primers are detailed in Table 1. Target gene expression was normalized to the expression of the house keeping gene *HPRT* (*HPRT*; 2^[Ct^ *^HPRT^* ^- Ct^ *^gene^*^]^). Data were analyzed using SDS 2.5 software (Applied Biosystems).

For gene expression analyses of intestinal organoids, organoids were generated as three biological replicates as described above and challenged as three technical replicates. For stimulation, organoids were incubated in Intesticult^TM^ Organoid Growth Medium (STEMCELL Technologies) supplemented with recombinant murine IFNγ (100 ng/mL; R&D systems) in the absence or presence of 1x MH27-2 metabolite extract or medium control extract for 24 hours. Media was then aspirated, and the domes of the first technical replicates lysed with 500 µL TRIzol (ThermoFisher Scientific). Once lysed, the TRIzol was carried through to lyse the second and third replicates, thereby pooling the three replicates. The TRIzol was then transferred to microcentrifuge tubes, incubated at room temperature for 5 minutes, after which 100 µL chloroform was added. The tubes were shaken vigorously for 15 seconds, incubated at room temperature for a further two minutes, then centrifuged at 4°C and 12,000 *g* for 15 minutes and the upper phase gently transferred to fresh microcentrifuge tubes. To each tube, 250 µL isopropanol was added, followed by inversion two times, and incubation at 4°C for ten minutes. Tubes were centrifuged at 4°C, 15,000 *g*, 10 minutes, the supernatant discarded, and RNA pellets resuspended in 500 µL 80% (v/v) EtOH. Tubes were centrifuged at 4°C, 7,500 *g*, 5 minutes, the supernatant discarded, and the RNA pellet air dried before being resuspended in 30 µL RNase-free water. RNA was incubated at 57°C for 10 minutes, then stored at -80°C until gene expression analysis as described above using gene specific primers (Table 1).

***Assessment of Treg polarization by monocyte-derived dendritic cells*.** MH27-2 cell suspension samples for assessment of Treg polarization were prepared as follows. Briefly, a colony of MH27-2 was inoculated and grown until early stationary phase and bacterial cells in the culture were then enumerated using a Helber counter. To prepare a bacterial cell suspension, the culture was centrifuged at 4000 rpm for 10 minutes and the supernatant was then carefully discarded into a waste container while taking care not to dislodge the cell pellet. The cell pellet was resuspended in 1 mL of sterile anaerobic diluent solution (salt solution 2^29^, 38 mL/L, salt solution 3^29^, 38 mL/L, sodium bicarbonate 8 g/L, resazurin (0.1% w/v) 1mL/L, L-cysteine 1 g/L) and then transferred to clean sterile microfuge tubes. The microfuge tubes were centrifuged at 10,000g for 3 minutes and the supernatant was carefully removed. The cell pellet was then resuspended in 15% v/v anaerobic glycerol solution and brought to a final volume in a sterile vessel such that the final mixture had 1x10^8^ bacterial cells/mL. The homogenous cell suspension was then aliquoted into sterile anaerobic vials and stored as single use aliquots at -80°C, until required.

Monocyte derived dendritic cells (MoDC) were generated as previously described^41,42^. Briefly, classical monocytes were isolated from PBMCs obtained from healthy donors using the Human Classical Monocyte Isolation Kit (Miltenyi) as per the manufacturer’s instructions. Isolated monocytes were differentiated into immature monocyte derived dendritic cells (MoDC) by culturing for 6 days in complete RPMI medium supplemented with 50 ng/mL rh-GM-CSF and 20 ng/mL rh-IL-4, with half the medium being exchanged for fresh medium containing cytokines on day three. Immature MoDCs were harvested by trypsinization of the plates and washed extensively, before being dispensed into a 96-well plate at a density of 2 x 10^4^ cells/well. Bacterial cell suspensions were then applied to the MoDCs at bacterium:DC ratios of 2:1, 10:1 and 50:1, and the co-cultures incubated in a humidified CO_2_ incubator for 24 hours. As a control stimulation condition, select wells were challenged with Escherichia coli O111:B4 LPS (100 ng/mL) (Sigma Aldrich). Once MoDC had been matured for 24 hours by exposure to bacterial cells, autologous naïve CD4+ T cells were isolated from PBMCs using the Human Naïve CD4+ T cell Isolation Kit (Miltenyi) according to the manufacturer’s instructions. Naïve CD4+ T cells were subsequently co-cultured with mature MoDCs at a T cell:DC ratio of 2.5:1 for 6 days, after which the T cells were harvested and surface-stained using Zombie Aqua viability dye (BioLegend)as well as fluorescent antibodies against CD4 (BioLegend) and CD25 (BioLegend). Nuclear staining for FoxP3 was then performed using the True-Nuclear Transcription Factor Staining Kit (Biolegend) according to manufacturer’s instructions. Cells were acquired on a CytoFLEX flow cytometer and analyzed in FlowJo v10.

Each experiment was performed using eight individual PBMC donors, each assayed as technical duplicates that were averaged on a per-donor basis. Statistics were generated by repeated measures ANOVA in Graphpad Prism v9, which was also used to plot results.

***Metabolomic analyses.*** Untargeted metabolomic analyses of fractionated MH27-2 metabolite extracts were conducted using an UltiMate 3000 UHPLC (ThermoFisher Scientific) coupled to a Q Exactive^TM^ Plus MS (ThermoFisher Scientific). A 3-80% acetonitrile gradient with 0.1% formic acid was run at a flow rate of 0.25 μL/min across 95 minutes. An electrospray ionization interface served as the ionization source. MS data were acquired in data-dependent acquisition mode, starting with an MS1 survey scan from m/z 100 to 1500 at 35,000 resolutions. Up to ten MS2 scans in CID/HCD mode were acquired per cycle at 35,000 resolutions with one microscan in positive or negative mode. The maximum injection time was set to 100 ms, and the MS2 precursor window was 1.4 m/z. Normalized collision energy was set to a stepped mode combining energies of 20, 30, and 40 eV, with the default charge state set at z=1. MS2 experiments were automatically triggered within 5 to 10 seconds of their first occurrence at the peak apex. Dynamic exclusion time was set to 10 seconds, excluding ion species with unassigned charge states and isotope peaks. Further metabolic profile analysis and identification of key metabolites was conducted using the GNPS platform following established protocols^43^.

***Quantitation of aromatic lactic acids.*** To assess aromatic lactic acid production by MH27-2, a stock solution of indole-3-pyruvic acid (IPyA) was prepared in dimethyl sulfoxide (DMSO), while phenyl pyruvic acid (PPyA) and 4-Hydroxyphenyl Pyruvic Acid (HPPyA) were prepared in 1 M sodium hydroxide. YG/V medium was supplemented with 1 mM IPyA, PPyA, and HPPyA as required. Cultures were inoculated with 100 µL of seed culture and incubated anaerobically for 48 h at 37°C.

Quantitation of the aromatic lactic acid metabolites ILA, HPLA, and 3-phenyllactic acid (PLA) in 3kDa-filtered culture supernatant was performed using liquid chromatography with tandem mass spectrometry (LC-MS/MS). For this, UltiMate 3000 UHPLC (ThermoFisher Scientific) coupled to a Q Exactive^TM^ Plus MS (ThermoFisher Scientific) was utilized. A 3-80% acetonitrile gradient with 0.1% formic acid was run at a flow rate of 0.25 μL/min across 95 minutes. An electrospray ionization interface served as the ionization source. MS data were acquired in parallel with scheduled parallel reaction monitoring (PRM). MS1 scan parameters included an m/z range of 100 to 500, mass resolution of 35,000, AGC target of 3e6, and maximum injection time (IT) of 110 ms. PRM parameters featured a default charge state of 1, mass resolution of 17,500, AGC target of 2e5, maximum IT of 50 ms, isolation window of 1.4 m/z, and stepped normalized collision energies of 20, 30, and 40 eV. Quantification of metabolites was based on peak signal responses of the samples compared to a standard curve generated from authentic chemical standards with known concentrations.

The analysis of PLA D- and L-enantiomers was conducted at Q-MAP (University of Queensland). PLA D- and L-enantiomers were separated and quantified using the Vanquish Duo UHPLC-PDA system (Thermo Fisher Scientific) and data analysis performed using Chromeleon 7.3 software. Analyte separation was achieved on a Waters ACQUITY Premier HSS T3 column (100 x 2.1 mm, 1.8 µm ID, Waters Corp) with hydroxypropyl-β-cyclodextrin added to the mobile phase as a chiral selector. Metabolite quantification was based on peak signal responses of the samples against a standard curve generated from authentic chemical standards with known concentrations.

***Cloning and expression of MH27-2 LDH1 and LDH2 genes for recombinant protein production.*** Four genes of MH27-2 have been annotated with a lactate dehydrogenase (LDH) function (WP_308449175.1, WP_308449177.1, WP_343285358.1, WP_395507319.1). To assess the role of the candidate lactate dehydrogenases from MH27-2 in aromatic lactic acid production, the MH27-2 WP_308449175.1, WP_308449177.1, WP_343285358.1 and WP_395507319.1 genes were codon-optimized for expression in *Escherichia coli* using the NovoPro codon optimization software ([www.novoprolabs.com/tools/codon-optimization](http://www.novoprolabs.com/tools/codon-optimization)), and all endogenous NcoI and XhoI sites were eliminated. Then, the genes were custom synthesized by Biomatik (Cambridge, ON, Canada) with NcoI and XhoI restriction sites added to the 5′ and 3′ ends, respectively, and the fragment was cloned into the pET28b(+) vector (Novagen) for recombinant protein expression. The *Clostridium sporogenes* ATCC 15579 *fldH* and the *Bifidobacterium longum* ssp. *infantis* DSM20088 *Blon_RS05510* and *Blon_RS04275* were similarly cloned into the pET28b(+) vector.

Recombinant vectors carrying WP_308449175.1, WP_308449177.1, WP_343285358.1, WP_395507319.1, *fldH*, *Blon_RS05510* and *Blon_RS04275* were individually transformed into *E. coli* BL21 (DE3) competent cells and plated on Luria Bertani medium supplemented with 30 µg/mL kanamycin. For recombinant protein expression, *E. coli* BL21(DE3) carrying individual LDH genes were inoculated from independent colonies and the biological triplicate cultures were grown overnight at 37°C in 20 mL of autoinduction ZYM-5052 medium supplemented with 100 µg/mL kanamycin^44^. Following overnight growth, the medium was supplemented with 1 mM of either IPyA, PPyA or HPPyA, or vehicle as appropriate, and then incubated at 37°C for a further 3 hours. Then, cell free supernatant was prepared, and aromatic lactic acids were quantified as previously outlined.

***MAP315-001, a Phase I clinical study in healthy volunteers.*** To progress *H. mulieris* MH27-2 towards clinical trials, a current Good Manufacturing Practice (cGMP) process was developed alongside Bacthera A/S (Hørsholm, Denmark). Animal component-free media and production conditions were optimized to support clinical scale production under anaerobic conditions. The production process consisted of fermentation, harvest of bulk bacteria by tangential flow filtration (TFF), mixing with proprietary cryopreservatives, and lyophilization. The resulting substance, named MAP315 drug substance, was further formulated with excipients and encapsulated in enteric capsules at Bacthera AG (Basel, Switzerland) to produce the MAP315 drug product. The capsules were designed to resist the acidic environment within the stomach, but dissolve once in the small intestine. No materials of human or animal origin were used in the manufacturing process. Placebo capsules consisted of matching enteric capsules filled with inert excipients. Capsules were stored at 2–8°C, and their shelf-life established using standard stability studies.

The primary objective of the study was to evaluate the safety and tolerability of MAP315 when administered to healthy adult participants. The study was a Phase 1, single center, randomized, double-blind, placebo-controlled, multiple dose. The study was conducted according to the protocol and Good Clinical Practice (GCP). The study was reviewed by the Alfred Health Human Research Ethics Committee (HREC), with first approval granted March 29^th^ 2023. The study was notified to TGA through the clinical trial notification (CTN) scheme and acknowledgement received April 6^th^ 2023. The study was registered on ANZCTR: ACTRN12623000291684.

For the study, 32 participants (two cohorts of 16 participants each) were enrolled and randomized 3:1 to receive MAP315 or its matching placebo for 14 consecutive days. The study enrolled healthy male and female participants without past or current history of significant gastrointestinal (GI) tract and/or bowel disease; aged between 18 to 65 years inclusive at screening; weighing ≥50 kg and with a body mass index (BMI) between 18 and 32 kg/m^2^ inclusive at screening; and with the ability and willingness to attend the necessary visits at the study center. Participation in the study comprised of a screening period, admission to the clinical research unit (CRU), a dosing period, and a follow-up period. Screening was conducted between Day -29 and Day -2 (up to 28 days prior to enrollment) and admission to the CRU was on Day -1. During the dosing period, treatment (MAP315 or placebo) was administered from Day 1 to Day 14. Cohort 1, the low dose cohort, received one MAP315 or placebo capsule daily. Cohort 2, the high dose cohort, received eight MAP315 or placebo capsules daily (four capsules morning and evening). Both cohorts included two sentinel participants, one receiving MAP315 and one receiving placebo, with sentinels evaluated for at least 72 h before proceeding with dosing of the remaining cohort. Study participants were confined to the CRU from Day -1 until Day 3, upon which they were discharged. Participants returned to the CRU for scheduled visits throughout the dosing period, and for an end-of-study visit on Day 28. Participants were provided with a study diary to record adverse events (AEs), concomitant medication, and study drug administration compliance, as applicable.

MAP315 safety and tolerability was assessed by the incidence, nature, and severity of adverse events (AEs) during the study period as well as clinical laboratory safety analyses (hematology, coagulation, clinical chemistry, and urinalysis), vital signs (respiratory rate, heart rate, systolic and diastolic blood pressure [SBP and DBP], and body temperature), 12-lead electrocardiogram (ECG), and physical examination. Plasma samples were collected at Day 0 (prior to dosing), Day 7, Day 14 and Day 28 to evaluate potential translocation of MAP315 into the bloodstream by qPCR and C-reactive protein (CRP). Fecal samples were collected at Day 0 (prior to dosing), Day 1 (post-dose), Day 3, Day 7, Day 14 and Day 28, to evaluate calprotectin (a marker of GI inflammation). Clinical laboratory safety analyses and plasma CRP measurements were performed by Australian Clinical Laboratories (ACL) using standard pathology laboratory protocols. Plasma qPCR for MAP315 and fecal calprotectin assays were performed by Microba Life Sciences, Australia. Plasma RT-qPCR was performed using a validated assay, with a lower limit of quantitation (LLOQ) in plasma of 1000 organisms/mL sample. Calprotectin was evaluated using a standard commercial assay. The plasma RT-qPCR for the detection of MAP 315 was performed using a validated assay with a lower limit of quantitation (LLOQ) in plasma of 1000 equivalent organisms/mL.

**Table 1. Primer pairs designed for RT-qPCR**

| **Target  (*Homo sapiens*)** | **Fw. (5ʹ-3ʹ)** | **Rv. (5’-3’)** |
| --- | --- | --- |
| *HPRT* | TCAGGCAGTATAATCCAAAGATGGT | AGTCTGGCTTATATCCAACACTTCG |
| *CCL3* | TCTGCAACCAGTTCTCTGC | GGTTAGGAAGATGACACTGGG |
| *CCL5* | TGCCCACATCAAGGAGTATTTC | CCATCCTAGCTCATCTCCAAAG |
| *CXCL10* | CCTTATCTTTCTGACTCTAAGTGGC | ACGTGGACAAAATTGGCTTG |
| *IL6* | CCAGCTATGAACTCCTTCTC | GCTTGTTCCTCACATCTCTC |
| *ISG15* | ACTCATCTTTGCCAGTACAGG | CAGCTCTGACACCGACATG |
| *OCLDN* | ACAAGCGGTTTTATCCAGAGTC | GTCATCCACAGGCGAAGTTAAT |
| *ZO1* | CGGTCCTCTGAGCCTGTAAG | GGATCTACATGCGACGACAA |
| **Target  (*Mus musculus*)** | **Fw. (5ʹ-3ʹ)** | **Rv. (5’-3’)** |
| *Hprt* | CCCCAAAATGGTTAAGGTTG | AACAAAGTCTGGCCTGTATCC |
| *Ccl3* | AAGGTCTCCACCACTGCCCTTG | CTCAGGCATTCAGTTCCAGGTC |
| *Ccl5* | AGATCTCTGCAGCTGCCCTCA | GGAGCACTTGCTGCTGGTGTAG |
| *Ctnnb1* | GCTATTCCACGACTAGTTCAGC | AGCTCCAGTACACCCTTCTAC |
| *Cxcl10* | TCAGCACCATGAACCCAA | CTATGGCCCTCATTCTCACTG |
| *Il6* | GAGGATACCACTCCCAACAGACC | AAGTGCATCATCGTTGTTCATACA |
| *Isg15* | CTGAAGAAGCAGATTGCCCAGAAG | CGCTGCAGTTCTGTACCACTAGC |
| *Ocldn* | TTGGGACAGAGGCTATGG | ACCCACTCTTCAACATTGGG |
| *Wnt1* | ATTTTGCGCTGTGACCTCTT | AGCAACCTCCTTTCCCACTT |
| *Wnt3* | CCCGCTCAGCTATGAACAAG | ACTTTAGGTGCATGTGGTCC |
| *Wnt5a* | CGCTAGAGAAAGGGAACGAATC | CTCCATGACACTTACAGGCTAC |
| *Wnt5b* | GACTGACGCCAACTCCTG | TGCTCCTGATACAACTGACAC |
| *Wnt7a* | ACGAGTGTCAGTTTCAGTTCC | AATCGCATAGGTGAAGGCAG |
| *Wnt9b* | CCAAGAGAGGAAGCAAGGAC | AACAGGTACGAACAGCACAG |
| *Wnt10a* | CGCTTCTCTAAGGACTTTCTGG | GTGGCATTTGCACTTACGC |
| *Zo1* | GCTAAGAGCACAGCAATG GA | GCATGTTCAACGTTATCCAT |

**References**

1.	Bolger AM, Lohse M, Usadel B. Trimmomatic: a flexible trimmer for Illumina sequence data. *Bioinformatics*. 2014;30(15):2114-2120. doi:10.1093/bioinformatics/btu170

2.	Li H. Aligning sequence reads, clone sequences and assembly contigs with BWA-MEM. *arXiv preprint arXiv:13033997*. Published online 2013.

3.	Li H, Handsaker B, Wysoker A, et al. The sequence alignment/map format and SAMtools. *bioinformatics*. 2009;25(16):2078-2079.

4.	Almeida A, Mitchell AL, Boland M, et al. A new genomic blueprint of the human gut microbiota. *Nature*. 2019;568(7753):499-504.

5.	Forster SC, Kumar N, Anonye BO, et al. A human gut bacterial genome and culture collection for improved metagenomic analyses. *Nature biotechnology*. 2019;37(2):186-192.

6.	Nayfach S, Shi ZJ, Seshadri R, Pollard KS, Kyrpides NC. New insights from uncultivated genomes of the global human gut microbiome. *Nature*. 2019;568(7753):505-510. doi:10.1038/s41586-019-1058-x

7.	Pasolli E, Asnicar F, Manara S, et al. Extensive unexplored human microbiome diversity revealed by over 150,000 genomes from metagenomes spanning age, geography, and lifestyle. *Cell*. 2019;176(3):649-662.

8.	Zou Y, Xue W, Luo G, et al. 1,520 reference genomes from cultivated human gut bacteria enable functional microbiome analyses. *Nature biotechnology*. 2019;37(2):179-185.

9.	Parks DH, Imelfort M, Skennerton CT, Hugenholtz P, Tyson GW. CheckM: assessing the quality of microbial genomes recovered from isolates, single cells, and metagenomes. *Genome Res*. 2015;25(7):1043-1055. doi:10.1101/gr.186072.114

10.	Parks DH, Chuvochina M, Waite DW, et al. A standardized bacterial taxonomy based on genome phylogeny substantially revises the tree of life. *Nat Biotechnol*. 2018;36(10):996-1004. doi:10.1038/nbt.4229

11.	Parks DH, Rigato F, Vera-Wolf P, et al. Evaluation of the Microba Community Profiler for Taxonomic Profiling of Metagenomic Datasets From the Human Gut Microbiome. *Front Microbiol*. 2021;12:643682. doi:10.3389/fmicb.2021.643682

12.	Angel NZ, Sullivan MJ, Alsheikh-Hussain A, et al. Metagenomics: a new frontier for routine pathology testing of gastrointestinal pathogens. *Gut Pathogens*. 2025;17(1):4.

13.	Schoch CL, Ciufo S, Domrachev M, et al. NCBI Taxonomy: a comprehensive update on curation, resources and tools. *Database*. 2020;2020:baaa062.

14.	Chaumeil PA, Mussig AJ, Hugenholtz P, Parks DH. GTDB-Tk v2: memory friendly classification with the genome taxonomy database. *Bioinformatics*. 2022;38(23):5315-5316.

15.	Price MN, Dehal PS, Arkin AP. FastTree 2–approximately maximum-likelihood trees for large alignments. *PloS one*. 2010;5(3):e9490.

16.	Yu G, Smith DK, Zhu H, Guan Y, Lam TT. ggtree: an R package for visualization and annotation of phylogenetic trees with their covariates and other associated data. *Methods in Ecology and Evolution*. 2017;8(1):28-36.

17.	Krause L, Boyd J, Vivian C, et al. A rational approach for the targeted discovery and characterisation of microbiome-derived therapeutics. *bioRxiv*. Published online 2024:2024-10.

18.	Caspi R, Billington R, Keseler IM, et al. The MetaCyc database of metabolic pathways and enzymes-a 2019 update. *Nucleic acids research*. 2020;48(D1):D445-D453.

19.	Suzek BE, Wang Y, Huang H, McGarvey PB, Wu CH, UniProt Consortium. UniRef clusters: a comprehensive and scalable alternative for improving sequence similarity searches. *Bioinformatics*. 2015;31(6):926-932.

20.	Steinegger M, Söding J. MMseqs2 enables sensitive protein sequence searching for the analysis of massive data sets. *Nature biotechnology*. 2017;35(11):1026-1028.

21.	Saier Jr MH, Reddy VS, Tsu BV, Ahmed MS, Li C, Moreno-Hagelsieb G. The transporter classification database (TCDB): recent advances. *Nucleic acids research*. 2016;44(D1):D372-D379.

22.	Larralde M. Pyrodigal: Python bindings and interface to Prodigal, an efficient method for gene prediction in prokaryotes. *Journal of Open Source Software*. 2022;7(72):4296.

23.	Cantalapiedra CP, Hernández-Plaza A, Letunic I, Bork P, Huerta-Cepas J. eggNOG-mapper v2: functional annotation, orthology assignments, and domain prediction at the metagenomic scale. *Molecular biology and evolution*. 2021;38(12):5825-5829.

24.	Finn RD, Clements J, Eddy SR. HMMER web server: interactive sequence similarity searching. *Nucleic acids research*. 2011;39(suppl_2):W29-W37.

25.	Finn RD, Bateman A, Clements J, et al. Pfam: the protein families database. *Nucleic acids research*. 2014;42(D1):D222-D230.

26.	Haft DH, Loftus BJ, Richardson DL, et al. TIGRFAMs: a protein family resource for the functional identification of proteins. *Nucleic acids research*. 2001;29(1):41-43.

27.	Zhang H, Yohe T, Huang L, et al. dbCAN2: a meta server for automated carbohydrate-active enzyme annotation. *Nucleic acids research*. 2018;46(W1):W95-W101.

28.	Pascal Andreu V, Augustijn HE, Chen L, et al. gutSMASH predicts specialized primary metabolic pathways from the human gut microbiota. *Nature Biotechnology*. Published online 2023:1-8.

29.	Makkar HP, McSweeney CS. *Methods in Gut Microbial Ecology for Ruminants*. Vol 10. Springer; 2005.

30.	Feldgarden M, Brover V, Gonzalez-Escalona N, et al. AMRFinderPlus and the Reference Gene Catalog facilitate examination of the genomic links among antimicrobial resistance, stress response, and virulence. *Scientific reports*. 2021;11(1):12728.

31.	Chen L, Yang J, Yu J, et al. VFDB: a reference database for bacterial virulence factors. *Nucleic acids research*. 2005;33(suppl_1):D325-D328.

32.	Marchler-Bauer A, Bo Y, Han L, et al. CDD/SPARCLE: functional classification of proteins via subfamily domain architectures. *Nucleic acids research*. 2017;45(D1):D200-D203.

33.	Johansson MH, Bortolaia V, Tansirichaiya S, Aarestrup FM, Roberts AP, Petersen TN. Detection of mobile genetic elements associated with antibiotic resistance in Salmonella enterica using a newly developed web tool: MobileElementFinder. *Journal of Antimicrobial Chemotherapy*. 2021;76(1):101-109.

34.	Li X, Xie Y, Liu M, et al. oriTfinder: a web-based tool for the identification of origin of transfers in DNA sequences of bacterial mobile genetic elements. *Nucleic acids research*. 2018;46(W1):W229-W234.

35.	Pradier L, Tissot T, Fiston-Lavier AS, Bedhomme S. PlasForest: a homology-based random forest classifier for plasmid detection in genomic datasets. *BMC bioinformatics*. 2021;22(1):349.

36.	Marks E, Goggins BJ, Cardona J, et al. Oral delivery of prolyl hydroxylase inhibitor: AKB-4924 promotes localized mucosal healing in a mouse model of colitis. *Inflammatory bowel diseases*. 2015;21(2):267-275.

37.	Park AM, Tsunoda I. Forensic luminol reaction for detecting fecal occult blood in experimental mice. *Biotechniques*. 2018;65(4):227-230.

38.	Vilaseca J, Salas A, Guarner F, Rodriguez R, Martinez M, Malagelada J. Dietary fish oil reduces progression of chronic inflammatory lesions in a rat model of granulomatous colitis. *Gut*. 1990;31(5):539-544.

39.	Dieleman, Palmen, Akol, et al. Chronic experimental colitis induced by dextran sulphate sodium (DSS) is characterized by Th1 and Th2 cytokines. *Clinical & Experimental Immunology*. 1998;114(3):385-391.

40.	Colosimo DA, Kohn JA, Luo PM, et al. Mapping interactions of microbial metabolites with human G-protein-coupled receptors. *Cell host & microbe*. 2019;26(2):273-282.

41.	Chometon TQ, Siqueira M da S, Sant´ anna JC, et al. A protocol for rapid monocyte isolation and generation of singular human monocyte-derived dendritic cells. *PLoS One*. 2020;15(4):e0231132.

42.	Posch W, Lass-Flörl C, Wilflingseder D. Generation of human monocyte-derived dendritic cells from whole blood. *Journal of visualized experiments: JoVE*. 2016;(118):54968.

43.	Nothias LF, Petras D, Schmid R, et al. Feature-based molecular networking in the GNPS analysis environment. *Nature methods*. 2020;17(9):905-908.

44.	Studier FW. Protein production by auto-induction in high-density shaking cultures. *Protein expression and purification*. 2005;41(1):207-234.
