## Supplementary Information for "A human microbiome-derived therapeutic for ulcerative colitis promotes mucosal healing and immune homeostasis"

**Isolation of strain MH27-2**

Our analysis suggested that *Hominenteromicrobium* *mulieris* is a sugar fermenter capable of utilizing a wide range of carbohydrate sources including maltose, galactose and mannose. It is predicted to produce nearly all amino acids although several pathways were identified for the uptake and fermentation of amino acids (e.g. Arginine, Aspartate, Cysteine, Glutamic acid, Glutamine, Methionine, Serine, and Threonine). We adapted the medium previously used to isolate *H.* *mulieris* MH27-1 principally by adding maltose as the sole carbon source based on our prediction of the bacterium’s ability to use it. A dilution to extinction culture was established using a fecal sample with the target species at 0.38% relative abundance and following outgrowth, a single enrichment with *H.* *mulieris* at a 72% relative abundance was produced. An axenic strain, MH27-2, was subsequently isolated and presented as Gram-variable coccoid/ovoid cells, often occurring as pairs but occasionally also as short chains (**Supplementary Figure 1A**).

**Genomic and metabolic characterization of MH27-2**

In accordance with GDI predictions, MH27-2 was shown to utilize a wide range of carbohydrate sources and amino acids as energy sources, as assessed through API-50CH and API RAPID 32A (**Table S2**). The major end products of MH27-2 glucose metabolism were acetic (1.78 mM in sterile YG/P vs 36.61±1.58 mM in cell free supernatant) and succinic acids (1.43 mM in sterile YG/P vs 46.71±6.39 mM in cell free supernatant), with small quantities of formic acid (1.78 mM in sterile YG/P vs 2.89±2.40 mM in cell free supernatant) and iso-valeric acid (0 mM in sterile YG/P vs 0.23±0.05 mM in cell free supernatant) also produced.

**MH27-2 promotes wound healing and improves gut barrier integrity in human cell assays**

JAK-STAT signaling is central to mucosal immunity and epithelial integrity^1^. Given the ability of MH27-2 to ameliorate IFNγ- and IL-6-driven effects on gut barrier integrity, its ability to modulate the JAK-STAT signaling pathway was examined. IL-6 induces immune responses by interacting with either the membrane bound IL-6 receptor (IL-6R; IL-6 classical signaling), or a soluble form of IL-6R (IL-6 trans-signaling). IL-6 trans-signaling contributes to IBD pathogenesis and chronic inflammation in the gut due to its pro-inflammatory and anti-apoptotic effects on immune cells^2,3^. MH27-2 supernatant significantly suppressed STAT3 activation mediated by both IL-6 classical- and trans-signaling in HEK-Blue™ IL-6 reporter cells (**Figure S7D-E**). Similarly, MH27-2 supernatant, and to a lesser extent metabolite extract, suppressed IFNγ-mediated STAT1 activation in HEK-Blue™ IFNγ reporter cells (**Figure S7F**).

**MH27-2 produces aromatic lactic acids that support mucosal immunity and barrier integrity**

Four candidate lactate dehydrogenasese were identified in the MH27-2 genome. PEG.475and PEG.793 are annotated as L-lactate dehydrogenases (EC 1.1.1.27) while PEG.477 and PEG.2593 are annotated as D-lactate dehydrogenases (EC 1.1.1.28). Both PEG.477 and PEG.2591 exhibit >58% sequence similarity to FldH, a D-lactate dehydrogenase from *Clostridium sporogenes* ATCC 15579^4^. FldH mediates conversion of aromatic pyruvic acids to aromatic lactic acids suggesting that MH27-2 produces aromatic lactic acids by a similar mechanism. In contrast, no enzymes known to convert aromatic amino acids to their corresponding pyruvic acids or other aromatic acid derivatives, such as aromatic acrylic, acetic or propionic acids were identified. Consistent with these observations, MH27-2 was found to produce high quantities of indole-3-lactic acid (ILA) and 4-hydroxyphenyl-3-lactic acid (HPLA) when supplemented with the respective aromatic pyruvic acids (**Table S3**). Moreover, MH27-2 was shown to produce both D- and L- phenyllactic acid (PLA), with the latter primarily produced upon supplementation with phenylpyruvic acid (**Figure S8B**). Heterologous expression of PEG475, PEG.477, PEG793 and PEG.2591 individually in *Escherichia coli* BL21(DE3), revealed that aromatic lactic acid production was underpinned by PEG.2591 (**Table S4**). As expected, heterologous expression of the *C. sporogenes* ATCC 15579 FldH (Cs_FldH) and the type 4 LDH from *Bifidobacterium longum* ssp. *infantis* DSM20088 (Blon_RS05510 (Bl_05510)), but not the type 1 LDH from *B. longum* ssp. *infantis* DSM20088 (Blon_RS04275 (Bl_04275)), also resulted in aromatic lactic acid production (**Table S4**)^4,5^.

**References**

1.	Atreya, R. & Neurath, M. F. Involvement of IL-6 in the pathogenesis of inflammatory bowel disease and colon cancer. *Clin. Rev. Allergy Immunol.* **28**, 187–196 (2005).

2.	Mitsuyama, K., Sata, M. & Rose-John, S. Interleukin-6 trans-signaling in inflammatory bowel disease. *Cytokine Growth Factor Rev.* **17**, 451–461 (2006).

3.	Mudter, J. & Neurath, M. F. Il-6 signaling in inflammatory bowel disease: pathophysiological role and clinical relevance. *Inflamm. Bowel Dis.* **13**, 1016–1023 (2007).

4.	Dodd, D. *et al.* A gut bacterial pathway metabolizes aromatic amino acids into nine circulating metabolites. *Nature* **551**, 648–652 (2017).

5.	Laursen, M. F. *et al.* Bifidobacterium species associated with breastfeeding produce aromatic lactic acids in the infant gut. *Nat. Microbiol.* **6**, 1367–1382 (2021).
